# Molecular Programs of Glomerular Hyperfiltration in Early Diabetic Kidney Disease

**DOI:** 10.1101/2021.09.13.21263340

**Authors:** Vidar T. N. Stefansson, Viji Nair, Toralf Melsom, Helen C. Looker, Laura H. Mariani, Damian Fermin, Felix Eichinger, Rajasree Menon, Lalita Subramanian, Jennifer L. Harder, Jeffrey B. Hodgin, Peter J. Nelson, Bjørn O. Eriksen, Robert G. Nelson, Matthias Kretzler

## Abstract

Hyperfiltration (HF) is a state of high glomerular filtration rate (GFR) observed in early diabetes that damages glomeruli, resulting in an iterative process of increasing filtration load on fewer and fewer remaining functional glomeruli. To delineate underlying cellular mechanisms of damage induced by HF, transcriptional profiles of kidney biopsies from Pima Indians with type 2 diabetes with or without early-stage diabetic kidney disease (DKD) were grouped into two HF categories based on annual iothalamate GFR measurements. Twenty-six participants with a peak GFR measurement within two years of biopsy were categorized as the “HF group”, and 26 in whom biopsy preceded peak GFR by >2 years were considered “pre-HF”. The HF group had higher hemoglobin A1c, higher urine albumin-to-creatinine ratio, increased glomerular basement membrane width and lower podocyte density compared to the pre-HF group. A glomerular 1240-gene transcriptional signature identified in the HF group was enriched for endothelial stress response signaling genes, including from endothelin-1, tec-kinase and TGF-β1 pathways, with the majority of the transcripts mapped to endothelial and inflammatory cell clusters in kidney single cell transcriptional data. This analysis reveals molecular pathomechanisms contributing to development of HF and early DKD and involving putative ligand-receptor pairs and downstream intracellular targets linked to cellular crosstalk between endothelial and mesangial cells.

## Introduction

Diabetic kidney disease (DKD) is the most common cause of end-stage kidney disease (ESKD) in the United States with rising prevalence worldwide, in tandem with the obesity epidemic^1, 2^. Alterations in glomerular hemodynamic function at the onset of diabetes often lead to sustained increases in the glomerular filtration rate (GFR), commonly referred to as hyperfiltration (HF). The presence of HF is considered a key early driver of DKD as HF is associated with development and progression of DKD^3-5^ and with increased mortality^6^ and is considered a prime therapeutic target in DKD. HF may also be part of the etiology of some non-diabetic chronic kidney diseases including obesity-related glomerulopathy^7, 8^. The pathomechanisms triggered in HF are not fully understood. Proposed mechanisms include increased intra-renal nitric oxide signaling, tubular sodium and glucose reabsorption, and intraglomerular mechanical stress from glomerular hypertension^7, 9-12^. However compelling evidence on how these and other pathways are regulated in the kidneys in HF are yet to be established.

We explored pathomechanisms activated in HF in Pima Indians from the Gila River Indian Community in Arizona who have a very high prevalence of obesity, type 2 diabetes and DKD. A group of individuals from this community has participated in decades-long prospective studies of early DKD that included clinical data and serial measurements of GFR by iothalamate clearance. A subset of these participants also underwent research kidney biopsies. Previous studies in this cohort documented the presence of HF and established a temporal link to the onset of diabetes ^5, 13^. Although the diagnosis of HF has typically relied on measures of whole kidney GFR, without accounting for individual differences in nephron numbers, a previous study of kidney donors found similar levels of single-nephron GFR across a range of whole-kidney GFR, underlining the problem with absolute whole-kidney GFR HF thresholds^14^. Therefore, to more accurately reflect hyperfiltration at a single nephron level, we defined HF as peak GFR at the individual level based on observed trends during long-term follow-up. We then examined transcriptional differences in kidney tissue obtained from individuals who reached peak GFR around the time of kidney biopsy and in those who reached peak GFR well after biopsy to identify signatures associated with HF that may contribute to progression of DKD. The aim of the present study was to integrate clinical measurements, structural morphometry, and gene expression analyses of kidney tissue, including single cell RNAseq (scRNAseq) analyses to identify pathways activated by HF that may promote progressive DKD and be amenable to targeted therapies to prevent DKD. An overview of the analytical strategy used to delineate HF associated glomerular cell types, molecular pathways, ligands, intracellular targets and intercellular crosstalk in this study is shown in **Figure 1**.

**Figure 1.**
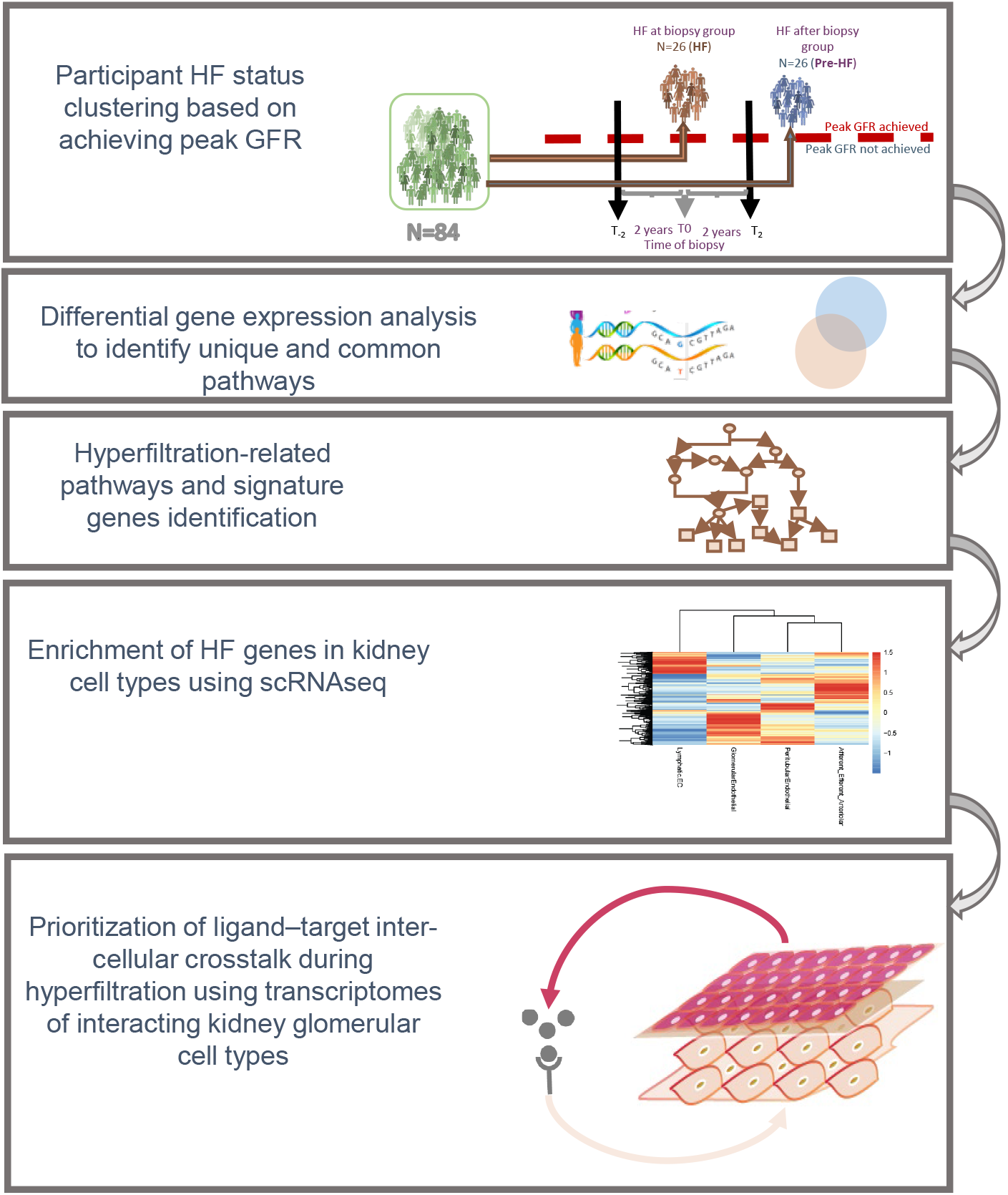
Explanatory figure describing the analytical approach of the study. Participants were clustered based on peak measured GFR and their kidney biopsy transcriptional profiles compared to identify hyperfiltration related pathways and the specific cells in the kidney involved in hyperfiltration related glomerular injury pathways.

## Methods

### Study population

Pima Indians from the Gila River Indian Community participated in a longitudinal study of diabetes and its complications between 1965 and 2007. In 1996, 169 Pima adults with type 2 diabetes participated in a prospective, randomized, placebo-controlled, double-blinded intervention trial (Renoprotection in Early Diabetic Nephropathy in Pima Indians trial, clinicaltrials.gov, NCT00340678) to ascertain the renoprotective efficacy of losartan, an angiotensin receptor blocker, relative to placebo^15^. At the end of the 6-year treatment trial, 111 of the participants underwent a kidney biopsy. All trial participants continued to undergo annual follow-up examinations after conclusion of the treatment trial. In 2015, 1^st^ degree relatives of clinical trial participants were also enrolled in the follow-up study, and tissue from kidney biopsies obtained at enrollment in 42 of them and in 2 clinical trial participants who had not previously been biopsied underwent scRNAseq analysis as described below. Follow-up of participants continued through 2019.

Each participant signed an informed consent document, and the study was approved by the Institutional Review Board of the National Institute of Diabetes and Digestive and Kidney Diseases. Due to ethical considerations including individual privacy protection concerns, the board stipulated that individual-level genotype and gene expression data from this study cannot be made publicly available.

### Peak GFR categorization

Individuals who had a mean GFR <60 ml/min and/or a mean urine albumin-to-creatinine ratio (ACR) >300 mg/g within 1 year of their kidney biopsy were considered to have advanced DKD and were excluded, leaving 84 participants considered for the present study. GFR measurements performed in all kidney study protocols were reviewed, and these participants were classified into 3 groups based on the date they had their highest recorded GFR measurement (peak GFR), regardless of the absolute GFR value. Twenty-six participants who achieved peak GFR <2 years from the time of biopsy were categorized as the “HF group”. The remaining 58 participants were split into two groups based on whether their peak was >2 years before (N=32) or >2 years after (N=26) the biopsy. Those whose GFR peak was >2 years before the biopsy were also considered likely to have more advanced DKD because of their declining GFR and were excluded from subsequent analysis, whereas the 26 participants with a peak >2 years after biopsy were included in the analyses and were referred to as the “Pre-HF group”. A flow chart of participant selection for this study is summarized in **Figure 2**. Seven participants reached the identical GFR peak twice. Of these, two had their peaks within the same time interval category whereas three participants had their initial peak >2 years before biopsy and the second peak within 2 years of biopsy, and two had their first peak within 2 years of biopsy and then a second peak >2 years after biopsy. In these cases, the time of the first GFR peak was used for the purpose of categorization. As a sensitivity analysis, peak GFR was also categorized based on creatinine-based eGFR computed using the CKD-EPI equation^16^.

**Figure 2.**
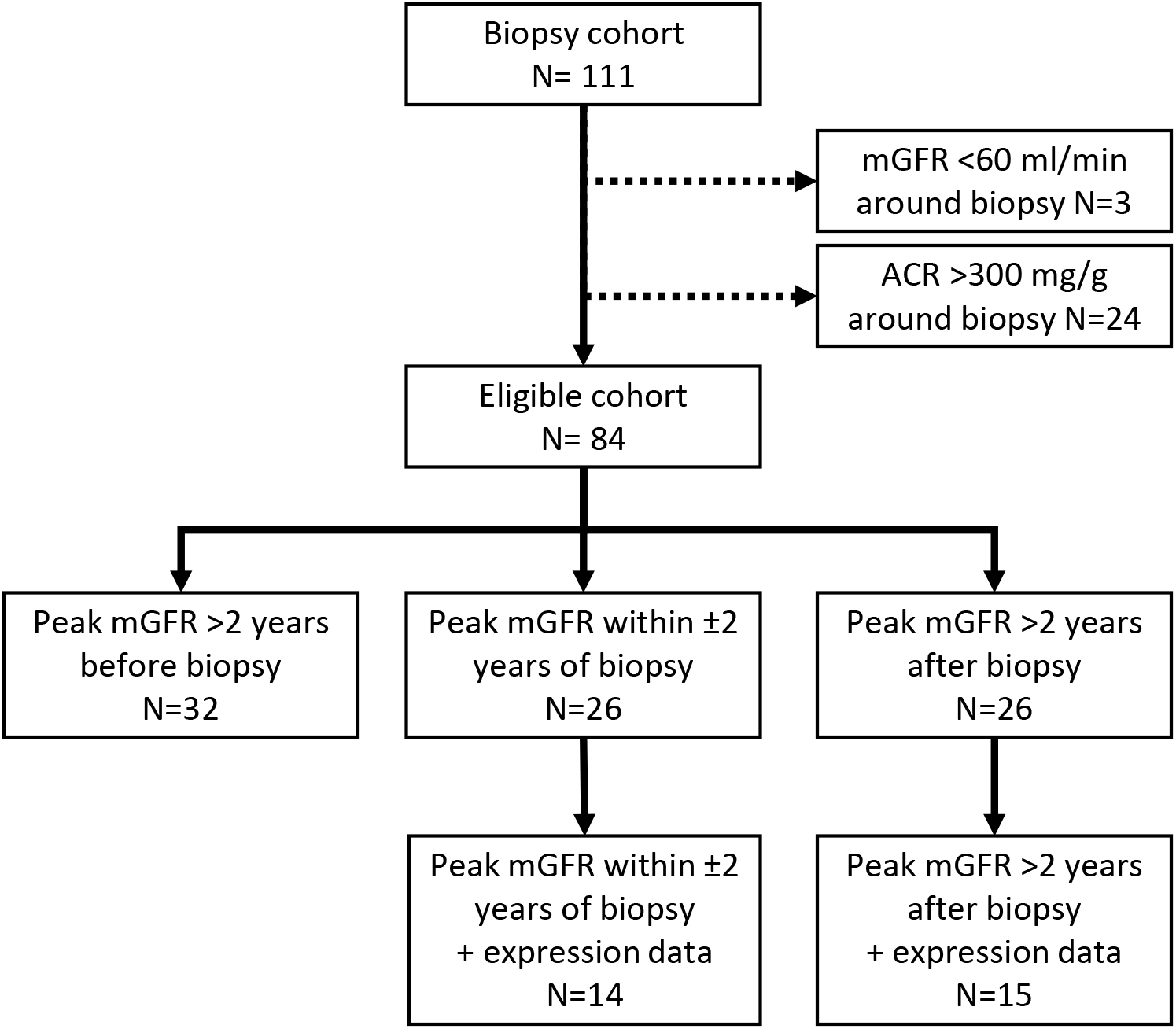
Flow chart describing participant selection. Participants, twenty-six each, in the HF group and pre-HF group were identified from those in the study cohort with a kidney biopsy (N=111), based on their peak GFR and in a subset with expression profiles.

### Laboratory measurements

Clinical laboratory procedures were described in detail previously^5, 15^. Briefly, participants were examined in the morning after an overnight fast. Urine creatinine was measured by a modified Jaffé reaction, and urinary albumin by nephelometric immunoassay. Any albumin concentrations below the detection threshold of 6.8 mg/L were set to 6.8 mg/L in the analyses.

GFR was measured by the urinary clearance of non-radioactive iothalamate^17^. Diuresis was initiated by an oral water load after the bladder was emptied. A loading dose of 300 mg iothalamate plus 3 mg/kg for each 1 kg >100 kg was given intravenously, followed by a continuous infusion to maintain a constant serum concentration. After equilibration, four serum and urine samples were collected at 20-minute intervals and were frozen at −80°C until the day of assay. Iothalamate concentrations in these samples were measured by high-performance liquid chromatography (Waters, Milford, MA).

### Quantitative morphometry

Structural parameters were measured by unbiased random sampling. Biopsy tissue was processed and embedded in epoxy resin (LX112; Ladd Research Industries, Williston, VT). Measurements were made from digital micrographs, and formal stereological methods were used to account for 2-dimensional sampling of 3-dimensional objects^15^. Kidney biopsy tissue was prepared for light and electron microscopy studies according to standard procedures^18-20^. The following glomerular structural parameters were measured on electron microscopy images as described in detail elsewhere^18, 19, 21^: glomerular basement membrane width^22, 23^, mesangial fractional volume, (including mesangial cell and mesangial matrix fractional volumes)^22, 23^, glomerular filtration surface density^22, 23^, foot process width^24^, percentage of endothelial fenestrations^25^, and the glomerular podocyte fractional volume per glomerulus^26^. Cortical interstitial fractional volume^20^ and mean glomerular volume^27, 28^ were estimated using light microscopy.

### Gene co-expression analyses

Tissue from the kidney biopsies underwent microdissection into glomerular and tubulointerstitial compartments. A GeneChip Human Genome series U133A and Plus 2.0 Array (Affymetrix, Santa Clara, CA) was used for gene expression profiling, and the resulting image files were obtained, processed, and normalized using quantile normalization in the Robust Multi Array Average method as implemented in the Affy package^29^. We used a Bayesian method, COMBAT, for correcting batch effects in the gene expression data^30^. Batch corrected log2 transformed data were used for all the downstream computational analyses.

Eigengene-based weighted gene co-expression network analysis modules were constructed from glomerular transcriptomes using Weighted Gene Co-expression Network Analysis (WGCNA)^31^. Pairwise Pearson correlations among all the genes in the expression matrix were computed. Modules (groups) of highly interconnected genes were constructed from a topological overlap matrix and hierarchical clustering. An eigen gene, a vector that summarizes the expression of all genes in that module akin to PC1, is constructed for all the modules. The modules whose eigen genes are highly correlated are merged. After the iterative process the gene expression matrix is reduced to co-expression modules. Eigen genes were then correlated (Pearson correlation) with the phenotypes of interest, the HF categories and the morphometric parameters. Transcripts contained in modules with statistically significant (p ≤0.05) associations to these traits were used for downstream functional analysis. All statistical analysis was done in R statistical software (www.r-project.org) and Stata MP 15 (Stata Corp., College Station, TX; www.stata.com). Ingenuity Pathway System (IPA) (Qiagen, Redwood City, CA, USA) was used to reveal statistically significantly associated functional pathways. Significance was set at a Bonferroni adjusted p-value of < 0.05.

### Pathway Analysis

Pathway analysis was performed using the IPA software querying the HR associated transcripts. Cytoscape^32^ visualization platform and ggplot2 package in R statistical platform was used to render the network images from the pathway network. An in-house custom python script was used to parse the IPA output for the Cytoscape subnetwork generations. Pathways with less than 5 genes shared with each other were filtered out to reduce the number of nodes in the pathway network. Major cancer pathways were also removed. The resulting network yielded 175 nodes and 5288 edges. To construct subnetworks, we used default parameters in the Cytoscape plugin, MCODE^33^.

### scRNAseq data generation and analysis

Single cell transcriptomes were generated with 2-3 mg of the biopsy core samples from CryoStor® (Stemcell Technologies) preserved DKD (N=44) and control (living donor, N=18) biopsies^34^. Tissue processing and single cell isolation were published previously^34, 35^. In brief, individual cell barcoding, reverse RNA transcription, library generation and single cell sequencing using Illumina were all performed using the 10X Genomics protocol13. The output from the sequencer was first processed by CellRanger (https://support.10xgenomics.com/single-cell-gene-expression/software/pipelines/latest/what-is-cell-ranger). Data analyses were performed on the CellRanger output data files using the Seurat 3 R package (https://cran.r-project.org/web/packages/Seurat/index.html)^34^.

### Cell-Cell Crosstalk Analysis

NicheNetR^36^ (https://github.com/saeyslab/nichenetr) was used to identify ligand-receptor (LR) interactions that drive the observed expression changes in the target cell population in the single cell transcriptome. NichNetR compiles literature-based ligand-receptor interactions, signal transductions and regulatory networks to prioritize the ligand-receptor-target gene identification. Based on cell type enrichment of HF genes, we focused on the cell-cell communication between the endothelial cells and mesangial cells to identify the LR interactions and downstream target genes.

## Results

### Participant characteristics

Summary clinical and demographic characteristics for the HF and pre-HF groups (**Table 1)** showed a mean GFR in the HF group of 173±48 ml/min, 21 ml/min higher, on average, than in the pre-HF group (152±38 ml/min, *p*=0.08). The HF group also had higher mean HbA1c (hemoglobin A1C, *p*=0.03) and median ACR (*p*=0.007). Though not statistically significant, there was also a trend towards longer duration of diabetes. Comparison of histopathological structural parameters from the kidney biopsies (**Table 2**) showed wider glomerular basement membrane (*p*=0.02) and higher mesangial fractional volume (*p*=0.004) among those in the HF group, reflecting greater structural changes near peak GFR, compared to pre-HF. Of note the difference in mesangial fractional volume was due predominantly to differences in the mesangial matrix fractional volume (p=0.0001) and not to differences in mesangial cell fractional volume (*p*=0.25). Podocyte density (*p*=0.03) was also slightly lower among those in the HF group, but podocyte fractional volume of the glomerulus was maintained, indicating that podocytes were fewer in number but larger. Clinical characteristics and structural parameters in the 32 participants not included in the primary analyses because their peak GFR occurred >2 years before the kidney biopsy compared with the 52 participants included in the study are provided in **Supplemental Table 1**. These patients had longer diabetes duration (*p*<0.001), lower GFR (*p*=0.007), and tissue morphometric measurements including increased cortical interstitial fractional volume and decreased glomerular podocyte fractional volume (*p*=0.032), reflective of their more advanced stage of DKD.

**Table 1.** Clinical characteristics at time of kidney biopsy by peak measured GFR group

| Characteristic | Pre-HF<br>(n=26) | HF<br>(n=26) | *p*-value |
| --- | --- | --- | --- |
| Male sex (%) | 7 (26.9) | 3 (11.5) | 0.16 |
| Age (years) | 44.8 ± 8.6 | 44.6 ± 12.0 | 0.95 |
| Diabetes duration (years) | 12.4 ± 3.4 | 14.4 ± 4.3 | 0.07 |
| BMI (kg/m<sup>2</sup>) | 37.0 ± 8.0 | 37.2 ± 9.2 | 0.91 |
| Systolic blood pressure (mmHg) | 120 ± 8 | 121 ± 8 | 0.70 |
| Diastolic blood pressure (mmHg) | 75 ± 5 | 77 ± 5 | 0.41 |
| HbA1c (%) | 8.7 ± 2.0 | 9.9 ± 1.8 | 0.03 |
| mGFR (ml/min) | 152 ± 38 | 173 ± 48 | 0.08 |
| ACR (mg/g) | 18 (13, 31) | 58 (34, 74) | 0.007 |
| RAS use (%) | 21 (80.8%) | 17 (65.4%) | 0.21 |

Values are means ± standard deviation, n and (frequency), or median (interquartile range).
Abbreviations: ACR = albumin-to-creatinine ratio, BMI = body mass index, HbA1c = hemoglobin A1c, mGFR = measured glomerular filtration rate; RAS = renin angiotensin system blockers.
*p*-value for male sex is from a chi-square test; *p*-values for continuous variables are based on *t*-tests; ACR was log transformed prior to analysis.

**Table 2.**
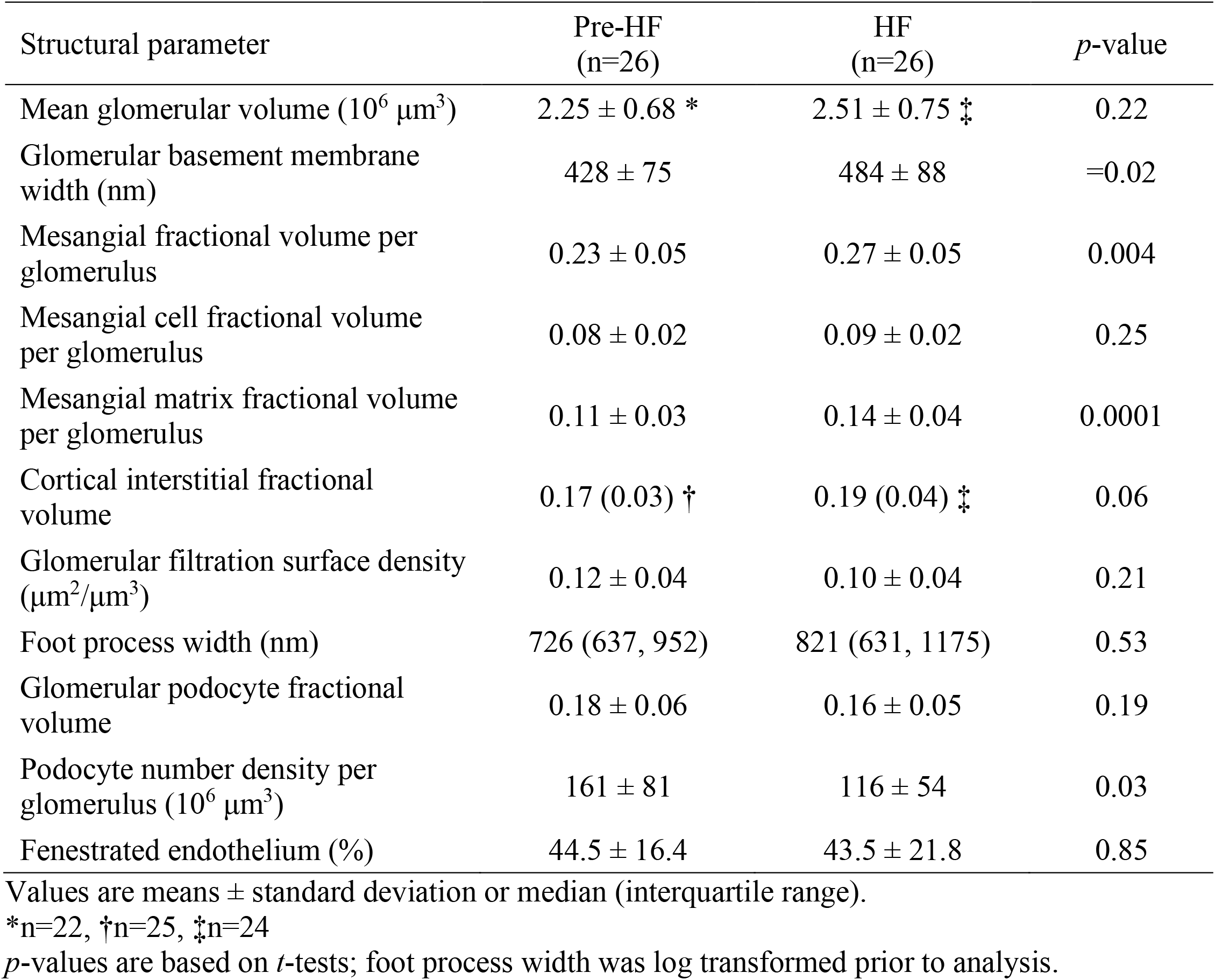
Morphometric measures by peak measured GFR group

| Structural parameter | Pre-HF<br>(n=26) | HF<br>(n=26) | *p*-value |
| --- | --- | --- | --- |
| Mean glomerular volume ($10^6 \mu\text{m}^3$) | $2.25 \pm 0.68$ * | $2.51 \pm 0.75$ ‡ | 0.22 |
| Glomerular basement membrane width (nm) | $428 \pm 75$ | $484 \pm 88$ | =0.02 |
| Mesangial fractional volume per glomerulus | $0.23 \pm 0.05$ | $0.27 \pm 0.05$ | 0.004 |
| Mesangial cell fractional volume per glomerulus | $0.08 \pm 0.02$ | $0.09 \pm 0.02$ | 0.25 |
| Mesangial matrix fractional volume per glomerulus | $0.11 \pm 0.03$ | $0.14 \pm 0.04$ | 0.0001 |
| Cortical interstitial fractional volume | $0.17 (0.03)$ † | $0.19 (0.04)$ ‡ | 0.06 |
| Glomerular filtration surface density ($\mu\text{m}^2/\mu\text{m}^3$) | $0.12 \pm 0.04$ | $0.10 \pm 0.04$ | 0.21 |
| Foot process width (nm) | 726 (637, 952) | 821 (631, 1175) | 0.53 |
| Glomerular podocyte fractional volume | $0.18 \pm 0.06$ | $0.16 \pm 0.05$ | 0.19 |
| Podocyte number density per glomerulus ($10^6 \mu\text{m}^3$) | $161 \pm 81$ | $116 \pm 54$ | 0.03 |
| Fenestrated endothelium (%) | $44.5 \pm 16.4$ | $43.5 \pm 21.8$ | 0.85 |

Values are means ± standard deviation or median (interquartile range).
\*n=22, †n=25, ‡n=24
*p*-values are based on *t*-tests; foot process width was log transformed prior to analysis.

To determine whether HF categorization based on measured GFR could be replicated using estimated GFR (eGFR), we repeated the analyses using the serum creatinine based CKD-EPI equation^16^ (**Supplemental Table 2**). eGFR trajectories resulted in significant misclassification with the vast majority (71/84; 85%) of study participants being classified as reaching peak eGFR >2 years before biopsy. Only 4 participants (5%) had a peak eGFR within 2 years of biopsy with the remaining 9 participants (11%) reaching peak eGFR >2 years after biopsy. Thus, eGFR is not a useful proxy for measured GFR in determining the timing of peak GFR. Accordingly, all gene expression analyses described below were based on measured GFR trajectories.

Twenty-nine of the 52 participants in the study cohort had sufficient tissue available from kidney biopsies for Affymetrix-based glomerular gene expression analyses (14 in the peak HF group and 15 in the HF after biopsy group). The clinical and morphometric characteristics of this subset with expression data were similar to those without expression data except that those with expression data were younger (*p*=0.005), had shorter diabetes duration (*p*=0.02), lower glomerular filtration surface density (*p*=0.02), and more fenestrated endothelium (*p*=0.005) (**Supplemental Table 3**). Just as in the full cohort, the HF group in the subset of participants with expression data had higher HbA1c (p=0.006) and ACR (p=0.05) than those in the pre-HF group. Unlike the full cohort there was significantly lower use of renin-angiotensin-system (RAS) inhibitors among the HF group compared to the pre-HF group (57.1% vs. 93.3%, p=0.04). Glomerular basement membrane width and mesangial fractional volume, again driven by mesangial matrix fractional volume, remained higher in the HF group, although only the difference in mesangial matrix fractional volume was statistically significant, and podocyte density per glomerulus was significantly lower. As with the full cohort, statistically significant differences in the other podocyte parameters were not observed in the subset of participants with expression data (**Supplemental Table 4**).

### Glomerular compartment gene expression associations and hyperfiltration gene modules

Gene expression profiles were first analyzed using data driven WGCNA resulting in 22 co-expression gene clusters (modules, **Figure 3A**). Three of these modules were significantly associated with HF. Of the 1240 differentially expressed genes represented by these modules, 95% were upregulated in the HF compared to pre-HF group (**Figure 3B, Supplemental Table 5**). These modules correlated (**Figure 3B**) positively with peak GFR at time of biopsy and negatively with mesangial cell volume, consistent with observed associations at the cohort level. However, these modules did not show strong association with GFR and ACR when analyzed as continuous variables.

**Figure 3.**
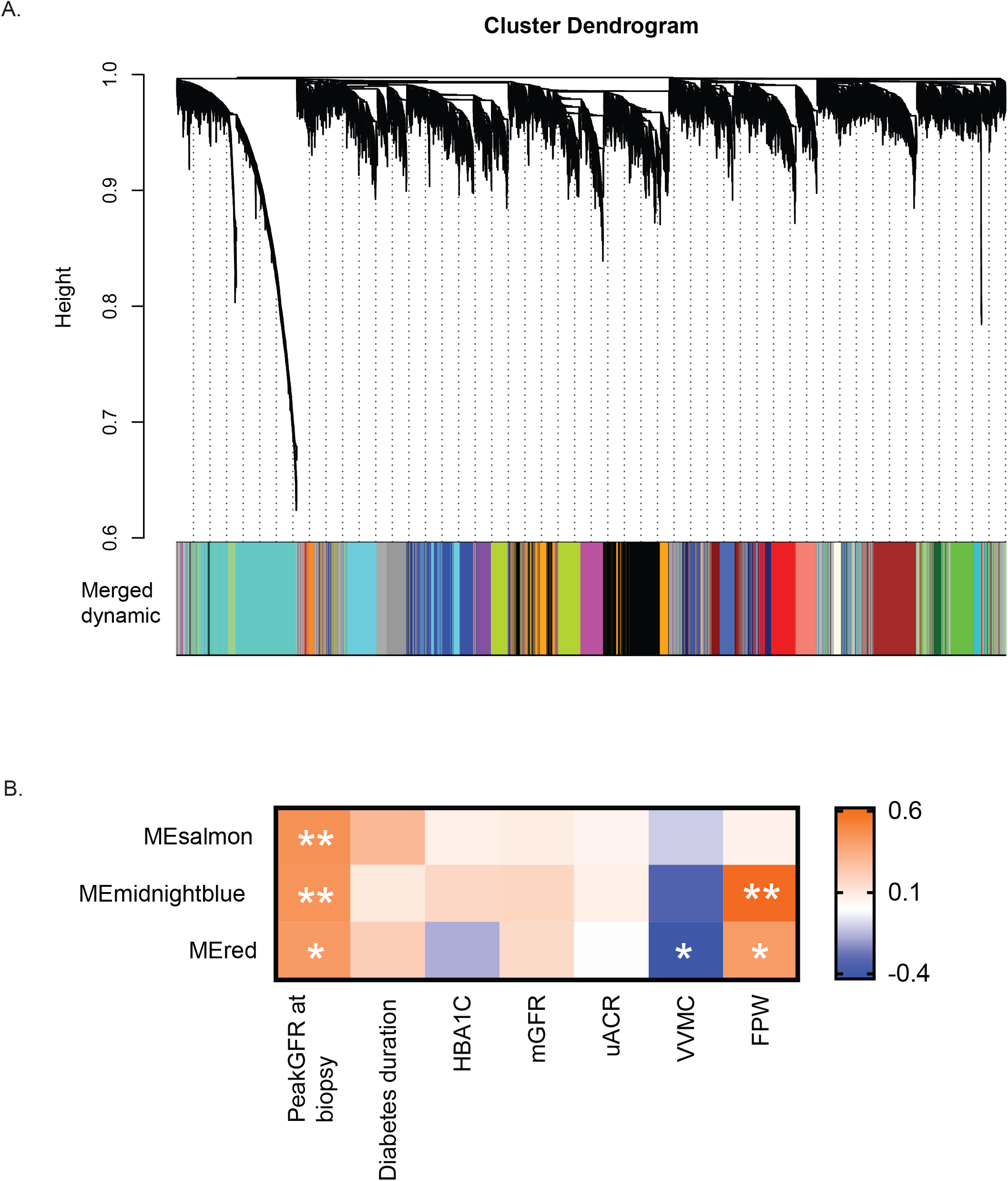
HF associated modules and their eigengenes’ association with duration of diabetic kidney disease. A. Cluster dendrogram of WGCNA based gene modules from glomerular transcription data from 29 participants. B. Three modules had ≥95% genes over-expressed in the HF group compared to pre-HF group. These modules were also significantly (*p<0.05, **p≤0.01) positively correlated with peak GFR at time of biopsy and negatively with mesangial cell volume (VVMC). Abbreviations: ME- modules; PeakGFRatBx – Peak GFR at kidney biopsy/HF group; HBA1C-Hemoglobin A1C; mGFR-measured glomerular filtration rate; uACR- urine albumin-to-creatinine ratio; VVMC - mesangial fractional volume per glomerulus; FPW- foot process width

### Hyperfiltration Pathways

Pathway analyses of the genes in these three modules yielded 194 significantly associated pathways (*p*≤0.05) (**Supplemental Table 6**). **Figure 4** lists the top 100 pathways associated with the HF genes. Pathways included in this list were selected based on the adjusted p-value and number of HF genes in each pathway and presented alphabetically. Among these key enriched pathways representing the hyperfiltration milieu were known growth factor signaling pathways such as platelet-derived growth factor (PDGF), vascular endothelial growth factor (VEGF) as well as endhothelin-1, TGFB1, integrin, angiogenesis and epithelial-mesenchymal transition pathways. The network maps of the top 30 interconnected HF genes (**Supplemental Figure 1**) as well as Cytoscape pathway analysis (**Supplemental Figure 2**) also identified many of the same key pathways.

**Figure 4.**
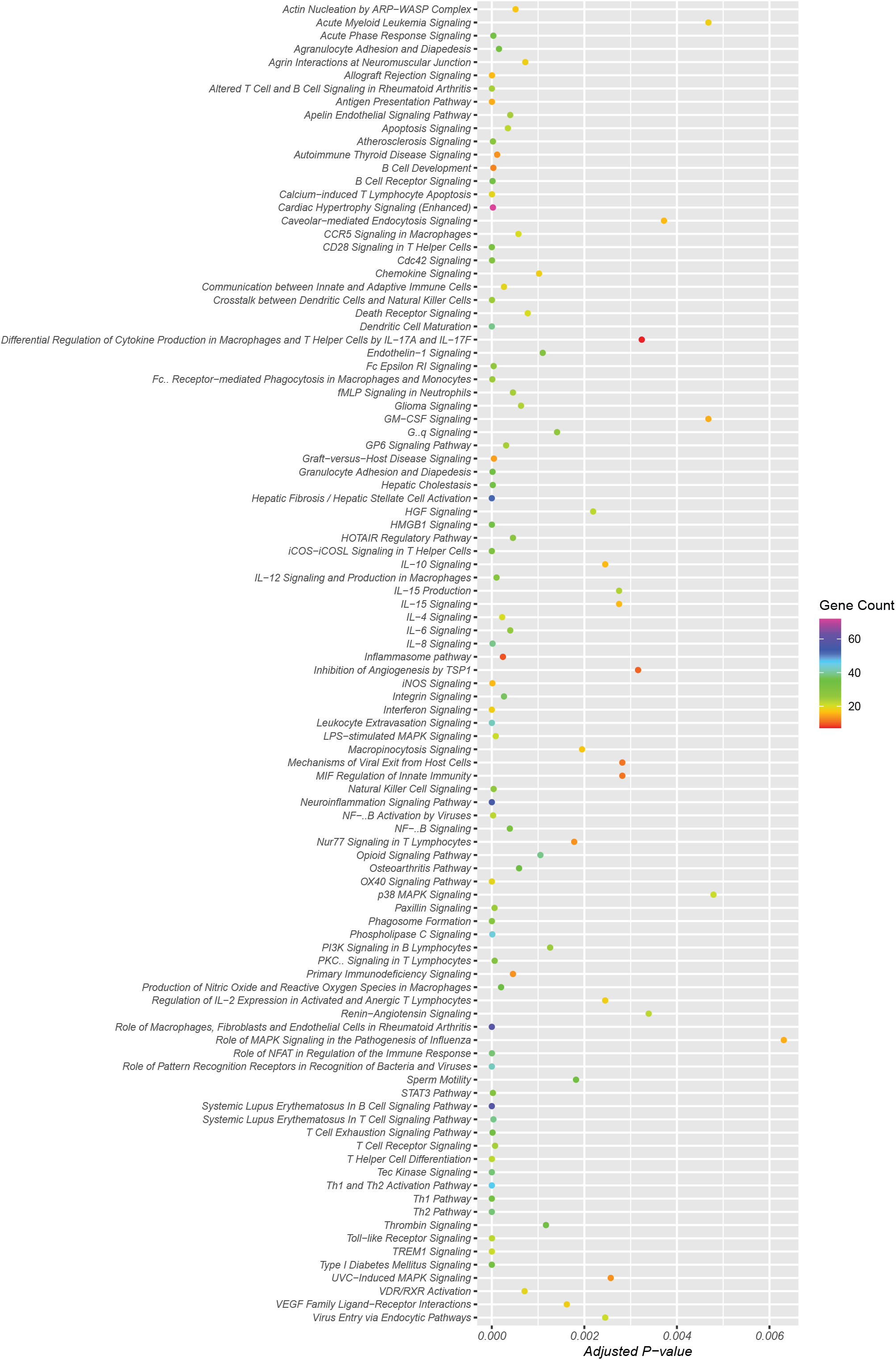
Top 100 significantly enriched pathways represented in the HF gene set arranged alphabetically. The top100 pathways encompassed by the HF genes were sorted based on adjusted p-values and plotted using the R package ggplot. The pathways in the figure are ordered alphabetically. The dots (Gene counts) are colored based on the number of pathway genes (purple >70; red <10) represented in the HF gene set and the x-axis describes membership of these genes in the corresponding pathway (adjusted p-values for all the genes in a dot)

### Kidney localization of hyperfiltration activation signals using scRNAseq analyses

To better elucidate the regulation of these pathways within the kidney, the cellular localization of active HF associated pathways were determined using scRNAseq data. Kidney biopsy samples from an independent group of 44 Pima Indians representing the same population pool with type 2 diabetes and 18 healthy controls were analyzed^34^. Clinical and morphometric measures for these groups are shown in **Supplemental Table 7**. Mean age of this single cell cohort of Pima Indians was 41.0±11.1 years, diabetes duration was 12.2±7.5 years, GFR was 159±58 ml/min, and median ACR was 18 (9, 48) mg/g. The 1240 genes from the 3 HF associated modules identified in the Pima cohort were mapped onto the 20 cell clusters identified from this cohort (**Figure 5A**). The highest enrichment (28%) was to the endothelial clusters indicating more than 350 of the HF genes were enriched(higher expression) in endothelial cells followed by fibroblasts and myeloid cells (**Figure 5B**). Further subclustering of the endothelial cell cluster and mapping of HF gene expression (**Figure 5C**) revealed a clear pattern of enriched activation of HF genes in intrinsic glomerular endothelial cells, where 30% of the HF genes showed maximal expression compared to all other cell clusters.

**Figure 5.**
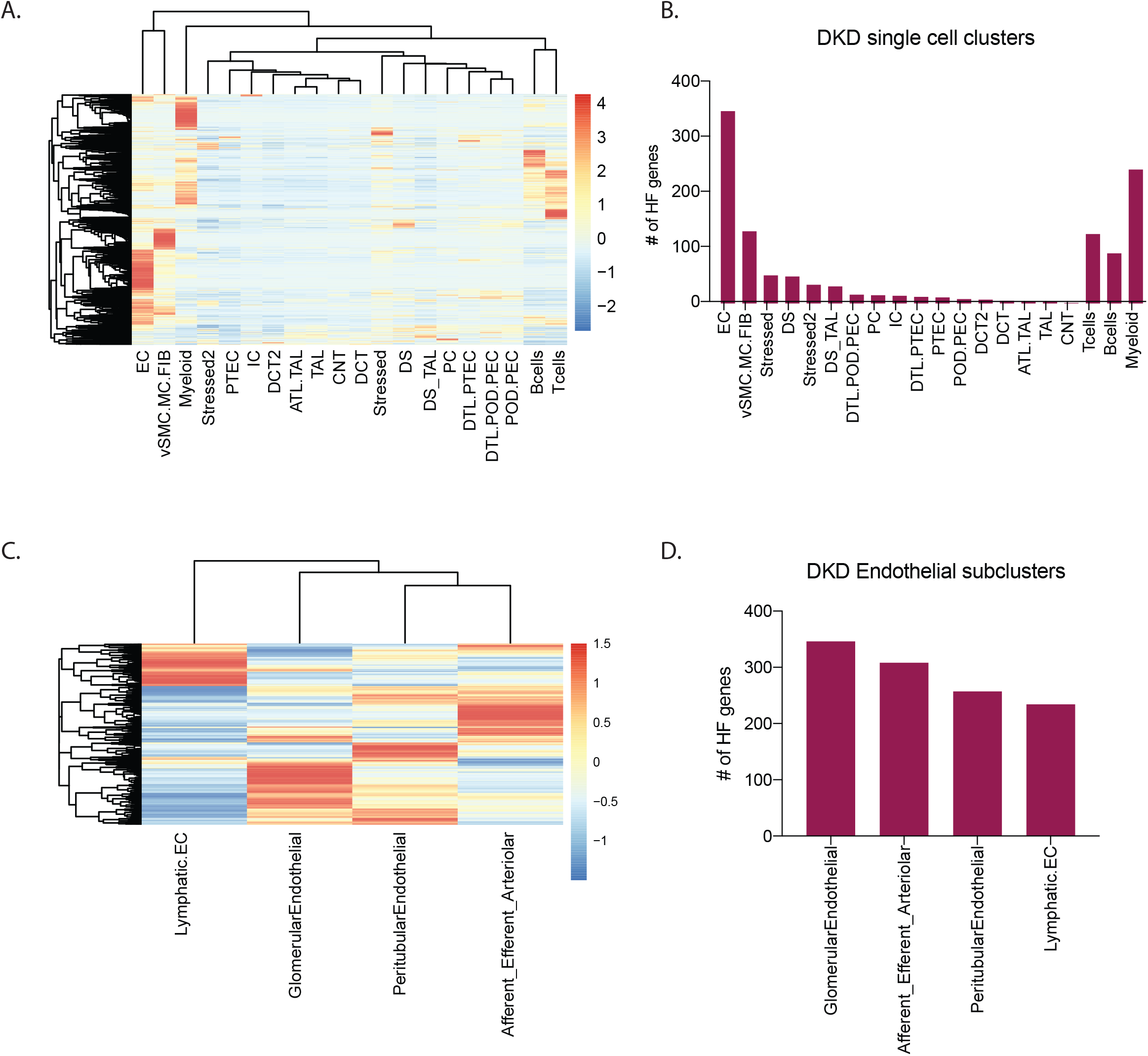
Single cell RNAseq (scRNAseq) analyses. A. Heatmap shows relative expression of HF genes in each of the cell type clusters derived from scRNAseq of kidney biopsies, with higher expression (red) of HF genes visualized in the endothelial cell (EC) cluster. B. Plot of the number of expressed HF genes in each scRNAseq cell type cluster also shows presence of more HF genes in the EC cluster. C. Heatmap shows expression profiles of HF genes and D. number of HF genes expressed in each EC subcluster. Number of genes in panels B and D were defined by their maximum average expression in each of the clusters. Each gene was assigned to only one cluster based on its maximum expression. Abbreviations: ATL-Ascending thin loop of Henle; CNT-Connecting tubule; DCT-Distal connecting tubule; DTL-Descending loop of Henle; EC-endothelial cells; IC-intercalated cells; MC-Mesangial Cell; PC-principal cells; PEC-Parietal epithelial cell: POD-Podocyte; PTEC-proximal tubular cells; TAL- thick ascending loop; vSMC-Vascular smooth muscle cells; Stressed/ Stressed 2-Clusters showing ribosomal/stress genes/ injury markers as top marker genes (Stressed cluster were akin to stressed proximal or descending loop of Henle cells whereas cells in the Stressed 2 cluster shared similarities with distal nephron cells.)

### Ligand-Receptor interactions during hyperfiltration

Pathway and single cell enrichment analysis of hyperfiltration associated genes all indicate activation of transcription pathways in endothelial cells (**Figure 5, Supplemental Table 6, Supplemental Figures 1 and 2**). This activation was further explored using a ligand-receptor-target gene network approach^36^. In addition to establishing the link between the ligand and receptors in the sender/receiver cells, NicheNetR also connects the ligand-receptor networks with upregulated downstream intracellular signaling target transcripts in the receiver cells as a surrogate for pathway activation. The top 15 ligand-receptor pairs based on the NicheNetR predicted ligand activity in the endothelial cell cluster and corresponding receptor activity in mesangial cells. Top ligands in endothelial cells include endothelin-1 (EDN1), transforming growth factor β1 (TGFB1), vascular endothelial growth factor A (VEGFA), C-X-C motif chemokine ligand 10 (CXCL10). The corresponding receptors in mesangial cells include endothelin receptors A and B (EDNRA/B), transforming growth factor β receptor 2/ Erb-B2 receptor tyrosine kinase 2 (TGFBR2/ERBB2) and protein tyrosine phosphatase receptor type B/ neuropilin 1/ platelet derived growth factor receptor β (PTPRB/NRP1/PDGFRB) in the mesangial cells. **Figure 6A** shows the relative enrichment of these ligands in endothelial cells versus other kidney cell types, while **Figure 6B** shows an increase in relative expression of ligands in endothelial cells in DKD versus living donor (LD) controls^34^. By mapping the main ligands from endothelial cells to the corresponding receptors in mesangial cells, we were then able to identify primary activation cascades in the endothelial-mesangial crosstalk during HF (**Figure 6C and Supplemental Figure 3**). These networks were enriched for genes in the signaling pathways of Rho GTPase small molecules, axon guidance, actin skeleton, VEGFA-NRP1 induced angiogenesis and the endothelin-1 pathway.

**Figure 6.**
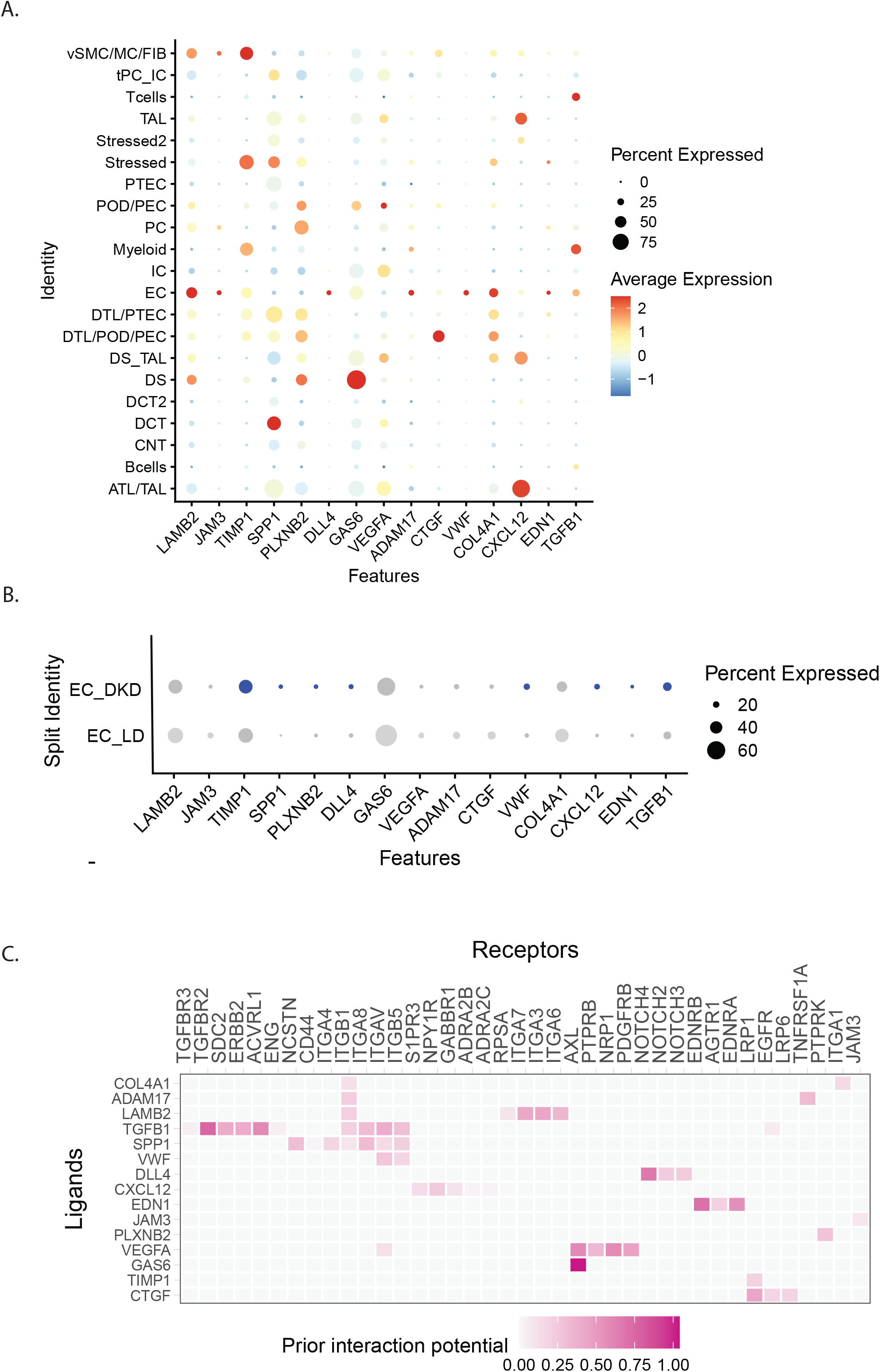
NicheNet Analysis of ligand receptor crosstalk. NicheNetR (https://github.com/saeyslab/nichenetr) was used to identify ligand-receptor (LR) interactions that drive the observed expression changes in the single cell transcriptome. A. Expression of the top 15 nichenetR predicted ligands were compared across cell types. B. Several of these ligands are upregulated in DKD endothelial cells (EC_DKD) compared to endothelial cell controls (EC_LD). The dot size represents the percentage of cells expressing the gene in the respective clusters and the color represent the intensity of the expression level from grey(low) to blue (high). C.Predicted ligand-receptor interaction for the top 15 ligands in the endothelial and mesangial cell. The interaction pairs are prioritized based on the weights derived from the ligand-receptor network from nichenet data sources. Abbreviations: ATL-Ascending thin loop of Henle; CNT-Connecting tubule; DCT-Distal connecting tubule; DTL- Descending loop of Henle; EC-endothelial cells; IC- intercalated cells; MC-Mesangial Cell; PC- principal cells; PEC- Parietal epithelial cell: POD- Podocyte; PTEC-proximal tubular cells; TAL- thick ascending loop; vSMC-Vascular smooth muscle cells; Stressed/ Stressed 2-Clusters showing ribosomal/stress genes/ injury markers as top marker genes (Stressed cluster were akin to stressed proximal or descending loop of Henle cells whereas cells in the Stressed 2 cluster shared similarities with distal nephron cells.)

## Discussion

To our knowledge, this study is the first to report differences in gene expression associated with HF in humans and to link these differences to contemporaneous glomerular structural lesions characteristic of HF^8, 14, 37^. These associations were identified through the successful use of an individual’s peak measured GFR to classify HF rather than an arbitrary absolute HF threshold that fails to account for individual variations in nephron numbers ^14^ and in the capacity of nephrons to handle prolonged states of HF. Gene expression differences between the HF groups as defined in this study suggest mechanisms uniquely associated with or contributing to the zenith of GFR in individuals with type 2 diabetes as potential therapeutic targets. Mapping the differentially regulated HF genes onto specific kidney cell populations allowed us to identify key cellular mediators of HF and demonstrate that HF genes are significantly enriched in endothelial cells, macrophages, and fibroblasts relative to other kidney cell types. These observations provide unbiased evidence of endothelial stress in the core regulation unit of glomerular hemodynamics.

Integration of kidney tissue level and single cell transcriptional data revealed the main ligands EDN1, TGFB1, VEGFA, growth arrest-specific gene 6 (Gas6) and delta like canonical notch ligand 4 (DLL4) were primarily produced by endothelial cells in the HF state and enabled identification of predicted downstream cellular targets with corresponding over-expressed receptors in the mesangial cells using NicheNet^36, 38, 39^ (**Figure 7**). Hyperglycemic conditions, AngII, and reactive oxygen species all activate TGFB1 signaling which then leads to inflammation and fibrosis in progressive DKD^40^. The receptors included multiple integrins in the cell matrix signaling mechanisms. Mesangial cells targeted by this receptor-ligand interaction showed enrichment of genes in the signaling pathways of Rho GTPase small molecules, axon guidance, actin skeleton, VEGFA-NRP1 induced angiogenesis, and the endothelin-1 pathway. Identification of the endothlin-1^41^ and TGFB1 pathways in our study is supported by prior literature on the role of these pathways in DKD susceptibility^42-53^. The Gas6-AKT pathway was shown to be involved in mesangial hypertrophy in rat models of DKD^54, 55^. DLL4 induced Notch signaling influences nephron number and segmentation during kidney development but in DKD, has been shown to promote glomerulopathy, tubulointerstitial fibrosis and possibly arteriopathy and inflammation, likely through VEGFA mediated signalling^56-59^. Several of these pathways have been identified and effectively targeted in established DKD^43^. Our results extend the activation of these pathways into the earliest stages of DKD, arguing for potential beneficial effects of these treatment modalities over the entire course of DKD. Further, early detection of maximum GFR could provide an opportunity for mitigating loss of kidney function. For example, treatment with sodium-glucose co-transporter 2 (SGLT2) inhibitors in early DKD reduced GFR while reducing serum glucose levels, likely through a reduction in hyperfiltration^11^, among other mechanisms.

**Figure 7.**
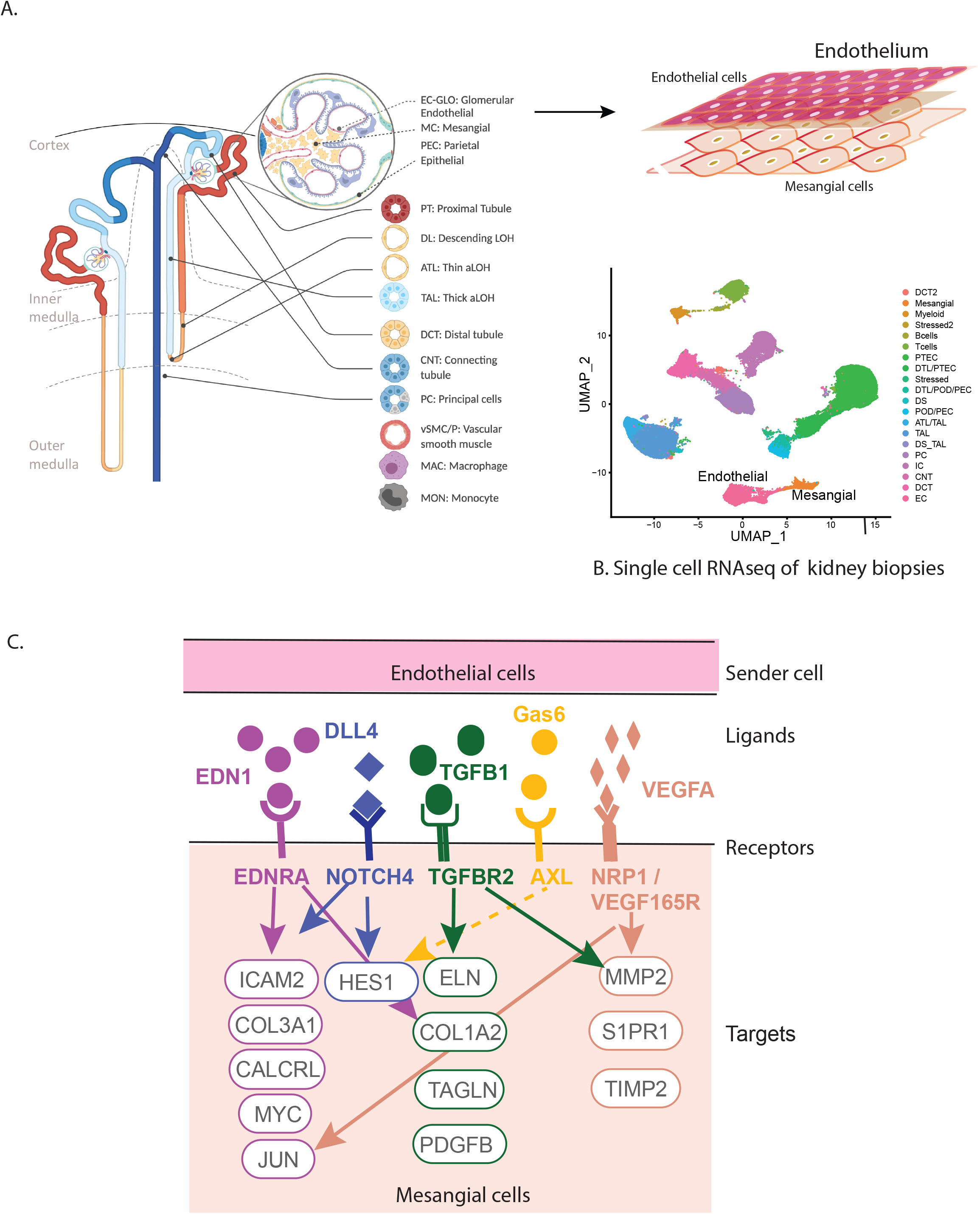
Endothelial signaling activates intracellular targets in mesangial cells. HF genes in the kidney are involved in the crosstalk between endothelial and mesangial cells in the glomerulus (A). The transcriptional profiles of these cell types in DKD could be distinguished using scRNAseq (B) leading to the identification of intracellular targets in mesangial cells (C) providing molecular insights into hyperfiltration associated with early DKD.

The presence of an inflammatory signal derived from an intraglomerular macrophage cell population indicates that HF is linked to immigrating immune cells in the first steps of glomerular volume expansion^60-63^. Indeed, increased macrophage infiltration in glomeruli and increased expression of several genes related to inflammation, including MCP-1, were observed in hyperfiltering mice with 5/6 nephrectomy^64^. Furthermore, increased excretion of cytokines in the urine is reported in persons with diabetes during hyperfiltration defined as mGFR >135 ml/min/1.73m^2 60^.

Nitric oxide signaling pathways and tumor necrosis factor-alpha receptor 2 (TNFR2) signaling pathways which have been associated with hyperfiltration in rats^10^, have been implicated in decline in eGFR, incident CKD and ESRD in diabetes^65-67^. Indeed, associations were found with early glomerular lesions and ESRD in a Pima Indian cohort^68, 69^. However, a study of the non-diabetic general population using measured GFR found an opposite relationship between TNFR2 and GFR decline, perhaps indicating a disease-specific role of these pathways in diabetes, or alternatively an association of TNFR2 with early hyperfiltration (longitudinal increase in GFR)^70^. More recently, the mineralocorticoid receptor antagonist finerenone was found to reduce CKD progression (eGFR decline) and cardiovascular events in patients with diabetes and CKD^63^. Mineralocorticoid receptor blocking counters several overexpressed pathways in the HF group in the present study, including the profibrotic TGF-β signaling, the vasoconstrictive endothelin-1 signaling and proinflammatory IL-6 signaling. Meanwhile, the GLP-1 receptor antagonist, Exedin-4, has been shown to ameliorate glomerular hyperfiltration, glomerular hypertrophy and albuminuria in rats, likely through its anti-inflammatory action^71^.

The main strengths of this study are the repeated GFR measurements using iothalamate clearance providing GFR trajectories for each participant through a substantial portion of their adult life. The GFR measurements preceded and followed research kidney biopsies in the same individuals, allowing an approach where the structural and gene expression findings could be placed in the context of each individual’s historical GFR trajectory. Some participants may have already been past their active HF stage before their GFR measurements began or their kidney biopsy was performed, which is why we excluded participants who had a low GFR and/or a high ACR at the time of biopsy, and those who had their measured GFR peak more than 2 years prior to their biopsy. To our knowledge, no other study population has combined frequently repeated GFR measurements in early diabetes with a kidney biopsy when participants could still plausibly be in the HF stage.

Some of the study’s strengths are also its limitations, in that replicating the study findings in other populations has not been possible as there are no other studies collecting these comprehensive invasive measures across decades. The rates and severity of obesity and DKD in Pima Indians are higher at a younger age than in most Caucasian or ethnically diverse research populations, and they also have a lower rate of concurrent cardiovascular disease. However, this does not necessarily mean the mechanisms of HF in this population are different from those in other populations, and previous mechanistic insights from gene expression findings in this population have indeed been replicated in unrelated populations^29^. Other limitations included the study population being reduced from 52 to 29 for gene expression analyses due to unavailability of tissue for these studies. This might have adversely affected statistical power. However, the results from analyses were significant through WGCNA based dimensionality reduction. The scRNAseq data were generated from tissue samples provided by a distinct set of study participants who did not undergo long-term serial GFR measurements, but they were part of the same population. Finally, we do not know the extent to which the use of RAS inhibitors altered the GFR trajectories among the study participants.

In conclusion, the integration of long-term GFR trajectories with morphometric and molecular analyses of research kidney biopsies enabled the identification of an endothelial stress response with concomitant activation of downstream mesangial cell pathways at peak HF. As these changes occur very early in the course of DKD, they may present an opportunity to prevent the serious complications that may follow. Taken together, this study, provides the molecular link between HF in humans and molecular pathways in the kidneys that may lead to structural injury and progressive GFR decline, providing a framework to map existing and new therapeutic targets for HF in DKD.

## Data Availability

Due to ethical considerations including individual privacy protection concerns, the board stipulated that individual-level genotype and gene expression data from this study cannot be made publicly available.

## Author contributions

The study was designed by RGN, MK, BOE; RGN, HCL enrolled participants and collected data; RM, FE, DF, VN developed analyses protocols and processed samples for bulk RNA and single cell analysis; VTNS, VN, T.M, BOE, HCL contributed to the data analysis; RGN, VTNS, TM, BOE, LM, HCL, MK, JLH, JH provided clinical input; scientific input for interpretation of results was provided by RGN, VTNS, VN, TM, BOE, HCL, PJN, MK and JH; VTNS, VN, LS, MK, RGN, HCL wrote and compiled the manuscript. All authors approved the final version.

## Study approval

All studies of the DKD population were approved by the Institutional Review Board of the National Institute of Diabetes and Digestive and Kidney Diseases.

## Acknowledgements

We thank Ms. Lois Jones, RN, Mr. Enrique Diaz, RN, Ms. Bernadine Waseta, and Ms. Camille Waseta for performing the studies in the diabetes cohort.

This work was performed in partial fulfillment of the requirements for the doctoral work of V.N. from the Medical Faculty of Ludwig-Maximilians-University Munich, Germany.

This work was supported in part by the Intramural Research Program at the National Institute of Diabetes and Digestive and Kidney Diseases (DK069062) to HCL and RGN, the American Diabetes Association (Clinical Science Award 1-08-CR-42) to RGN, and (DK083912, DK082841, DK020572, DK092926) to RGK, by the extramural research program of the National Institute of Diabetes and Digestive and Kidney Diseases R24 DK082841 and P30 DK081943 University of Michigan O’Brien Kidney Translational Core Center grants to MK, by JDRF 5-COE-2019-861-S-B ‘JDRF and M-Diabetes Center of Excellence’ at the University of Michigan to MK. The content is solely the responsibility of the authors and does not necessarily represent the official views of the National Institutes of Health.

## Supplementary Materials

### Supplemental Tables

Supplemental Table 1: Clinical and morphometric measurements comparing the participants included in the study (HF and pre-HF) and those who were excluded because their peak GFR occurred >2 years before their kidney biopsy

Supplemental Table 2: Timing of peak measured GFR compared to timing of peak eGFR in relation to kidney biopsy

Supplemental Table 3: Clinical and morphometric measures at the time of biopsy for the subset of participants with gene expression data compared to those without expression data

Supplemental Table 4: Clinical and morphometric measures at the time of biopsy by peak measured GFR group for the subset of participants with gene expression data

Supplemental Table 5: Complete list of genes in the three WGCNA modules where 95% genes (1240 genes) were upregulated in the HF compared to pre-HF group

Supplemental Table 6: List of 194 significantly associated pathways (p<0.05) from genes in the three HF modules

Supplemental Table 7: Clinical and morphometric measures for the participants who contributed single-cell expression data from kidney biopsies

### Supplemental Figures

Supplemental Figure 1: Top 30 interconnected genes in HF associated modules

Supplemental Figure 2: Hyperfiltration gene networks and pathways using cytoscape visualization

Supplemental Figure 3. NicheNet predicted intracellular targets of endothelial cell activated ligands in crosstalk with mesangial cells

## References

1. System USRD. 2020 USRDS Annual Data Report: Epidemiology of kidney disease in the United States. National Institutes of Health, National Institute of Diabetes and Digestive and Kidney Diseases, Bethesda, MD.

2. Ng M, Fleming T, Robinson M, et al. Global, regional, and national prevalence of overweight and obesity in children and adults during 1980-2013: a systematic analysis for the Global Burden of Disease Study 2013. Lancet 2014; 384: 766–781.

3. Brenner BM, Hostetter TH, Olson JL, et al. The role of glomerular hyperfiltration in the initiation and progression of diabetic nephropathy. Acta Endocrinol Suppl (Copenh) 1981; 242: 7–10.

4. Ruggenenti P, Porrini EL, Gaspari F, et al. Glomerular hyperfiltration and renal disease progression in type 2 diabetes. Diabetes Care 2012; 35: 2061–2068.

5. Nelson RG, Bennett PH, Beck GJ, et al. Development and progression of renal disease in Pima Indians with non-insulin-dependent diabetes mellitus. Diabetic Renal Disease Study Group. N Engl J Med 1996; 335: 1636–1642.

6. Park M, Yoon E, Lim YH, et al. Renal hyperfiltration as a novel marker of all-cause mortality. J Am Soc Nephrol 2015; 26: 1426–1433.

7. Helal I, Fick-Brosnahan GM, Reed-Gitomer B, et al. Glomerular hyperfiltration: definitions, mechanisms and clinical implications. Nat Rev Nephrol 2012; 8: 293–300.

8. D’Agati VD, Chagnac A, de Vries AP, et al. Obesity-related glomerulopathy: clinical and pathologic characteristics and pathogenesis. Nat Rev Nephrol 2016; 12: 453–471.

9. Lewko B, Stepinski J. Hyperglycemia and mechanical stress: targeting the renal podocyte. J Cell Physiol 2009; 221: 288–295.

10. Veelken R, Hilgers KF, Hartner A, et al. Nitric oxide synthase isoforms and glomerular hyperfiltration in early diabetic nephropathy. J Am Soc Nephrol 2000; 11: 71–79.

11. Fioretto P, Zambon A, Rossato M, et al. SGLT2 Inhibitors and the Diabetic Kidney. Diabetes Care 2016; 39 Suppl 2: oS165–171.

12. Kriz W, Lemley KV. A potential role for mechanical forces in the detachment of podocytes and the progression of CKD. J Am Soc Nephrol 2015; 26: 258–269.

13. Nelson RG, Tan M, Beck GJ, et al. Changing glomerular filtration with progression from impaired glucose tolerance to Type II diabetes mellitus. Diabetologia 1999; 42: 90–93.

14. Denic A, Mathew J, Lerman LO, et al. Single-Nephron Glomerular Filtration Rate in Healthy Adults. N Engl J Med 2017; 376: 2349–2357.

15. Weil EJ, Fufaa G, Jones LI, et al. Effect of losartan on prevention and progression of early diabetic nephropathy in American Indians with type 2 diabetes. Diabetes 2013; 62: 3224–3231.

16. MacIsaac RJ, Ekinci EI, Premaratne E, et al. The Chronic Kidney Disease-Epidemiology Collaboration (CKD-EPI) equation does not improve the underestimation of Glomerular Filtration Rate (GFR) in people with diabetes and preserved renal function. BMC Nephrol 2015; 16: 198.

17. Myers BD, Nelson RG, Tan M, et al. Progression of overt nephropathy in non-insulin-dependent diabetes. Kidney Int 1995; 47: 1781–1789.

18. Fioretto P, Kim Y, Mauer M. Diabetic nephropathy as a model of reversibility of established renal lesions. Curr Opin Nephrol Hypertens 1998; 7: 489–494.

19. Mauer M, Zinman B, Gardiner R, et al. Renal and retinal effects of enalapril and losartan in type 1 diabetes. N Engl J Med 2009; 361: 40–51.

20. Ibrahim HN, Jackson S, Connaire J, et al. Angiotensin II blockade in kidney transplant recipients. J Am Soc Nephrol 2013; 24: 320–327.

21. Mauer M, Caramori ML, Fioretto P, et al. Glomerular structural-functional relationship models of diabetic nephropathy are robust in type 1 diabetic patients. Nephrol Dial Transplant 2015; 30: 918–923.

22. Caramori ML, Kim Y, Huang C, et al. Cellular basis of diabetic nephropathy: 1. Study design and renal structural-functional relationships in patients with long-standing type 1 diabetes. Diabetes 2002; 51: 506–513.

23. Klein R, Zinman B, Gardiner R, et al. The relationship of diabetic retinopathy to preclinical diabetic glomerulopathy lesions in type 1 diabetic patients: the Renin-Angiotensin System Study. Diabetes 2005; 54: 527–533.

24. Toyoda M, Najafian B, Kim Y, et al. Podocyte detachment and reduced glomerular capillary endothelial fenestration in human type 1 diabetic nephropathy. Diabetes 2007; 56: 2155–2160.

25. Najafian B, Mauer M. Quantitating glomerular endothelial fenestration: an unbiased stereological approach. Am J Nephrol 2011; 33 Suppl 1: 34–39.

26. Najafian B, Tondel C, Svarstad E, et al. One Year of Enzyme Replacement Therapy Reduces Globotriaosylceramide Inclusions in Podocytes in Male Adult Patients with Fabry Disease. PLoS One 2016; 11: e0152812.

27. Weibel E. Stereological Methods. Practical methods for biological morphometry. Academic Press: London, 1979.

28. Lane PH, Steffes MW, Mauer SM. Estimation of glomerular volume: a comparison of four methods. Kidney Int 1992; 41: 1085–1089.

29. Nair V, Komorowsky CV, Weil EJ, et al. A molecular morphometric approach to diabetic kidney disease can link structure to function and outcome. Kidney Int 2018; 93: 439–449.

30. Johnson WE, Li C, Rabinovic A. Adjusting batch effects in microarray expression data using empirical Bayes methods. Biostatistics 2007; 8: 118–127.

31. Langfelder P, Horvath S. WGCNA: an R package for weighted correlation network analysis. BMC bioinformatics 2008; 9: 559.

32. Shannon P, Markiel A, Ozier O, et al. Cytoscape: a software environment for integrated models of biomolecular interaction networks. Genome Res 2003; 13: 2498–2504.

33. Bader GD, Hogue CWV. An automated method for finding molecular complexes in large protein interaction networks. BMC Bioinformatics 2003; 4: 2.

34. Menon R, Otto EA, Sealfon R, et al. SARS-CoV-2 receptor networks in diabetic and COVID-19-associated kidney disease. Kidney international 2020; 98: 1502–1518.

35. Menon R, Otto EA, Hoover P, et al. Single cell transcriptomics identifies focal segmental glomerulosclerosis remission endothelial biomarker. JCI insight 2020; 5.

36. Browaeys R, Saelens W, Saeys Y. NicheNet: modeling intercellular communication by linking ligands to target genes. Nat Methods 2020; 17: 159–162.

37. Elsherbiny HE, Alexander MP, Kremers WK, et al. Nephron hypertrophy and glomerulosclerosis and their association with kidney function and risk factors among living kidney donors. Clin J Am Soc Nephrol 2014; 9: 1892–1902.

38. Gladka MM. Cellular communication in a ‘virtual lab’: going beyond the classical ligand-receptor interaction. Cardiovasc Res 2020; 116: e67–e69.

39. Matsui I, Matsumoto A, Inoue K, et al. Single cell RNA sequencing uncovers cellular developmental sequences and novel potential intercellular communications in embryonic kidney. Sci Rep 2021; 11: 73.

40. Zhao L, Zou Y, Liu F. Transforming Growth Factor-Beta1 in Diabetic Kidney Disease. Front Cell Dev Biol 2020; 8: 187–187.

41. Dagamajalu SA-O, Rex DA-O, Gopalakrishnan L, et al. A network map of endothelin mediated signaling pathway. LID - 10.1007/s12079-020-00581-4 [doi] FAU - Dagamajalu, Shobha.

42. Barton M, Yanagisawa M. Endothelin: 30 Years From Discovery to Therapy. Hypertension 2019; 74: 1232–1265.

43. Cherney DZI, Bakris GL. Novel therapies for diabetic kidney disease. Kidney Int Suppl (2011) 2018; 8: 18–25.

44. Fu J, Lee K, Chuang PY, et al. Glomerular endothelial cell injury and cross talk in diabetic kidney disease. Am J Physiol Renal Physiol 2015; 308: F287–297.

45. Kohan DE, Pollock DM. Endothelin antagonists for diabetic and non-diabetic chronic kidney disease. Br J Clin Pharmacol 2013; 76: 573–579.

46. Li YH, Sheu WH, Lee WJ, et al. Synergistic effect of renalase and chronic kidney disease on endothelin-1 in patients with coronary artery disease - a cross-sectional study. Sci Rep 2018; 8: 7378.

47. Qi H, Casalena G, Shi S, et al. Glomerular Endothelial Mitochondrial Dysfunction Is Essential and Characteristic of Diabetic Kidney Disease Susceptibility. Diabetes 2017; 66: 763.

48. Zanatta CM, Veronese FV, Loreto Mda S, et al. Endothelin-1 and endothelin a receptor immunoreactivity is increased in patients with diabetic nephropathy. Ren Fail 2012; 34: 308–315.

49. Zhang L, Chen L, Gao C, et al. Loss of Histone H3 K79 Methyltransferase Dot1l Facilitates Kidney Fibrosis by Upregulating Endothelin 1 through Histone Deacetylase 2. J Am Soc Nephrol 2020; 31: 337–349.

50. Hong Q, Zhang L, Fu J, et al. LRG1 Promotes Diabetic Kidney Disease Progression by Enhancing TGF-beta-Induced Angiogenesis. J Am Soc Nephrol 2019; 30: 546–562.

51. Jung KY, Chen K, Kretzler M, et al. TGF-beta1 regulates the PINCH-1-integrin-linked kinase-alpha-parvin complex in glomerular cells. J Am Soc Nephrol 2007; 18: 66–73.

52. Loeffler I, Hopfer U, Koczan D, et al. Type VIII Collagen Modulates TGF-β1-induced Proliferation of Mesangial Cells. J Am Soc Nephrol 2011; 22: 649.

53. Mariani LH, Martini S, Barisoni L, et al. Interstitial fibrosis scored on whole-slide digital imaging of kidney biopsies is a predictor of outcome in proteinuric glomerulopathies. Nephrol Dial Transplant 2018; 33: 310–318.

54. Arai H, Nagai K, Doi T. Role of growth arrest-specific gene 6 in diabetic nephropathy. Vitam Horm 2008; 78: 375–392.

55. Nagai K, Matsubara T, Mima A, et al. Gas6 induces Akt/mTOR-mediated mesangial hypertrophy in diabetic nephropathy. Kidney Int 2005; 68: 552–561.

56. Bonegio R, Susztak K. Notch signaling in diabetic nephropathy. Exp Cell Res 2012; 318: 986–992.

57. Kretzler M, Allred L. Notch inhibition reverses kidney failure. Nat Med 2008; 14: 246–247.

58. Lin CL, Wang FS, Hsu YC, et al. Modulation of notch-1 signaling alleviates vascular endothelial growth factor-mediated diabetic nephropathy. Diabetes 2010; 59: 1915–1925.

59. Walsh DW, Roxburgh SA, McGettigan P, et al. Co-regulation of Gremlin and Notch signalling in diabetic nephropathy. Biochim Biophys Acta 2008; 1782: 10–21.

60. Har R, Scholey JW, Daneman D, et al. The effect of renal hyperfiltration on urinary inflammatory cytokines/chemokines in patients with uncomplicated type 1 diabetes mellitus. Diabetologia 2013; 56: 1166–1173.

61. Tonneijck L, Muskiet MHA, Smits MM, et al. Glomerular Hyperfiltration in Diabetes: Mechanisms, Clinical Significance, and Treatment. J Am Soc Nephrol 2017; 28: 1023–1039.

62. Heerspink HJL, Perco P, Mulder S, et al. Canagliflozin reduces inflammation and fibrosis biomarkers: a potential mechanism of action for beneficial effects of SGLT2 inhibitors in diabetic kidney disease. Diabetologia 2019; 62: 1154–1166.

63. Bakris GL, Agarwal R, Anker SD, et al. Effect of Finerenone on Chronic Kidney Disease Outcomes in Type 2 Diabetes. N Engl J Med 2020.

64. Sasaki M, Shikata K, Okada S, et al. The macrophage is a key factor in renal injuries caused by glomerular hyperfiltration. Acta Med Okayama 2011; 65: 81–89.

65. Gohda T, Niewczas MA, Ficociello LH, et al. Circulating TNF receptors 1 and 2 predict stage 3 CKD in type 1 diabetes. J Am Soc Nephrol 2012; 23: 516–524.

66. Niewczas MA, Gohda T, Skupien J, et al. Circulating TNF receptors 1 and 2 predict ESRD in type 2 diabetes. J Am Soc Nephrol 2012; 23: 507–515.

67. Shankar A, Sun L, Klein BE, et al. Markers of inflammation predict the long-term risk of developing chronic kidney disease: a population-based cohort study. Kidney Int 2011; 80: 1231–1238.

68. Pavkov ME, Weil EJ, Fufaa GD, et al. Tumor necrosis factor receptors 1 and 2 are associated with early glomerular lesions in type 2 diabetes. Kidney Int 2016; 89: 226–234.

69. Pavkov ME, Nelson RG, Knowler WC, et al. Elevation of circulating TNF receptors 1 and 2 increases the risk of end-stage renal disease in American Indians with type 2 diabetes. Kidney Int 2015; 87: 812–819.

70. Schei J, Stefansson VT, Eriksen BO, et al. Association of TNF Receptor 2 and CRP with GFR Decline in the General Nondiabetic Population. Clin J Am Soc Nephrol 2017; 12: 624–634.

71. Kodera R, Shikata K, Kataoka HU, et al. Glucagon-like peptide-1 receptor agonist ameliorates renal injury through its anti-inflammatory action without lowering blood glucose level in a rat model of type 1 diabetes. Diabetologia 2011; 54: 965–978.

