## Supplemental materials for "Molecular Programs of Glomerular Hyperfiltration in Early Diabetic Kidney Disease"

Supplemental Figure 1: Top 30 interconnected genes in HF associated modules

A. Salmon module

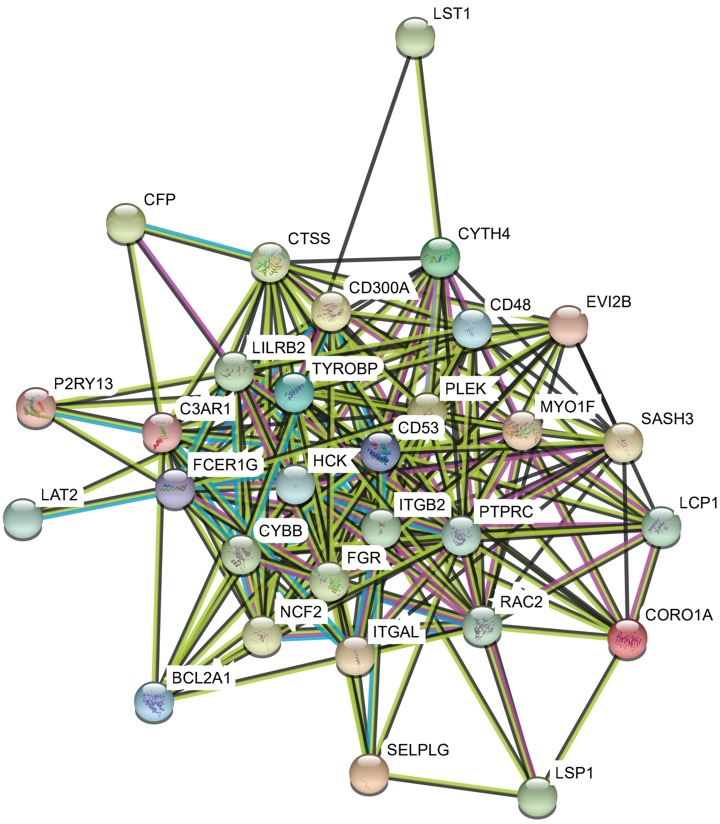


B. Midnight Blue module

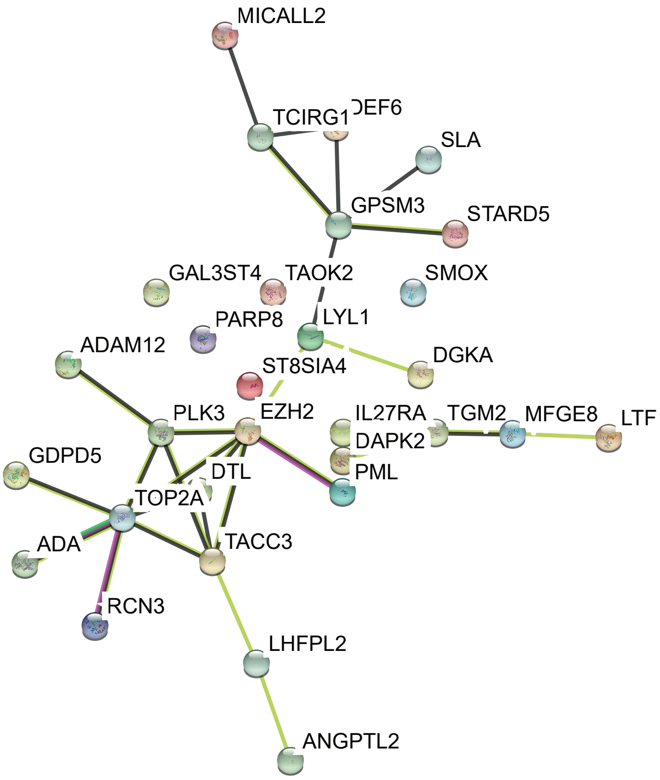


C. Red module

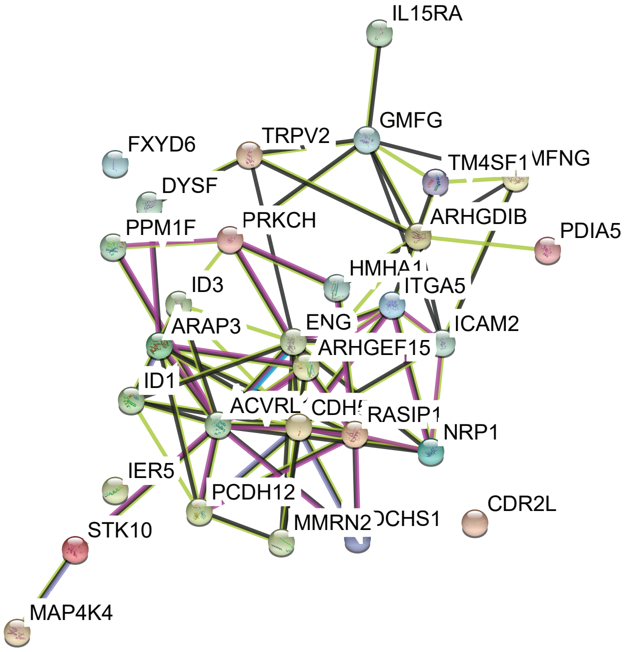


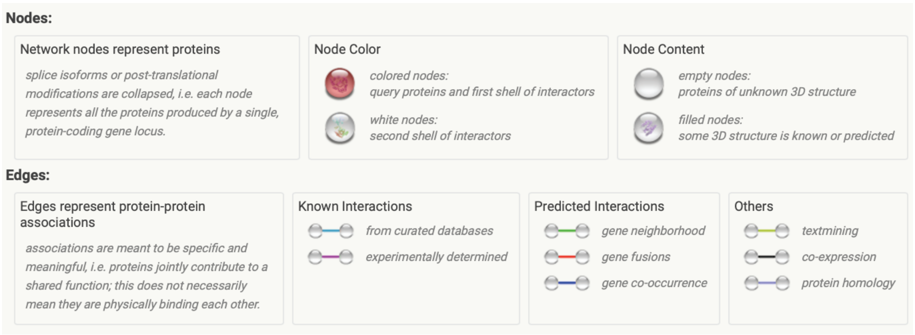

Supplemental Figure 2: Hyperfiltration gene networks and pathways using cytoscape visualization

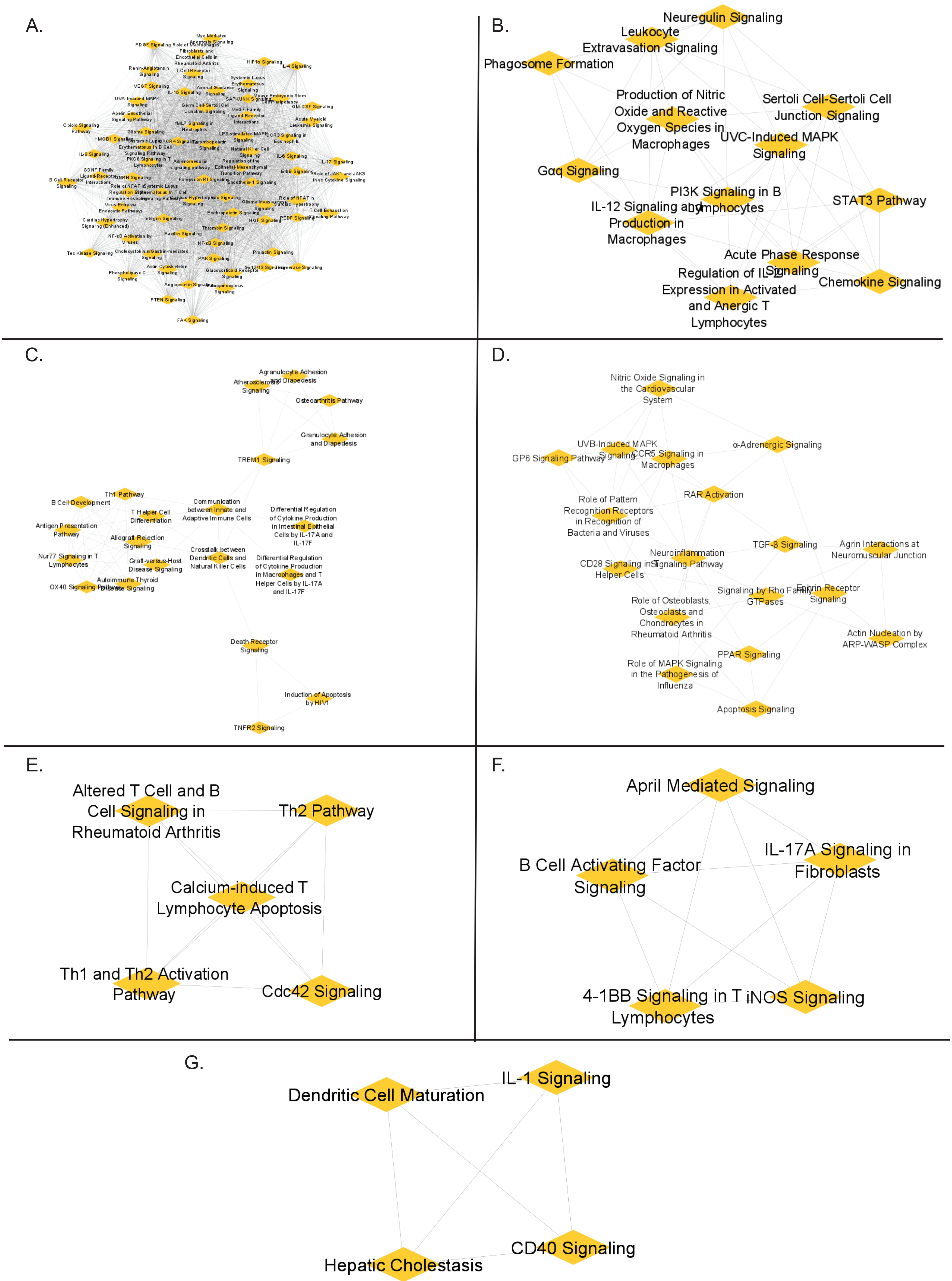

Supplemental Figure 3. NicheNet predicted intracellular targets of endothelial cell activated ligands in crosstalk with mesangial cells

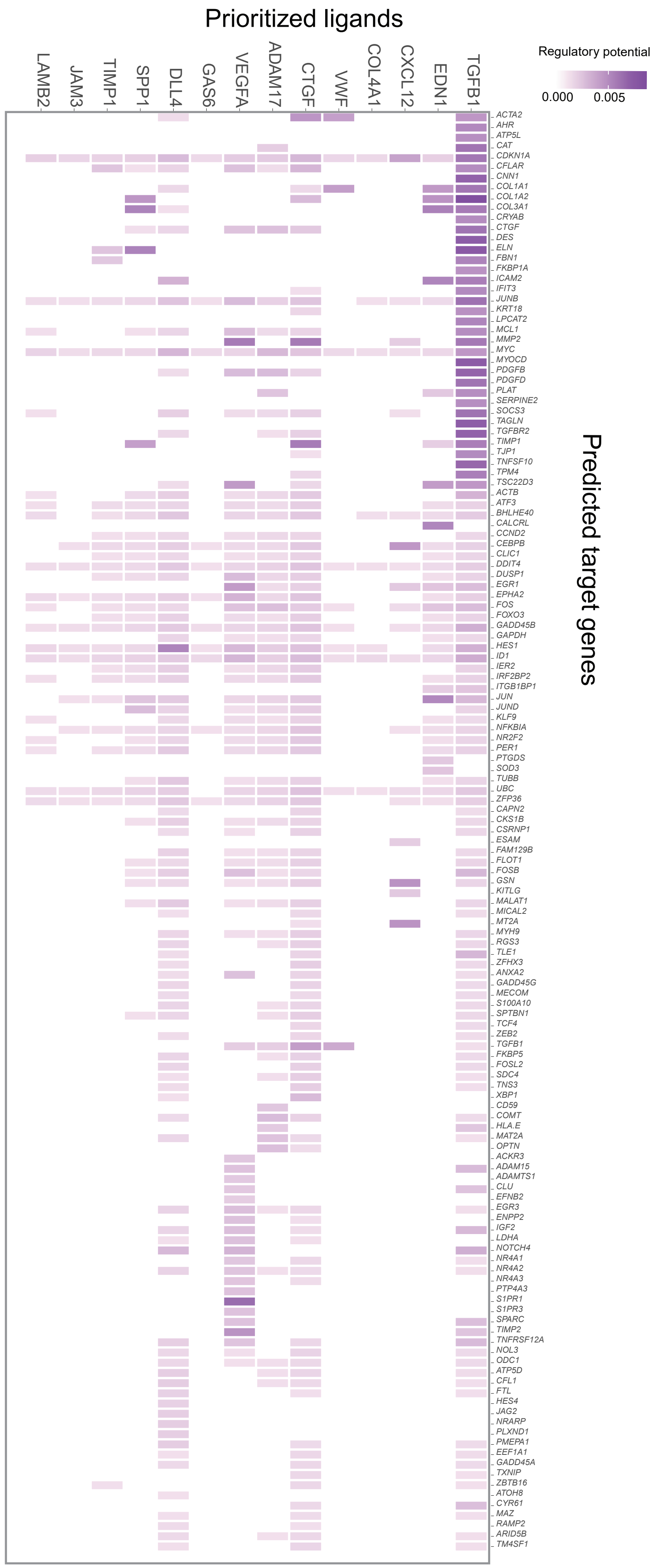

**Supplemental Table 1**: Clinical and morphometric measurements comparing the participants included in the study (peak GFR at or after kidney biopsy) and those who were excluded because their peak GFR occurred < 2 years before the kidney biopsy

| Characteristic | Included (n=52) | Excluded (n=32) | p-value |
|---|---|---|---|
| Male sex (%) | 10 (19.2%) | 10 (31.3%) | 0.21 |
| Age (years) | 44.7 ± 10.3 | 48.9 ± 9.2 | 0.07 |
| Diabetes duration (years) | 13.4 ± 4.0 | 18.6 ± 7.4 | <0.001 |
| BMI (kg/m$^2$) | 37.1 ± 8.5 | 35.5 ± 7.8 | 0.40 |
| Systolic blood pressure (mmHg) | 121 ± 8 | 122 ± 12 | 0.55 |
| Diastolic blood pressure (mmHg) | 76 ± 5 | 77 ± 6 | 0.39 |
| HbA1c (%) | 9.3 ± 2.0 | 9.1 ± 1.6 | 0.67 |
| GFR (ml/min) | 163 ± 44 | 137 ± 40 | 0.007 |
| ACR (mg/g) | 32 (14-71) | 21 (11-71) | 0.57 |
| RAS use (%) | 38 (73.1%) | 27 (84.4%) | 0.23 |
| Mean glomerular volume ($10^6$ μm$^3$) | 2.38 ± 0.72* | 2.27 ± 0.74† | 0.49 |
| Glomerular basement membrane width (nm) | 456 ± 86 | 458 ± 93 | 0.92 |
| Mesangial fractional volume per glomerulus (%) | 0.25 ± 0.06 | 0.27 ± 0.06 | 0.23 |
| Cortical interstitial fractional volume (%) | 0.18 ± 0.04‡ | 0.19 ± 0.05† | 0.07 |
| Glomerular filtration surface density (μm$^2$/μm$^3$) | 0.11 ± 0.04 | 0.10 ± 0.03 | 0.27 |
| Foot process width (nm) | 740 (634, 1014) | 768 (575, 1000) | 0.55 |
| Glomerular podocyte fractional volume (%) | 0.17 ± 0.05 | 0.14 ± 0.04 | 0.032 |
| Polocyte number density per glomerulus ($10^6$ μm$^3$) | 139 ± 72 | 144 ± 90 | 0.76 |
| Fenestrated endothelium (%) | 44.0 ± 19.1 | 44.4 ± 16.4 | 0.92 |

Values are means ± standard deviation or median (interquartile range).
Abbreviations: ACR = albumin:creatinine ratio; BMI = body mass index; GBM = glomerular basement membrane; GFR = glomerular filtration rate; RAS = renin angiotensin system blockers.
*n=46, †n=31, ‡n=49
*p*-value for differences between groups based on *t*-test for continuous variables and chi square for categorical variables. ACR and foot process width were log transformed prior to analysis.

**Supplemental Table 2:** Timing of peak measured GFR compared to timing of peak eGFR in relation to kidney biopsy

| | | Timing of peak eGFR | | |
|---|---|---|---|---|
| | | >2 years before biopsy | Within 2 years of biopsy | >2 years after biopsy |
| Timing of peak measured GFR | >2 years before biopsy | 31 | 0 | 1 |
| | Within 2 years of biopsy | 22 | 1 | 3 |
| | >2 years after biopsy | 18 | 3 | 5 |

Abbreviations: GFR = glomerular filtration rate directly measured with iothalamate clearance; eGFR = estimated glomerular filtration rate calculated using the CKD-epi equation (eGFR = 141 × min $(Scr/k)^{\alpha}$ × $\max(Scr/k)^{-1.209}$ × $0.993^{Age}$ × 1.018 (if female). For males k = 0.9 and α = -0.411; for females k = 0.7 and α = -0.329.

**Supplemental Table 3**: Clinical and morphometric measures at the time of biopsy for the subset of participants with gene expression data compared to those without expression data

| Characteristic | No expression data (n=23) | Expression data (n=29) | *p*-value |
|---|---|---|---|
| Male sex (%) | 6 (26.1%) | 4 (13.8%) | 0.21 |
| Age (years) | 49.1 ± 10.8 | 41.2 ± 8.6 | 0.005 |
| Diabetes duration (years) | 15.0 ± 4.6 | 12.2 ± 2.9 | 0.016 |
| BMI ($kg/m^2$) | 36.5 ± 6.6 | 37.5 ± 9.9 | 0.66 |
| Systolic blood pressure (mmHg) | 123 ± 6 | 119 ± 9 | 0.09 |
| Diastolic blood pressure (mmHg) | 76 ± 5 | 76 ± 5 | 0.58 |
| HbA1c (%) | 9.1 ± 2.0 | 9.4 ± 2.0 | 0.63 |
| GFR (ml/min) | 160 ± 46 | 165 ± 42 | 0.65 |
| ACR (mg/g) | 36 (11, 68) | 31 (18, 73) | 0.65 |
| RAS use (%) | 16 (69.6%) | 22 (75.9%) | 0.23 |
| Mean glomerular volume ($10^6$ $\mu m^3$) | 2.40 ± 0.76* | 2.37 ± 0.70† | 0.92 |
| Glomerular basement membrane width (nm) | 460 ± 663 | 452 ± 100 | 0.74 |
| Mesangial fractional volume per glomerulus (%) | 0.26 ± 0.06 | 0.25 ± 0.05 | 0.52 |
| Cortical interstitial fractional volume (%) | 0.18 ± 0.04‡ | 0.17 ± 0.03§ | 0.51 |
| Glomerular filtration surface density ($\mu m^2/\mu m^3$) | 0.13 ± 0.05 | 0.10 ± 0.02 | 0.021 |
| Foot process width (nm) | 777 (696, 1030) | 716 (564, 908) | 0.28 |
| Glomerular podocyte fractional volume (%) | 0.17 ± 0.04 | 0.16 ± 0.06 | 0.55 |
| Podocyte number density per glomerulus ($10^6$ $\mu m^3$) | 130 ± 69 | 145 ± 74 | 0.44 |
| Fenestrated endothelium (%) | 35.8 ± 17.3 | 50.5 ± 18.2 | 0.005 |

Values are means ± standard deviation or median (interquartile range).
Abbreviations: ACR = albumin:creatinine ratio; BMI = body mass index; GBM = glomerular basement membrane; GFR = glomerular filtration rate; RAS = renin angiotensin system blockers.
*n=21, †n=25, ‡n=22, §n=27
*p*-value for differences between groups based on t-test for continuous variables and chi square for categorical variables. ACR and foot process width were log transformed prior to analysis.

**Supplemental Table 4**: Clinical and morphometric measures at the time of biopsy by peak measured GFR group for the subset of participants with gene expression data

| Characteristic | Hyperfiltration after biopsy (n=15) | Hyperfiltration at biopsy (n=14) | *p*-value |
|---|---|---|---|
| Male sex (%) | 3 (20.0%) | 1 (7.1%) | 0.32 |
| Age (years) | 42.7 ± 6.7 | 39.7 ± 10.2 | 0.36 |
| Diabetes duration (years) | 11.5 ± 2.6 | 12.9 ± 3.1 | 0.19 |
| BMI ($kg/m^2$) | 36.2 ± 9.6 | 38.9 ± 10.3 | 0.47 |
| Systolic blood pressure (mmHg) | 118 ± 9 | 121 ± 10 | 0.36 |
| Diastolic blood pressure (mmHg) | 75 ± 5 | 77 ± 4 | 0.09 |
| HbA1c (%) | 8.5 ± 1.9 | 10.4 ± 1.6 | 0.006 |
| GFR (ml/min) | 152 ± 36 | 180 ± 44 | 0.07 |
| ACR (mg/g) | 24 (13, 42) | 51 (23, 77) | 0.05 |
| RAS use (%) | 14 (93.3%) | 8 (57.1%) | 0.035 |
| Mean glomerular volume ($10^6$ $\mu m^3$) | 2.13 ± 0.69* | 2.60 ± 0.6† | 0.09 |
| Glomerular basement membrane width (nm) | 422 ± 93 | 485 ± 101 | 0.09 |
| Mesangial fractional volume per glomerulus (%) | 0.23 ± 0.05 | 0.27 ± 0.05 | 0.06 |
| Cortical interstitial fractional volume (%) | 0.17 ± 0.02* | 0.18 ± 0.04 | 0.41 |
| Glomerular filtration surface density ($\mu m^2/\mu m^3$) | 0.10 ± 0.02 | 0.10 ± 0.02 | 0.61 |
| Foot process width (nm) | 713 (560, 908) | 747 (564, 916) | 0.96 |
| Glomerular podocyte fractional volume (%) | 0.18 ± 0.07 | 0.15 ± 0.03 | 0.11 |
| Podocyte number density per glomerulus ($10^6$ $\mu m^3$) | 181 ± 81 | 108 ± 43 | 0.006 |
| Fenestrated endothelium (%) | 51.0 ± 16.0 | 49.8 ± 21.0 | 0.85 |

Values are means ± standard deviation or median (interquartile range).
Abbreviations: ACR = albumin:creatinine ratio; BMI = body mass index; GBM = glomerular basement membrane; GFR = glomerular filtration rate; RAS = renin angiotensin system blockers.
*n=13, †n=12
*p*-value for differences between groups based on t-test for continuous variables and chi square for categorical variables except for RAS use which was compared by Fisher's exact test. ACR and foot process width were log transformed prior to analysis.

**Supplemental Table 5: Complete list of genes in the three WGCNA modules where 95% genes (1240 genes) were upregulated in the HF compared to pre-HF group**

| GeneID | Symbol | ModuleAssignment |
|---|---|---|
| 10112 | KIF20A | midnightblue |
| 2146 | EZH2 | midnightblue |
| 8838 | WISP3 | midnightblue |
| 1356 | CP | midnightblue |
| 59 | ACTA2 | midnightblue |
| 2305 | FOXM1 | midnightblue |
| 9173 | IL1RL1 | midnightblue |
| 9180 | OSMR | midnightblue |
| 79727 | LIN28A | midnightblue |
| 5118 | PCOLCE | midnightblue |
| 79908 | BTNL8 | midnightblue |
| 10095 | ARPC1B | midnightblue |
| 79690 | GAL3ST4 | midnightblue |
| 7153 | TOP2A | midnightblue |
| 55872 | PBK | midnightblue |
| 391020 | LOC391020 | midnightblue |
| 3240 | HP | midnightblue |
| 70 | ACTC1 | midnightblue |
| 10633 | RASL10A | midnightblue |
| 4609 | MYC | midnightblue |
| 79850 | FAM57A | midnightblue |
| 332 | BIRC5 | midnightblue |
| 6241 | RRM2 | midnightblue |
| 80774 | LIMD2 | midnightblue |
| 1844 | DUSP2 | midnightblue |
| 11065 | UBE2C | midnightblue |
| 50619 | DEF6 | midnightblue |
| 7447 | VSNL1 | midnightblue |
| 79075 | DSCC1 | midnightblue |
| 51514 | DTL | midnightblue |
| 253982 | ASPHD1 | midnightblue |
| 57706 | DENND1A | midnightblue |
| 81930 | KIF18A | midnightblue |
| 29902 | FAM216A | midnightblue |
| 10205 | MPZL2 | midnightblue |
| 12 | SERPINA3 | midnightblue |

| | | |
|---|---|---|
| 9133 | CCNB2 | midnightblue |
| 51274 | KLF3 | midnightblue |
| 952 | CD38 | midnightblue |
| 57556 | SEMA6A | midnightblue |
| 2004 | ELK3 | midnightblue |
| 2242 | FES | midnightblue |
| 9181 | ARHGEF2 | midnightblue |
| 4240 | MFGE8 | midnightblue |
| 841 | CASP8 | midnightblue |
| 55240 | STEAP3 | midnightblue |
| 55711 | FAR2 | midnightblue |
| 5939 | RBMS2 | midnightblue |
| 79955 | PDZD7 | midnightblue |
| 8605 | PLA2G4C | midnightblue |
| 995 | CDC25C | midnightblue |
| 84444 | DOT1L | midnightblue |
| 6362 | CCL18 | midnightblue |
| 27036 | SIGLEC7 | midnightblue |
| 10785 | WDR4 | midnightblue |
| 4615 | MYD88 | midnightblue |
| 6489 | ST8SIA1 | midnightblue |
| 6509 | SLC1A4 | midnightblue |
| 11254 | SLC6A14 | midnightblue |
| 79668 | PARP8 | midnightblue |
| 10246 | SLC17A2 | midnightblue |
| 4313 | MMP2 | midnightblue |
| 56886 | UGGT1 | midnightblue |
| 79683 | ZDHHC14 | midnightblue |
| 51564 | HDAC7 | midnightblue |
| 3978 | LIG1 | midnightblue |
| 10493 | VAT1 | midnightblue |
| 80765 | STARD5 | midnightblue |
| 58189 | WFDC1 | midnightblue |
| 80111 | C3orf36 | midnightblue |
| 51291 | GMIP | midnightblue |
| 54498 | SMOX | midnightblue |
| 27185 | DISC1 | midnightblue |
| 9138 | ARHGEF1 | midnightblue |
| 1788 | DNMT3A | midnightblue |
| 7052 | TGM2 | midnightblue |
| 3801 | KIFC3 | midnightblue |

| | | |
|---|---|---|
| 4314 | MMP3 | midnightblue |
| 55190 | NUDT11 | midnightblue |
| 81029 | WNT5B | midnightblue |
| 9997 | SCO2 | midnightblue |
| 1949 | EFNB3 | midnightblue |
| 54187 | NANS | midnightblue |
| 7042 | TGFB2 | midnightblue |
| 10799 | RPP40 | midnightblue |
| 7903 | ST8SIA4 | midnightblue |
| 3710 | ITPR3 | midnightblue |
| 400506 | KNOP1 | midnightblue |
| 7408 | VASP | midnightblue |
| 8988 | HSPB3 | midnightblue |
| 3003 | GZMK | midnightblue |
| 5155 | PDGFB | midnightblue |
| 6134 | RPL10 | midnightblue |
| 53820 | RIPPLY3 | midnightblue |
| 80868 | HCG4B | midnightblue |
| 51332 | SPTBN5 | midnightblue |
| 80256 | FAM214B | midnightblue |
| 10287 | RGS19 | midnightblue |
| 5371 | PML | midnightblue |
| 6533 | SLC6A6 | midnightblue |
| 6503 | SLA | midnightblue |
| 9645 | MICAL2 | midnightblue |
| 79047 | KCTD15 | midnightblue |
| 57596 | BEGAIN | midnightblue |
| 4066 | LYL1 | midnightblue |
| 1263 | PLK3 | midnightblue |
| 9219 | MTA2 | midnightblue |
| 2319 | FLOT2 | midnightblue |
| 1302 | COL11A2 | midnightblue |
| 79847 | TMEM180 | midnightblue |
| 2000 | ELF4 | midnightblue |
| 7004 | TEAD4 | midnightblue |
| 22839 | DLGAP4 | midnightblue |
| 5055 | SERPINB2 | midnightblue |
| 23166 | STAB1 | midnightblue |
| 843 | CASP10 | midnightblue |
| 5979 | RET | midnightblue |
| 11186 | RASSF1 | midnightblue |

| | | |
|---|---|---|
| 79825 | EFCC1 | midnightblue |
| 79879 | CCDC134 | midnightblue |
| 63940 | GPSM3 | midnightblue |
| 57172 | CAMK1G | midnightblue |
| 5365 | PLXNB3 | midnightblue |
| 2245 | FGD1 | midnightblue |
| 11182 | SLC2A6 | midnightblue |
| 84823 | LMNB2 | midnightblue |
| 990 | CDC6 | midnightblue |
| 29781 | NCAPH2 | midnightblue |
| 7010 | TEK | midnightblue |
| 23329 | TBC1D30 | midnightblue |
| 604 | BCL6 | midnightblue |
| 1841 | DTYMK | midnightblue |
| 2077 | ERF | midnightblue |
| 55320 | MIS18BP1 | midnightblue |
| 8091 | HMGA2 | midnightblue |
| 639 | PRDM1 | midnightblue |
| 9344 | TAOK2 | midnightblue |
| 489 | ATP2A3 | midnightblue |
| 10556 | RPP30 | midnightblue |
| 7873 | MANF | midnightblue |
| 11156 | PTP4A3 | midnightblue |
| 81544 | GDPD5 | midnightblue |
| 79905 | TMC7 | midnightblue |
| 4776 | NFATC4 | midnightblue |
| 64748 | LPPR2 | midnightblue |
| 28985 | MCTS1 | midnightblue |
| 55850 | USE1 | midnightblue |
| 8884 | SLC5A6 | midnightblue |
| 9948 | WDR1 | midnightblue |
| 55357 | TBC1D2 | midnightblue |
| 22808 | MRAS | midnightblue |
| 29780 | PARVB | midnightblue |
| 6929 | TCF3 | midnightblue |
| 50515 | CHST11 | midnightblue |
| 5130 | PCYT1A | midnightblue |
| 79901 | CYBRD1 | midnightblue |
| 3603 | IL16 | midnightblue |
| 91300 | R3HDM4 | midnightblue |
| 11339 | OIP5 | midnightblue |

| | | |
|---|---|---|
| 920 | CD4 | midnightblue |
| 7857 | SCG2 | midnightblue |
| 10184 | LHFPL2 | midnightblue |
| 4058 | LTK | midnightblue |
| 5916 | RARG | midnightblue |
| 23351 | KHNYN | midnightblue |
| 5993 | RFX5 | midnightblue |
| 7083 | TK1 | midnightblue |
| 57333 | RCN3 | midnightblue |
| 2331 | FMOD | midnightblue |
| 79927 | FAM110D | midnightblue |
| 2788 | GNG7 | midnightblue |
| 7299 | TYR | midnightblue |
| 79682 | CENPU | midnightblue |
| 4858 | NOVA2 | midnightblue |
| 55223 | TRIM62 | midnightblue |
| 4899 | NRF1 | midnightblue |
| 309 | ANXA6 | midnightblue |
| 5742 | PTGS1 | midnightblue |
| 3111 | HLA-DOA | midnightblue |
| 3910 | LAMA4 | midnightblue |
| 25806 | VAX2 | midnightblue |
| 9466 | IL27RA | midnightblue |
| 8310 | ACOX3 | midnightblue |
| 7127 | TNFAIP2 | midnightblue |
| 1606 | DGKA | midnightblue |
| 1789 | DNMT3B | midnightblue |
| 5600 | MAPK11 | midnightblue |
| 5675 | PSG6 | midnightblue |
| 100 | ADA | midnightblue |
| 393 | ARHGAP4 | midnightblue |
| 56548 | CHST7 | midnightblue |
| 26508 | HEYL | midnightblue |
| 141 | ADPRH | midnightblue |
| 2494 | NR5A2 | midnightblue |
| 8038 | ADAM12 | midnightblue |
| 408 | ARRB1 | midnightblue |
| 90379 | DCAF15 | midnightblue |
| 55188 | RIC8B | midnightblue |
| 83448 | PUS7L | midnightblue |
| 8874 | ARHGEF7 | midnightblue |

| | | |
|---|---|---|
| 57118 | CAMK1D | midnightblue |
| 863 | CBFA2T3 | midnightblue |
| 79159 | NOL12 | midnightblue |
| 29109 | FHOD1 | midnightblue |
| 51559 | NT5DC3 | midnightblue |
| 1183 | CLCN4 | midnightblue |
| 11169 | WDHD1 | midnightblue |
| 5271 | SERPINB8 | midnightblue |
| 50488 | MINK1 | midnightblue |
| 23452 | ANGPTL2 | midnightblue |
| 6574 | SLC20A1 | midnightblue |
| 83729 | INHBE | midnightblue |
| 4741 | NEFM | midnightblue |
| 8882 | ZPR1 | midnightblue |
| 56882 | CDC42SE1 | midnightblue |
| 9013 | TAF1C | midnightblue |
| 84617 | TUBB6 | midnightblue |
| 3455 | IFNAR2 | midnightblue |
| 1445 | CSK | midnightblue |
| 3574 | IL7 | midnightblue |
| 8940 | TOP3B | midnightblue |
| 8515 | ITGA10 | midnightblue |
| 22846 | VASH1 | midnightblue |
| 9057 | SLC7A6 | midnightblue |
| 11230 | PRAF2 | midnightblue |
| 51755 | CDK12 | midnightblue |
| 7040 | TGFB1 | midnightblue |
| 3227 | HOXC11 | midnightblue |
| 10046 | MAMLD1 | midnightblue |
| 4828 | NMB | midnightblue |
| 23604 | DAPK2 | midnightblue |
| 8751 | ADAM15 | midnightblue |
| 399664 | MEX3D | midnightblue |
| 4650 | MYO9B | midnightblue |
| 5423 | POLB | midnightblue |
| 1774 | DNASE1L1 | midnightblue |
| 26097 | CHTOP | midnightblue |
| 114899 | C1QTNF3 | midnightblue |
| 11064 | CNTRL | midnightblue |
| 4093 | SMAD9 | midnightblue |
| 54908 | SPDL1 | midnightblue |

| | | |
|---|---|---|
| 2035 | EPB41 | midnightblue |
| 3191 | HNRNPL | midnightblue |
| 602 | BCL3 | midnightblue |
| 79838 | TMC5 | midnightblue |
| 6103 | RPGR | midnightblue |
| 983 | CDK1 | midnightblue |
| 29094 | LGALSL | midnightblue |
| 10533 | ATG7 | midnightblue |
| 2275 | FHL3 | midnightblue |
| 10750 | GRAP | midnightblue |
| 4957 | ODF2 | midnightblue |
| 9928 | KIF14 | midnightblue |
| 865 | CBFB | midnightblue |
| 7450 | VWF | midnightblue |
| 11165 | NUDT3 | midnightblue |
| 577 | ADGRB3 | midnightblue |
| 55117 | SLC6A15 | midnightblue |
| 79778 | MICALL2 | midnightblue |
| 9020 | MAP3K14 | midnightblue |
| 79870 | BAALC | midnightblue |
| 81605 | URM1 | midnightblue |
| 6491 | STIL | midnightblue |
| 2717 | GLA | midnightblue |
| 775 | CACNA1C | midnightblue |
| 55621 | TRMT1 | midnightblue |
| 79899 | PRR5L | midnightblue |
| 64761 | PARP12 | midnightblue |
| 54840 | APTX | midnightblue |
| 10195 | ALG3 | midnightblue |
| 65012 | SLC26A10 | midnightblue |
| 85377 | MICALL1 | midnightblue |
| 23180 | RFTN1 | midnightblue |
| 80169 | CTC1 | midnightblue |
| 10460 | TACC3 | midnightblue |
| 84722 | PSRC1 | midnightblue |
| 55966 | AJAP1 | midnightblue |
| 8795 | TNFRSF10B | midnightblue |
| 3339 | HSPG2 | midnightblue |
| 54478 | FAM64A | midnightblue |
| 949 | SCARB1 | midnightblue |
| 10312 | TCIRG1 | midnightblue |

| | | |
|---|---|---|
| 2634 | GBP2 | midnightblue |
| 23467 | NPTXR | midnightblue |
| 3832 | KIF11 | midnightblue |
| 9363 | RAB33A | midnightblue |
| 6659 | SOX4 | midnightblue |
| 4815 | NINJ2 | midnightblue |
| 25802 | LMOD1 | midnightblue |
| 57099 | AVEN | midnightblue |
| 25796 | PGLS | midnightblue |
| 10535 | RNASEH2A | midnightblue |
| 8450 | CUL4B | midnightblue |
| 3592 | IL12A | midnightblue |
| 51067 | YARS2 | midnightblue |
| 54674 | LRRN3 | midnightblue |
| 26032 | SUSD5 | midnightblue |
| 53346 | TM6SF1 | midnightblue |
| 1259 | CNGA1 | midnightblue |
| 11013 | TMSB15A | midnightblue |
| 9263 | STK17A | midnightblue |
| 55143 | CDCA8 | midnightblue |
| 4900 | NRGN | midnightblue |
| 27351 | DESI1 | midnightblue |
| 3460 | IFNGR2 | midnightblue |
| 4057 | LTF | midnightblue |
| 5026 | P2RX5 | midnightblue |
| 578 | BAK1 | midnightblue |
| 1663 | DDX11 | midnightblue |
| 29015 | SLC43A3 | midnightblue |
| 4794 | NFKBIE | midnightblue |
| 9134 | CCNE2 | midnightblue |
| 313 | AOAH | midnightblue |
| 5873 | RAB27A | midnightblue |
| 8349 | HIST2H2BE | midnightblue |
| 3620 | IDO1 | midnightblue |
| 10643 | IGF2BP3 | midnightblue |
| 1290 | COL5A2 | midnightblue |
| 8821 | INPP4B | midnightblue |
| 22943 | DKK1 | midnightblue |
| 7941 | PLA2G7 | midnightblue |
| 10801 | 9-Sep | midnightblue |
| 28442 | IGHV3-23 | red |

| | | |
|---|---|---|
| 81831 | NETO2 | red |
| 6876 | TAGLN | red |
| 929 | CD14 | red |
| 9314 | KLF4 | red |
| 57016 | AKR1B10 | red |
| 51299 | NRN1 | red |
| 7805 | LAPTM5 | red |
| 3059 | HCLS1 | red |
| 7852 | CXCR4 | red |
| 55340 | GIMAP5 | red |
| 54541 | DDIT4 | red |
| 6578 | SLCO2A1 | red |
| 1901 | S1PR1 | red |
| 3122 | HLA-DRA | red |
| 8692 | HYAL2 | red |
| 2167 | FABP4 | red |
| 5328 | PLAU | red |
| 5552 | SRGN | red |
| 8322 | FZD4 | red |
| 1051 | CEBPB | red |
| 8516 | ITGA8 | red |
| 4629 | MYH11 | red |
| 51296 | SLC15A3 | red |
| 355 | FAS | red |
| 3428 | IFI16 | red |
| 6376 | CX3CL1 | red |
| 390 | RND3 | red |
| 4811 | NID1 | red |
| 54517 | PUS7 | red |
| 595 | CCND1 | red |
| 11067 | C10orf10 | red |
| 3185 | HNRNPF | red |
| 8291 | DYSF | red |
| 185 | AGTR1 | red |
| 3115 | HLA-DPB1 | red |
| 6507 | SLC1A3 | red |
| 26207 | PITPNC1 | red |
| 8875 | VNN2 | red |
| 972 | CD74 | red |
| 51705 | EMCN | red |
| 8503 | PIK3R3 | red |

| | | |
|---|---|---|
| 54453 | RIN2 | red |
| 6035 | RNASE1 | red |
| 79652 | TMEM204 | red |
| 2995 | GYPC | red |
| 3384 | ICAM2 | red |
| 3119 | HLA-DQB1 | red |
| 834 | CASP1 | red |
| 27122 | DKK3 | red |
| 26251 | KCNG2 | red |
| 11138 | TBC1D8 | red |
| 109 | ADCY3 | red |
| 8743 | TNFSF10 | red |
| 3123 | HLA-DRB1 | red |
| 3429 | IFI27 | red |
| 7088 | TLE1 | red |
| 4145 | MATK | red |
| 2013 | EMP2 | red |
| 23643 | LY96 | red |
| 6926 | TBX3 | red |
| 10536 | P3H3 | red |
| 23208 | SYT11 | red |
| 8835 | SOCS2 | red |
| 27303 | RBMS3 | red |
| 2687 | GGT5 | red |
| 221395 | ADGRF5 | red |
| 129080 | EMID1 | red |
| 28232 | SLCO3A1 | red |
| 64061 | TSPYL2 | red |
| 55843 | ARHGAP15 | red |
| 947 | CD34 | red |
| 3182 | HNRNPAB | red |
| 79812 | MMRN2 | red |
| 2050 | EPHB4 | red |
| 11007 | CCDC85B | red |
| 80310 | PDGFD | red |
| 581 | BAX | red |
| 3397 | ID1 | red |
| 80723 | SLC35G2 | red |
| 27065 | NSG1 | red |
| 2040 | STOM | red |
| 25960 | ADGRA2 | red |

| | | |
|---|---|---|
| 1293 | COL6A3 | red |
| 8446 | DUSP11 | red |
| 6939 | TCF15 | red |
| 80177 | MYCT1 | red |
| 10875 | FGL2 | red |
| 4052 | LTBP1 | red |
| 2313 | FLI1 | red |
| 5175 | PECAM1 | red |
| 4242 | MFNG | red |
| 3554 | IL1R1 | red |
| 10019 | SH2B3 | red |
| 79172 | CENPO | red |
| 3128 | HLA-DRB6 | red |
| 54922 | RASIP1 | red |
| 80833 | APOL3 | red |
| 25992 | SNED1 | red |
| 55303 | GIMAP4 | red |
| 8573 | CASK | red |
| 2014 | EMP3 | red |
| 5915 | RARB | red |
| 64174 | DPEP2 | red |
| 5613 | PRKX | red |
| 27190 | IL17B | red |
| 55691 | FRMD4A | red |
| 6300 | MAPK12 | red |
| 1193 | CLIC2 | red |
| 347544 | RPL18AP16 | red |
| 10580 | SORBS1 | red |
| 3488 | IGFBP5 | red |
| 9771 | RAPGEF5 | red |
| 5327 | PLAT | red |
| 1071 | CETP | red |
| 367 | AR | red |
| 2702 | GJA5 | red |
| 9448 | MAP4K4 | red |
| 3398 | ID2 | red |
| 6236 | RRAD | red |
| 8543 | LMO4 | red |
| 9590 | AKAP12 | red |
| 51002 | TPRKB | red |
| 284403 | WDR62 | red |

| | | |
|---|---|---|
| 1393 | CRHBP | red |
| 22845 | DOLK | red |
| 6672 | SP100 | red |
| 2828 | GPR4 | red |
| 8805 | TRIM24 | red |
| 55196 | KIAA1551 | red |
| 26578 | OSTF1 | red |
| 9767 | JADE3 | red |
| 54538 | ROBO4 | red |
| 3430 | IFI35 | red |
| 54863 | TOR4A | red |
| 22918 | CD93 | red |
| 474344 | GIMAP6 | red |
| 874 | CBR3 | red |
| 2114 | ETS2 | red |
| 871 | SERPINH1 | red |
| 30850 | CDR2L | red |
| 51192 | CKLF | red |
| 1305 | COL13A1 | red |
| 1030 | CDKN2B | red |
| 51474 | LIMA1 | red |
| 2324 | FLT4 | red |
| 9899 | SV2B | red |
| 6403 | SELP | red |
| 9844 | ELMO1 | red |
| 10231 | RCAN2 | red |
| 51703 | ACSL5 | red |
| 8536 | CAMK1 | red |
| 9580 | SOX13 | red |
| 23261 | CAMTA1 | red |
| 64129 | TINAGL1 | red |
| 4256 | MGP | red |
| 23597 | ACOT9 | red |
| 26052 | DNM3 | red |
| 104 | ADARB1 | red |
| 23063 | WAPAL | red |
| 57198 | ATP8B2 | red |
| 22900 | CARD8 | red |
| 5287 | PIK3C2B | red |
| 79974 | CPED1 | red |
| 9535 | GMFG | red |

| | | |
|---|---|---|
| 8436 | SDPR | red |
| 5583 | PRKCH | red |
| 3937 | LCP2 | red |
| 6197 | RPS6KA3 | red |
| 8125 | ANP32A | red |
| 5967 | REG1A | red |
| 3459 | IFNGR1 | red |
| 53826 | FXYD6 | red |
| 27292 | DIMT1 | red |
| 50807 | ASAP1 | red |
| 5899 | RALB | red |
| 10365 | KLF2 | red |
| 397 | ARHGDIB | red |
| 9537 | TP53I11 | red |
| 9938 | ARHGAP25 | red |
| 54910 | SEMA4C | red |
| 3714 | JAG2 | red |
| 3215 | HOXB5 | red |
| 56901 | NDUFA4L2 | red |
| 1965 | EIF2S1 | red |
| 2294 | FOXF1 | red |
| 2887 | GRB10 | red |
| 11322 | TMC6 | red |
| 11177 | BAZ1A | red |
| 2824 | GPM6B | red |
| 29108 | PYCARD | red |
| 9976 | CLEC2B | red |
| 2157 | F8 | red |
| 10567 | RABAC1 | red |
| 64411 | ARAP3 | red |
| 65108 | MARCKSL1 | red |
| 4212 | MEIS2 | red |
| 3725 | JUN | red |
| 5228 | PGF | red |
| 1009 | CDH11 | red |
| 22861 | NLRP1 | red |
| 9510 | ADAMTS1 | red |
| 79630 | C1orf54 | red |
| 79005 | SCNM1 | red |
| 8829 | NRP1 | red |
| 4893 | NRAS | red |

| | | |
|---|---|---|
| 1535 | CYBA | red |
| 10161 | LPAR6 | red |
| 51393 | TRPV2 | red |
| 10384 | BTN3A3 | red |
| 7126 | TNFAIP1 | red |
| 4147 | MATN2 | red |
| 2247 | FGF2 | red |
| 57523 | NYNRIN | red |
| 2037 | EPB41L2 | red |
| 9124 | PDLIM1 | red |
| 55577 | NAGK | red |
| 4855 | NOTCH4 | red |
| 958 | CD40 | red |
| 3109 | HLA-DMB | red |
| 10954 | PDIA5 | red |
| 4149 | MAX | red |
| 2523 | FUT1 | red |
| 51465 | UBE2J1 | red |
| 26112 | CCDC69 | red |
| 6591 | SNAI2 | red |
| 2876 | GPX1 | red |
| 9337 | CNOT8 | red |
| 7322 | UBE2D2 | red |
| 92822 | ZNF276 | red |
| 3481 | IGF2 | red |
| 2113 | ETS1 | red |
| 2117 | ETV3 | red |
| 23076 | RRP1B | red |
| 7099 | TLR4 | red |
| 442175 | RPLP2P1 | red |
| 7075 | TIE1 | red |
| 5836 | PYGL | red |
| 9710 | KIAA0355 | red |
| 2621 | GAS6 | red |
| 10212 | DDX39A | red |
| 10491 | CRTAP | red |
| 8563 | THOC5 | red |
| 4851 | NOTCH1 | red |
| 4065 | LY75 | red |
| 7293 | TNFRSF4 | red |
| 3113 | HLA-DPA1 | red |

| | | |
|---|---|---|
| 51499 | TRIAP1 | red |
| 7089 | TLE2 | red |
| 10203 | CALCRL | red |
| 22903 | BTBD3 | red |
| 57048 | PLSCR3 | red |
| 3628 | INPP1 | red |
| 7102 | TSPAN7 | red |
| 3108 | HLA-DMA | red |
| 8541 | PPFIA3 | red |
| 10528 | NOP56 | red |
| 5797 | PTPRM | red |
| 3383 | ICAM1 | red |
| 51155 | HN1 | red |
| 2791 | GNG11 | red |
| 94 | ACVRL1 | red |
| 6256 | RXRA | red |
| 3678 | ITGA5 | red |
| 10634 | GAS2L1 | red |
| 56936 | CCDC177 | red |
| 9034 | CCRL2 | red |
| 10280 | SIGMAR1 | red |
| 5728 | PTEN | red |
| 6397 | SEC14L1 | red |
| 3601 | IL15RA | red |
| 219654 | ZCCHC24 | red |
| 2683 | B4GALT1 | red |
| 6793 | STK10 | red |
| 1396 | CRIP1 | red |
| 135 | ADORA2A | red |
| 26277 | TINF2 | red |
| 3566 | IL4R | red |
| 23462 | HEY1 | red |
| 11259 | FILIP1L | red |
| 2624 | GATA2 | red |
| 7090 | TLE3 | red |
| 57228 | SMAGP | red |
| 51278 | IER5 | red |
| 2615 | LRRC32 | red |
| 10266 | RAMP2 | red |
| 4208 | MEF2C | red |
| 3399 | ID3 | red |

| | | |
|---|---|---|
| 974 | CD79B | red |
| 4651 | MYO10 | red |
| 7123 | CLEC3B | red |
| 23333 | DPY19L1 | red |
| 9630 | GNA14 | red |
| 55885 | LMO3 | red |
| 5156 | PDGFRA | red |
| 4056 | LTC4S | red |
| 4071 | TM4SF1 | red |
| 51763 | INPP5K | red |
| 55790 | CSGALNACT1 | red |
| 4170 | MCL1 | red |
| 9754 | STARD8 | red |
| 493 | ATP2B4 | red |
| 6525 | SMTN | red |
| 9555 | H2AFY | red |
| 634 | CEACAM1 | red |
| 7433 | VIPR1 | red |
| 2078 | ERG | red |
| 4301 | MLLT4 | red |
| 79677 | SMC6 | red |
| 4363 | ABCC1 | red |
| 9760 | TOX | red |
| 9886 | RHOBTB1 | red |
| 55813 | UTP6 | red |
| 51100 | SH3GLB1 | red |
| 10003 | NAALAD2 | red |
| 7329 | UBE2I | red |
| 10410 | IFITM3 | red |
| 6886 | TAL1 | red |
| 5912 | RAP2B | red |
| 6405 | SEMA3F | red |
| 391132 | LOC391132 | red |
| 5799 | PTPRN2 | red |
| 404672 | GTF2H5 | red |
| 953 | ENTPD1 | red |
| 6624 | FSCN1 | red |
| 4846 | NOS3 | red |
| 58494 | JAM2 | red |
| 51118 | UTP11L | red |
| 5896 | RAG1 | red |

| | | |
|---|---|---|
| 8642 | DCHS1 | red |
| 57608 | KIAA1462 | red |
| 81577 | GFOD2 | red |
| 7637 | ZNF84 | red |
| 29095 | ORMDL2 | red |
| 9781 | RNF144A | red |
| 9638 | FEZ1 | red |
| 83442 | SH3BGRL3 | red |
| 3992 | FADS1 | red |
| 4330 | MN1 | red |
| 8890 | EIF2B4 | red |
| 3726 | JUNB | red |
| 57513 | CASKIN2 | red |
| 4258 | MGST2 | red |
| 7453 | WARS | red |
| 63895 | PIEZO2 | red |
| 60673 | ATG101 | red |
| 9647 | PPM1F | red |
| 8487 | GEMIN2 | red |
| 6890 | TAP1 | red |
| 55795 | PCID2 | red |
| 9079 | LDB2 | red |
| 55107 | ANO1 | red |
| 7048 | TGFBR2 | red |
| 3270 | HRC | red |
| 22899 | ARHGEF15 | red |
| 3382 | ICA1 | red |
| 965 | CD58 | red |
| 753 | LDLRAD4 | red |
| 8496 | PPFIBP1 | red |
| 442171 | RPL10P2 | red |
| 9644 | SH3PXD2A | red |
| 640 | BLK | red |
| 28960 | DCPS | red |
| 2091 | FBL | red |
| 57157 | PHTF2 | red |
| 10436 | EMG1 | red |
| 8742 | TNFSF12 | red |
| 23179 | RGL1 | red |
| 3487 | IGFBP4 | red |
| 23231 | SEL1L3 | red |

| | | |
|---|---|---|
| 51063 | CALHM2 | red |
| 51177 | PLEKHO1 | red |
| 7706 | TRIM25 | red |
| 55619 | DOCK10 | red |
| 81619 | TSPAN14 | red |
| 2487 | FRZB | red |
| 56911 | MAP3K7CL | red |
| 2 | A2M | red |
| 6940 | ZNF354A | red |
| 3579 | CXCR2 | red |
| 57124 | CD248 | red |
| 51226 | COPZ2 | red |
| 2022 | ENG | red |
| 55269 | PSPC1 | red |
| 256949 | KANK3 | red |
| 6004 | RGS16 | red |
| 5699 | PSMB10 | red |
| 55106 | SLFN12 | red |
| 23526 | HMHA1 | red |
| 5998 | RGS3 | red |
| 1909 | EDNRA | red |
| 51634 | RBMX2 | red |
| 1036 | CDO1 | red |
| 3087 | HHEX | red |
| 976 | ADGRE5 | red |
| 57576 | KIF17 | red |
| 6915 | TBXA2R | red |
| 9530 | BAG4 | red |
| 3707 | ITPKB | red |
| 1054 | CEBPG | red |
| 80301 | PLEKHO2 | red |
| 1432 | MAPK14 | red |
| 6238 | RRBP1 | red |
| 11119 | BTN3A1 | red |
| 30845 | EHD3 | red |
| 57572 | DOCK6 | red |
| 5787 | PTPRB | red |
| 5269 | SERPINB6 | red |
| 1282 | COL4A1 | red |
| 5550 | PREP | red |
| 64943 | NT5DC2 | red |

| | | |
|---|---|---|
| 5698 | PSMB9 | red |
| 51285 | RASL12 | red |
| 7035 | TFPI | red |
| 3133 | HLA-E | red |
| 7056 | THBD | red |
| 51294 | PCDH12 | red |
| 2395 | FXN | red |
| 1003 | CDH5 | red |
| 55785 | FGD6 | red |
| 1235 | CCR6 | red |
| 10109 | ARPC2 | red |
| 22998 | LIMCH1 | red |
| 55038 | CDCA4 | red |
| 4854 | NOTCH3 | red |
| 6480 | ST6GAL1 | red |
| 7168 | TPM1 | red |
| 51267 | CLEC1A | red |
| 6494 | SIPA1 | red |
| 81848 | SPRY4 | red |
| 6158 | RPL28 | red |
| 9168 | TMSB10 | red |
| 4092 | SMAD7 | red |
| 107 | ADCY1 | red |
| 4487 | MSX1 | red |
| 55707 | NECAP2 | red |
| 3791 | KDR | red |
| 3977 | LIFR | red |
| 706 | TSPO | red |
| 10631 | POSTN | red |
| 23683 | PRKD3 | red |
| 54621 | VSIG10 | red |
| 1513 | CTSK | red |
| 54345 | SOX18 | red |
| 5982 | RFC2 | red |
| 9459 | ARHGEF6 | red |
| 187 | APLNR | red |
| 3551 | IKBKB | red |
| 912 | CD1D | red |
| 25966 | C2CD2 | red |
| 1050 | CEBPA | red |
| 1175 | AP2S1 | red |

| | | |
|---|---|---|
| 5770 | PTPN1 | red |
| 8260 | NAA10 | red |
| 4690 | NCK1 | red |
| 1466 | CSRP2 | red |
| 7291 | TWIST1 | red |
| 10228 | STX6 | red |
| 64785 | GINS3 | red |
| 57817 | HAMP | red |
| 56654 | NPDC1 | red |
| 9843 | HEPH | red |
| 30851 | TAX1BP3 | red |
| 9880 | ZBTB39 | red |
| 83468 | GLT8D2 | red |
| 10241 | CALCOCO2 | red |
| 54707 | GPN2 | red |
| 93349 | SP140L | red |
| 3632 | INPP5A | red |
| 1476 | CSTB | red |
| 11118 | BTN3A2 | red |
| 7203 | CCT3 | red |
| 5928 | RBBP4 | red |
| 4734 | NEDD4 | red |
| 3750 | KCND1 | red |
| 10121 | ACTR1A | red |
| 6883 | TAF12 | red |
| 1284 | COL4A2 | red |
| 54809 | SAMD9 | red |
| 389 | RHOC | red |
| 9592 | IER2 | red |
| 3142 | HLX | red |
| 55119 | PRPF38B | red |
| 746 | TMEM258 | red |
| 54861 | SNRK | red |
| 3659 | IRF1 | red |
| 5138 | PDE2A | red |
| 55740 | ENAH | red |
| 5305 | PIP4K2A | red |
| 8905 | AP1S2 | red |
| 1012 | CDH13 | red |
| 5216 | PFN1 | red |
| 1847 | DUSP5 | red |

| | | |
|---|---|---|
| 10656 | KHDRBS3 | red |
| 3956 | LGALS1 | red |
| 80025 | PANK2 | red |
| 5082 | PDCL | red |
| 23266 | ADGRL2 | red |
| 55003 | PAK1IP1 | red |
| 9462 | RASAL2 | red |
| 837 | CASP4 | red |
| 51373 | MRPS17 | red |
| 10252 | SPRY1 | red |
| 64091 | POPDC2 | red |
| 1942 | EFNA1 | red |
| 7318 | UBA7 | red |
| 5696 | PSMB8 | red |
| 56603 | CYP26B1 | red |
| 2200 | FBN1 | red |
| 4521 | NUDT1 | red |
| 79720 | VPS37B | red |
| 56000 | NXF3 | red |
| 5947 | RBP1 | red |
| 8459 | TPST2 | red |
| 7846 | TUBA1A | red |
| 51162 | EGFL7 | red |
| 1306 | COL15A1 | red |
| 4296 | MAP3K11 | red |
| 6275 | S100A4 | red |
| 51690 | LSM7 | red |
| 116496 | FAM129A | red |
| 8612 | PPAP2C | red |
| 8764 | TNFRSF14 | red |
| 65998 | C11orf95 | red |
| 10985 | GCN1L1 | red |
| 10403 | NDC80 | red |
| 5321 | PLA2G4A | red |
| 9457 | FHL5 | red |
| 3908 | LAMA2 | red |
| 23545 | ATP6V0A2 | red |
| 2625 | GATA3 | red |
| 55281 | TMEM140 | red |
| 10769 | PLK2 | red |
| 2826 | CCR10 | red |

| | | |
|---|---|---|
| 2669 | GEM | red |
| 8925 | HERC1 | red |
| 64321 | SOX17 | red |
| 283298 | OLFML1 | red |
| 11000 | SLC27A3 | red |
| 22913 | RALY | red |
| 23336 | SYNM | red |
| 51550 | CINP | red |
| 10234 | LRRC17 | red |
| 81578 | COL21A1 | red |
| 8406 | SRPX | red |
| 2620 | GAS2 | red |
| 8525 | DGKZ | red |
| 5376 | PMP22 | red |
| 3117 | HLA-DQA1 | red |
| 5414 | 4-Sep | red |
| 1277 | COL1A1 | salmon |
| 9332 | CD163 | salmon |
| 1591 | CYP24A1 | salmon |
| 7058 | THBS2 | salmon |
| 4837 | NNMT | salmon |
| 7462 | LAT2 | salmon |
| 55165 | CEP55 | salmon |
| 2219 | FCN1 | salmon |
| 719 | C3AR1 | salmon |
| 11326 | VSIG4 | salmon |
| 2268 | FGR | salmon |
| 6351 | CCL4 | salmon |
| 6039 | RNASE6 | salmon |
| 712 | C1QA | salmon |
| 4069 | LYZ | salmon |
| 11151 | CORO1A | salmon |
| 3553 | IL1B | salmon |
| 91543 | RSAD2 | salmon |
| 7305 | TYROBP | salmon |
| 9582 | APOBEC3B | salmon |
| 6347 | CCL2 | salmon |
| 9636 | ISG15 | salmon |
| 79019 | CENPM | salmon |
| 64581 | CLEC7A | salmon |
| 2537 | IFI6 | salmon |

| | | |
|---|---|---|
| 10437 | IFI30 | salmon |
| 2123 | EVI2A | salmon |
| 64231 | MS4A6A | salmon |
| 2207 | FCER1G | salmon |
| 3872 | KRT17 | salmon |
| 2215 | FCGR3B | salmon |
| 728 | C5AR1 | salmon |
| 3689 | ITGB2 | salmon |
| 5341 | PLEK | salmon |
| 9450 | LY86 | salmon |
| 1278 | COL1A2 | salmon |
| 83716 | CRISPLD2 | salmon |
| 9235 | IL32 | salmon |
| 1475 | CSTA | salmon |
| 53829 | P2RY13 | salmon |
| 7409 | VAV1 | salmon |
| 10874 | NMU | salmon |
| 56253 | CRTAM | salmon |
| 963 | CD53 | salmon |
| 4599 | MX1 | salmon |
| 695 | BTK | salmon |
| 9055 | PRC1 | salmon |
| 5199 | CFP | salmon |
| 4064 | CD180 | salmon |
| 64127 | NOD2 | salmon |
| 717 | C2 | salmon |
| 51316 | PLAC8 | salmon |
| 1959 | EGR2 | salmon |
| 597 | BCL2A1 | salmon |
| 24137 | KIF4A | salmon |
| 1675 | CFD | salmon |
| 1536 | CYBB | salmon |
| 962 | CD48 | salmon |
| 9595 | CYTIP | salmon |
| 55247 | NEIL3 | salmon |
| 57214 | CEMIP | salmon |
| 3669 | ISG20 | salmon |
| 10993 | SDS | salmon |
| 2357 | FPR1 | salmon |
| 7096 | TLR1 | salmon |
| 10346 | TRIM22 | salmon |

| | | |
|---|---|---|
| 241 | ALOX5AP | salmon |
| 6614 | SIGLEC1 | salmon |
| 409 | ARRB2 | salmon |
| 51311 | TLR8 | salmon |
| 9051 | PSTPIP1 | salmon |
| 4688 | NCF2 | salmon |
| 4938 | OAS1 | salmon |
| 5831 | PYCR1 | salmon |
| 50856 | CLEC4A | salmon |
| 2335 | FN1 | salmon |
| 752 | FMNL1 | salmon |
| 79968 | WDR76 | salmon |
| 4600 | MX2 | salmon |
| 3903 | LAIR1 | salmon |
| 54739 | XAF1 | salmon |
| 7538 | ZFP36 | salmon |
| 6283 | S100A12 | salmon |
| 28231 | SLCO4A1 | salmon |
| 1520 | CTSS | salmon |
| 2921 | CXCL3 | salmon |
| 4689 | NCF4 | salmon |
| 2205 | FCER1A | salmon |
| 2650 | GCNT1 | salmon |
| 1164 | CKS2 | salmon |
| 7133 | TNFRSF1B | salmon |
| 4050 | LTB | salmon |
| 6688 | SPI1 | salmon |
| 55911 | APOBR | salmon |
| 284021 | MILR1 | salmon |
| 6402 | SELL | salmon |
| 3702 | ITK | salmon |
| 51284 | TLR7 | salmon |
| 5359 | PLSCR1 | salmon |
| 6648 | SOD2 | salmon |
| 5788 | PTPRC | salmon |
| 51411 | BIN2 | salmon |
| 10288 | LILRB2 | salmon |
| 79444 | BIRC7 | salmon |
| 11010 | GLIPR1 | salmon |
| 3936 | LCP1 | salmon |
| 6280 | S100A9 | salmon |

| | | |
|---|---|---|
| 57823 | SLAMF7 | salmon |
| 11314 | CD300A | salmon |
| 7940 | LST1 | salmon |
| 10333 | TLR6 | salmon |
| 969 | CD69 | salmon |
| 6404 | SELPLG | salmon |
| 9770 | RASSF2 | salmon |
| 4046 | LSP1 | salmon |
| 4542 | MYO1F | salmon |
| 3665 | IRF7 | salmon |
| 64108 | RTP4 | salmon |
| 3732 | CD82 | salmon |
| 3684 | ITGAM | salmon |
| 4038 | LRP4 | salmon |
| 4332 | MNDA | salmon |
| 2124 | EVI2B | salmon |
| 3587 | IL10RA | salmon |
| 27128 | CYTH4 | salmon |
| 5978 | REST | salmon |
| 3687 | ITGAX | salmon |
| 390411 | NPM1P22 | salmon |
| 3055 | HCK | salmon |
| 5366 | PMAIP1 | salmon |
| 2533 | FYB | salmon |
| 915 | CD3D | salmon |
| 64135 | IFIH1 | salmon |
| 3394 | IRF8 | salmon |
| 10561 | IFI44 | salmon |
| 970 | CD70 | salmon |
| 51611 | DPH5 | salmon |
| 6286 | S100P | salmon |
| 9046 | DOK2 | salmon |
| 11245 | GPR176 | salmon |
| 8519 | IFITM1 | salmon |
| 55379 | LRRC59 | salmon |
| 2863 | GPR39 | salmon |
| 3965 | LGALS9 | salmon |
| 11262 | SP140 | salmon |
| 945 | CD33 | salmon |
| 5265 | SERPINA1 | salmon |
| 3560 | IL2RB | salmon |

| | | |
|---|---|---|
| 5794 | PTPRH | salmon |
| 1441 | CSF3R | salmon |
| 7454 | WAS | salmon |
| 10475 | TRIM38 | salmon |
| 28954 | REM1 | salmon |
| 3159 | HMGA1 | salmon |
| 6398 | SECTM1 | salmon |
| 3676 | ITGA4 | salmon |
| 55008 | HERC6 | salmon |
| 23092 | ARHGAP26 | salmon |
| 9242 | MSC | salmon |
| 9900 | SV2A | salmon |
| 5610 | EIF2AK2 | salmon |
| 11219 | TREX2 | salmon |
| 28639 | TRBC1 | salmon |
| 1436 | CSF1R | salmon |
| 7351 | UCP2 | salmon |
| 4940 | OAS3 | salmon |
| 5097 | PCDH1 | salmon |
| 899 | CCNF | salmon |
| 23219 | FBXO28 | salmon |
| 25830 | SULT4A1 | salmon |
| 10261 | IGSF6 | salmon |
| 7456 | WIPF1 | salmon |
| 2253 | FGF8 | salmon |
| 586 | BCAT1 | salmon |
| 3431 | SP110 | salmon |
| 3437 | IFIT3 | salmon |
| 4074 | M6PR | salmon |
| 861 | RUNX1 | salmon |
| 7086 | TKT | salmon |
| 11027 | LILRA2 | salmon |
| 1871 | E2F3 | salmon |
| 8819 | SAP30 | salmon |
| 9111 | NMI | salmon |
| 8698 | S1PR4 | salmon |
| 27087 | B3GAT1 | salmon |
| 57089 | ENTPD7 | salmon |
| 54815 | GATAD2A | salmon |
| 9517 | SPTLC2 | salmon |
| 3779 | KCNMB1 | salmon |

| | | |
|---|---|---|
| 56905 | C15orf39 | salmon |
| 10211 | FLOT1 | salmon |
| 79865 | TREML2 | salmon |
| 7179 | TPTE | salmon |
| 27023 | FOXB1 | salmon |
| 4067 | LYN | salmon |
| 8477 | GPR65 | salmon |
| 79845 | RNF122 | salmon |
| 6891 | TAP2 | salmon |
| 6556 | SLC11A1 | salmon |
| 54974 | THG1L | salmon |
| 81030 | ZBP1 | salmon |
| 9473 | THEMIS2 | salmon |
| 3068 | HDGF | salmon |
| 116372 | LYPD1 | salmon |
| 1524 | CX3CR1 | salmon |
| 389856 | USP27X | salmon |
| 6662 | SOX9 | salmon |
| 5471 | PPAT | salmon |
| 64919 | BCL11B | salmon |
| 729230 | CCR2 | salmon |
| 6274 | S100A3 | salmon |
| 9957 | HS3ST1 | salmon |
| 7124 | TNF | salmon |
| 24138 | IFIT5 | salmon |
| 54149 | C21orf91 | salmon |
| 10570 | DPYSL4 | salmon |
| 8654 | PDE5A | salmon |
| 3071 | NCKAP1L | salmon |
| 4068 | SH2D1A | salmon |
| 5949 | RBP3 | salmon |
| 57128 | LYRM4 | salmon |
| 79723 | SUV39H2 | salmon |
| 960 | CD44 | salmon |
| 23368 | PPP1R13B | salmon |
| 6737 | TRIM21 | salmon |
| 1043 | CD52 | salmon |
| 4939 | OAS2 | salmon |
| 6455 | SH3GL1 | salmon |
| 1521 | CTSW | salmon |
| 7703 | PCGF2 | salmon |

| | | |
|---|---|---|
| 79149 | ZSCAN5A | salmon |
| 1058 | CENPA | salmon |
| 4153 | MBL2 | salmon |
| 80380 | PDCD1LG2 | salmon |
| 3037 | HAS2 | salmon |
| 6199 | RPS6KB2 | salmon |
| 2915 | GRM5 | salmon |
| 8530 | CST7 | salmon |
| 951 | CD37 | salmon |
| 51806 | CALML5 | salmon |
| 9447 | AIM2 | salmon |
| 2848 | GPR25 | salmon |
| 6732 | SRPK1 | salmon |
| 221692 | PHACTR1 | salmon |
| 10379 | IRF9 | salmon |
| 921 | CD5 | salmon |
| 22821 | RASA3 | salmon |
| 8828 | NRP2 | salmon |
| 672 | BRCA1 | salmon |
| 9050 | PSTPIP2 | salmon |
| 3340 | NDST1 | salmon |
| 5880 | RAC2 | salmon |
| 51154 | MRTO4 | salmon |
| 7517 | XRCC3 | salmon |
| 3002 | GZMB | salmon |
| 3939 | LDHA | salmon |
| 29992 | PILRA | salmon |
| 90861 | HN1L | salmon |
| 3932 | LCK | salmon |
| 10849 | CD3EAP | salmon |
| 27040 | LAT | salmon |
| 2920 | CXCL2 | salmon |
| 998 | CDC42 | salmon |
| 3683 | ITGAL | salmon |
| 27071 | DAPP1 | salmon |
| 140801 | RPL10L | salmon |
| 54440 | SASH3 | salmon |
| 3980 | LIG3 | salmon |
| 3635 | INPP5D | salmon |
| 5292 | PIM1 | salmon |
| 5721 | PSME2 | salmon |

| | | |
|---|---|---|
| 8048 | CSRP3 | salmon |
| 26099 | SZRD1 | salmon |
| 254102 | EHBP1L1 | salmon |
| 5999 | RGS4 | salmon |
| 433 | ASGR2 | salmon |
| 56993 | TOMM22 | salmon |
| 713 | C1QB | salmon |
| 5317 | PKP1 | salmon |
| 6515 | SLC2A3 | salmon |
| 653639 | LYPLA2P1 | salmon |
| 7980 | TFPI2 | salmon |
| 1794 | DOCK2 | salmon |
| 6357 | CCL13 | salmon |
| 9143 | SYNGR3 | salmon |
| 5087 | PBX1 | salmon |
| 10347 | ABCA7 | salmon |
| 10578 | GNLY | salmon |
| 22914 | KLRK1 | salmon |
| 4920 | ROR2 | salmon |
| 8361 | HIST1H4F | salmon |
| 7851 | MALL | salmon |
| 5579 | PRKCB | salmon |
| 5581 | PRKCE | salmon |
| 9493 | KIF23 | salmon |
| 5433 | POLR2D | salmon |
| 3561 | IL2RG | salmon |
| 8540 | AGPS | salmon |
| 8407 | TAGLN2 | salmon |
| 10721 | POLQ | salmon |
| 7402 | UTRN | salmon |
| 10544 | PROCR | salmon |
| 925 | CD8A | salmon |
| 51657 | STYXL1 | salmon |
| 2526 | FUT4 | salmon |
| 4001 | LMNB1 | salmon |
| 28526 | TRDC | salmon |
| 4017 | LOXL2 | salmon |
| 4224 | MEP1A | salmon |
| 51338 | MS4A4A | salmon |
| 9734 | HDAC9 | salmon |
| 1437 | CSF2 | salmon |

| | | |
|---|---|---|
| 54976 | C20orf27 | salmon |
| 8869 | ST3GAL5 | salmon |
| 10326 | SIRPB1 | salmon |
| 3433 | IFIT2 | salmon |
| 919 | CD247 | salmon |
| 8348 | HIST1H2BO | salmon |
| 81285 | OR51E2 | salmon |
| 79180 | EFHD2 | salmon |
| 30817 | ADGRE2 | salmon |
| 1848 | DUSP6 | salmon |
| 23133 | PHF8 | salmon |
| 51571 | FAM49B | salmon |
| 9334 | B4GALT5 | salmon |
| 220134 | SKA1 | salmon |
| 6664 | SOX11 | salmon |
| 5551 | PRF1 | salmon |
| 64806 | IL25 | salmon |
| 2119 | ETV5 | salmon |
| 4818 | NKG7 | salmon |
| 7128 | TNFAIP3 | salmon |
| 7157 | TP53 | salmon |
| 284119 | PTRF | salmon |
| 9410 | SNRNP40 | salmon |
| 51365 | PLA1A | salmon |
| 3594 | IL12RB1 | salmon |
| 10123 | ARL4C | salmon |
| 5892 | RAD51D | salmon |
| 864 | RUNX3 | salmon |
| 4317 | MMP8 | salmon |
| 3489 | IGFBP6 | salmon |
| 6279 | S100A8 | salmon |
| 51348 | KLRF1 | salmon |
| 5970 | RELA | salmon |
| 2026 | ENO2 | salmon |
| 5266 | PI3 | salmon |
| 2161 | F12 | salmon |
| 2999 | GZMH | salmon |
| 5179 | PENK | salmon |
| 55140 | ELP3 | salmon |
| 5920 | RARRES3 | salmon |
| 56935 | SMCO4 | salmon |

| | | |
|---|---|---|
| 2841 | GPR18 | salmon |
| 3371 | TNC | salmon |
| 3043 | HBB | salmon |
| 64795 | RMND5A | salmon |
| 3012 | HIST1H2AE | salmon |
| 440738 | MAP1LC3C | salmon |
| 87 | ACTN1 | salmon |
| 80830 | APOL6 | salmon |
| 968 | CD68 | salmon |
| 3040 | HBA2 | salmon |
| 3001 | GZMA | salmon |
| 154 | ADRB2 | salmon |
| 4162 | MCAM | salmon |
| 6352 | CCL5 | salmon |
| 115123 | 3-Mar | salmon |

**Supplemental Table 6: List of 194 significantly associated pathways (p<0.05) from genes in the three HF modules**

| Ingenuity Canonical Pathways | -log(B-H p-value) | Molecules |
|---|---|---|
| Hepatic Fibrosis / Hepatic Stellate Cell Activation | 1.11E+01 | A2M,ACTA2,AGTR1,BAX,CCL2,CCL5,CD14,CD40,COL11A2,COL13A1,COL15A1,COL1A1,COL1A2,COL21A1,COL4A1,COL4A2,COL5A2,COL6A3,CXCL3,EDNRA,FAS,FGF2,FLT4,FN1,ICAM1,IFNAR2,IFNGR1,IFNGR2,IGF2,IGFBP4,IGFBP5,IL10RA,IL1B,IL1R1,IL1RL1,IL4R,KDR,LY96,MMP2,MYH11,PDGFB,PDGFD,PDGFRA,PGF,RELA,SMAD7,TGFB1,TGFB2,TGFBR2,TLR4,TNF,TNFRSF1B |
| Th1 and Th2 Activation Pathway | 1.05E+01 | CD247,CD3D,CD4,CD40,CD8A,CXCR4,GATA3,HLA-DMA,HLA-DMB,HLA-DOA,HLA-DPA1,HLA-DPB1,HLA-DQA1,HLA-DQB1,HLA-DRA,HLA-DRB1,ICAM1,IFNGR1,IFNGR2,IL10RA,IL12A,IL12RB1,IL1RL1,IL25,IL27RA,IL2RB,IL2RG,IL4R,IRF1,ITGB2,JAG2,JUN,LGALS9,NFATC4,NOTCH1,NOTCH3,NOTCH4,PIK3C2B,PIK3R3,RUNX3,S1PR1,SPI1,TGFB1,TGFBR2,TNFRSF4,VAV1 |
| Role of Pattern Recognition Receptors in Recognition of Bacteria and Viruses | 9.79E+00 | C1QA,C1QB,C3AR1,C5AR1,CASP1,CCL5,CD70,CLEC7A,CSF2,EIF2AK2,EIF2S1,IFIH1,IL12A,IL17B,IL1B,IL25,IRF7,LTB,MAPK12,MBL2,MYD88,NOD2,OAS1,OAS2,OAS3,PIK3C2B,PIK3R3,PRKCB,PRKCE,PRKCH,PRKD3,RELA,TGFB1,TGFB2,TLR1,TLR4,TLR6,TLR7,TLR8,TNF,TNFSF10,TNFSF12 |
| Th2 Pathway | 8.89E+00 | CD247,CD3D,CD4,CD40,CXCR4,GATA3,HLA-DMA,HLA-DMB,HLA-DOA,HLA-DPA1,HLA-DPB1,HLA-DQA1,HLA-DQB1,HLA-DRA,HLA-DRB1,ICAM1,IL12A,IL12RB1,IL1RL1,IL25,IL2RB,IL2RG,IL4R,ITGB2,JAG2,JUN,NOTCH1,NOTCH3,NOTCH4,PIK3C2B,PIK3R3,RUNX3,S1PR1,SPI1,TGFB1,TGFBR2,TNFRSF4,VAV1 |
| Systemic Lupus Erythematosus In B Cell Signaling Pathway | 8.49E+00 | BTK,CALML5,CCND1,CD40,CD5,CD70,CD79B,CSF2,CSK,FGR,HCK,IFIH1,IFIT2,IFIT3,IFNAR2,IFNGR1,IFNGR2,IL12A,IL17B,IL1B,IL25,INPP5D,INPP5K,IRF7,IRF9,ISG15,ISG20,JUN,LCK,LTB,LYN,MAP3K14,MAP4K4,MCL1,MRAS,MYC,MYD88,NFATC4,NRAS,PIK3C2B,PIK3R3,PLAAT4,PRKCB,PRKCE,PRKCH,PRKD3,RAC2,RALB,RAP2B,RELA,TGFB1,TGFB2,TLR7,TLR8,TNF,TNFSF10,TNFSF12,VAV1 |

| | | |
|---|---|---|
| Th1 Pathway | 8.16E+00 | CD247,CD3D,CD4,CD40,CD8A,GATA3,HLA-DMA,HLA-DMB,HLA-DOA,HLA-DPA1,HLA-DPB1,HLA-DQA1,HLA-DQB1,HLA-DRA,HLA-DRB1,ICAM1,IFNGR1,IFNGR2,IL10RA,IL12A,IL12RB1,IL27RA,IRF1,ITGB2,LGALS9,NFATC4,NOTCH1,NOTCH3,NOTCH4,PIK3C2B,PIK3R3,RUNX3,VAV1 |
| Type I Diabetes Mellitus Signaling | 8.09E+00 | CASP8,CD247,CD3D,FAS,FCER1G,GZMB,HLA-DMA,HLA-DMB,HLA-DOA,HLA-DQA1,HLA-DQB1,HLA-DRA,HLA-DRB1,HLA-E,ICA1,IFNGR1,IFNGR2,IKBKB,IL12A,IL1B,IL1R1,IRF1,MAP3K14,MAPK11,MAPK12,MAPK14,MYD88,NFKBIE,PRF1,RELA,SOCS2,TNF,TNFRSF1B |
| iCOS-iCOSL Signaling in T Helper Cells | 7.45E+00 | CALML5,CD247,CD3D,CD4,CD40,CSK,FCER1G,HLA-DMA,HLA-DMB,HLA-DOA,HLA-DQA1,HLA-DQB1,HLA-DRA,HLA-DRB1,IKBKB,IL2RB,IL2RG,INPP5D,ITK,ITPR3,LAT,LCK,LCP2,NFATC4,NFKBIE,PIK3C2B,PIK3R3,PTEN,PTPRC,RELA,VAV1 |
| **Interferon Signaling** | 7.33E+00 | BAK1,BAX,IFI35,IFI6,IFIT3,IFITM1,IFITM3,IFNAR2,IFNGR1,IFNGR2,IRF1,IRF9,ISG15,MX1,OAS1,PSMB8,RELA,TAP1 |
| CD28 Signaling in T Helper Cells | 6.98E+00 | ARPC1B,ARPC2,CALML5,CD247,CD3D,CD4,CDC42,CSK,FCER1G,HLA-DMA,HLA-DMB,HLA-DOA,HLA-DQA1,HLA-DQB1,HLA-DRA,HLA-DRB1,IKBKB,ITK,ITPR3,JUN,LAT,LCK,LCP2,MAPK12,NFATC4,NFKBIE,PIK3C2B,PIK3R3,PTPRC,RELA,VAV1,WAS |
| Role of Macrophages, Fibroblasts and Endothelial Cells in Rheumatoid Arthritis | 6.80E+00 | C5AR1,CALML5,CCL2,CCL5,CCND1,CEBPA,CEBPB,CEBPG,CSF2,DKK1,DKK3,FCGR3A/FCGR3B,FGF2,FN1,FRZB,FZD4,ICAM1,IKBKB,IL16,IL1B,IL1R1,IL1RL1,IL32,IL7,JUN,LTB,MAP3K14,MAPK14,MMP3,MRAS,MYC,MYD88,NFATC4,NFKBIE,NRAS,PDGFB,PDGFD,PGF,PIK3C2B,PIK3R3,PRKCB,PRKCE,PRKCH,PRKD3,RALB,RAP2B,RELA,ROR2,TCF3,TGFB1,TLR1,TLR4,TLR6,TLR7,TLR8,TNF,TNFRSF1B,WNT5B |
| Dendritic Cell Maturation | 6.33E+00 | CD1D,CD40,CD58,COL11A2,COL1A1,COL1A2,CSF2,FCER1G,FCGR3A/FCGR3B,FSCN1,HLA-DMA,HLA-DMB,HLA-DOA,HLA-DQA1,HLA-DQB1,HLA-DRA,HLA-DRB1,HLA-E,ICAM1,IKBKB,IL12A,IL1B,IL32,IRF8,LTB,LY75,MAP3K14,MAPK11,MAPK12,MAPK14,MYD88,NFKBIE,PIK3C2B,PIK3R3,RELA,TLR4,TNF,TNFRSF1B,TYROBP |

| | | |
|---|---|---|
| OX40 Signaling Pathway | 6.24E+00 | CD247,CD3D,CD4,FCER1G,HLA-DMA,HLA-DMB,HLA-DOA,HLA-DPA1,HLA-DPB1,HLA-DQA1,HLA-DQB1,HLA-DRA,HLA-DRB1,HLA-E,JUN,MAPK12,NFKBIE,RELA,TNFRSF4 |
| **Tec Kinase Signaling** | 6.24E+00 | ACTA2,ACTC1,BLK,BTK,CDC42,FAS,FCER1A,FCER1G,FGR,GNA14,GNG11,GNG7,HCK,ITGA4,ITGA5,ITK,LCK,LYN,MAPK12,MRAS,PIK3C2B,PIK3R3,PRKCB,PRKCE,PRKCH,PRKD3,RAC2,RELA,RHOBTB1,RHOC,RND3,TLR4,TNF,TNFRSF10B,TNFSF10,TNFSF12,VAV1,WAS |
| PD-1, PD-L1 cancer immunotherapy pathway | 6.24E+00 | CD247,CSK,HLA-DMA,HLA-DMB,HLA-DOA,HLA-DPA1,HLA-DPB1,HLA-DQA1,HLA-DQB1,HLA-DRA,HLA-DRB1,HLA-E,IFNGR1,IFNGR2,IL12A,IL2RB,IL2RG,LAT,LCK,LCP2,PDCD1LG2,PIK3C2B,PIK3R3,PTEN,TGFB1,TGFB2,TNF,TNFRSF1B |
| Neuroinflammation Signaling Pathway | 6.06E+00 | BIRC5,BIRC7,CASP1,CASP8,CCL2,CCL5,CD40,CSF1R,CX3CL1,CX3CR1,CYBB,FAS,HLA-DMA,HLA-DMB,HLA-DOA,HLA-DQA1,HLA-DQB1,HLA-DRA,HLA-DRB1,HLA-E,ICAM1,IFNGR1,IFNGR2,IKBKB,IL12A,IL1B,IL1R1,IRF7,JUN,MAPK11,MAPK12,MAPK14,MFGE8,MMP3,MYD88,NCF2,NFATC4,PIK3C2B,PIK3R3,PLA2G4A,PLA2G4C,PYCARD,RELA,SLC1A3,SOD2,TGFB1,TGFB2,TGFBR2,TLR1,TLR4,TLR6,TLR7,TLR8,TNF,TYROBP |
| T Helper Cell Differentiation | 6.05E+00 | BCL6,CD40,FCER1G,GATA3,HLA-DMA,HLA-DMB,HLA-DOA,HLA-DQA1,HLA-DQB1,HLA-DRA,HLA-DRB1,IFNGR1,IFNGR2,IL10RA,IL12A,IL12RB1,IL2RG,IL4R,TGFB1,TGFBR2,TNF,TNFRSF1B |
| Altered T Cell and B Cell Signaling in Rheumatoid Arthritis | 6.05E+00 | CD40,CD79B,CSF2,FAS,FCER1G,HLA-DMA,HLA-DMB,HLA-DOA,HLA-DQA1,HLA-DQB1,HLA-DRA,HLA-DRB1,IL12A,IL1B,LTB,MAP3K14,RELA,TGFB1,TLR1,TLR4,TLR6,TLR7,TLR8,TNF |
| TREM1 Signaling | 5.94E+00 | CASP1,CCL2,CD40,CSF2,CXCL3,ICAM1,IL1B,IL1RL1,ITGA5,ITGAX,LAT2,MYD88,NOD2,RELA,TLR1,TLR4,TLR6,TLR7,TLR8,TNF,TYROBP |
| Phagosome Formation | 5.89E+00 | C3AR1,C5AR1,CDC42,CLEC7A,FCER1A,FCER1G,FCGR3A/FCGR3B,FN1,ITGA4,ITGA5,ITGAM,ITGAX,ITGB2,MBL2,PIK3C2B,PIK3R3,PRKCB,PRKCE,PRKCH,PRKD3,RAC2,RHOBTB1,RHOC,RND3,TLR1,TLR4,TLR6,TLR7,TLR8 |

| | | |
|---|---|---|
| Leukocyte Extravasation Signaling | 5.87E+00 | ACTA2,ACTC1,ACTN1,AFDN,ARHGAP4,BTK,CD44,CDC42,CDH5,CXCR4,CYBA,CYBB,ICAM1,ITGA4,ITGA5,ITGAL,ITGAM,ITGB2,ITK,JAM2,MAPK11,MAPK12,MAPK14,MMP2,MMP3,MMP8,NCF2,NCF4,PECAM1,PIK3C2B,PIK3R3,PRKCB,PRKCE,PRKCH,PRKD3,RAC2,SELPLG,SIPA1,VASP,VAV1,WAS,WIPF1 |
| HMGB1 Signaling | 5.87E+00 | CCL2,CD70,CDC42,CSF2,ICAM1,IFNGR1,IFNGR2,IL12A,IL17B,IL1B,IL1R1,IL25,JUN,LTB,MAPK11,MAPK12,MAPK14,MRAS,NRAS,PIK3C2B,PIK3R3,PLAT,RAC2,RALB,RAP2B,RELA,RHOBTB1,RHOC,RND3,TGFB1,TGFB2,TLR4,TNF,TNFRSF1B,TNFSF10,TNFSF12 |
| Role of NFAT in Regulation of the Immune Response | 5.63E+00 | BTK,CALML5,CD247,CD3D,CD4,CD79B,FCER1A,FCER1G,FCGR3A/FCGR3B,GNA14,GNG11,GNG7,HLA-DMA,HLA-DMB,HLA-DOA,HLA-DQA1,HLA-DQB1,HLA-DRA,HLA-DRB1,IKBKB,ITK,ITPR3,JUN,LAT,LCK,LCP2,LYN,MEF2C,MRAS,NFATC4,NFKBIE,NRAS,PIK3C2B,PIK3R3,RALB,RAP2B,RCAN2,RELA |
| **Antigen Presentation Pathway** | 5.63E+00 | CD74,HLA-DMA,HLA-DMB,HLA-DOA,HLA-DPA1,HLA-DPB1,HLA-DQA1,HLA-DQB1,HLA-DRA,HLA-DRB1,HLA-E,PSMB8,PSMB9,TAP1,TAP2 |
| Calcium-induced T Lymphocyte Apoptosis | 5.52E+00 | ATP2A3,CALML5,CD247,CD3D,CD4,FCER1G,HLA-DMA,HLA-DMB,HLA-DOA,HLA-DQA1,HLA-DQB1,HLA-DRA,HLA-DRB1,ITPR3,LCK,PRKCB,PRKCE,PRKCH,PRKD3 |
| Allograft Rejection Signaling | 5.52E+00 | CD40,FAS,FCER1G,GZMB,HLA-DMA,HLA-DMB,HLA-DOA,HLA-DPA1,HLA-DPB1,HLA-DQA1,HLA-DQB1,HLA-DRA,HLA-DRB1,HLA-E,PRF1,TNF |
| Toll-like Receptor Signaling | 5.52E+00 | CD14,EIF2AK2,IKBKB,IL12A,IL1B,IL1RL1,JUN,LY96,MAP3K14,MAP4K4,MAPK11,MAPK12,MAPK14,MYD88,RELA,TLR1,TLR4,TLR6,TLR7,TLR8,TNF,TNFAIP3 |
| Crosstalk between Dendritic Cells and Natural Killer Cells | 5.45E+00 | ACTA2,ACTC1,CD40,CD69,CSF2,FAS,FSCN1,HLA-DRA,HLA-DRB1,HLA-E,IL12A,IL15RA,IL2RB,IL2RG,ITGAL,KLRC4-KLRK1/KLRK1,LTB,PRF1,RELA,TLR4,TLR7,TNF,TNFRSF1B,TNFSF10,TYROBP |
| Cdc42 Signaling | 5.35E+00 | ARHGEF6,ARPC1B,ARPC2,CD247,CD3D,CDC42,CDC42SE1,FCER1G,FGD1,HLA-DMA,HLA-DMB,HLA-DOA,HLA-DPA1,HLA-DPB1,HLA-DQA1,HLA-DQB1,HLA-DRA,HLA-DRB1,HLA-E,ITGA4,ITGA5,ITK,JUN,MAP3K11,MAPK11,MAPK12,MAPK14,VAV1,WAS,WIPF1 |

| | | |
|---|---|---|
| Phospholipase C Signaling | 4.97E+00 | ADCY1,ADCY3,ARHGEF15,ARHGEF2,ARHGEF6,ARHGEF7,BTK,CALML5,CD247,CD3D,CD79B,CDC42,FCER1G,GNG11,GNG7,HDAC7,HDAC9,ITGA4,ITGA5,ITK,ITPR3,LAT,LCK,LCP2,LYN,MEF2C,MRAS,NFATC4,NRAS,PLA2G4A,PLA2G4C,PRKCB,PRKCE,PRKCH,PRKD3,RAC2,RALB,RAP2B,RELA,RHOBTB1,RHOC,RND3,RPS6KA3,TGM2 |
| FcŒ≥ Receptor-mediated Phagocytosis in Macrophages and Monocytes | 4.96E+00 | ACTA2,ACTC1,ARPC1B,ARPC2,CDC42,CSF2,FCGR3A/FCGR3B,FGR,FYB1,HCK,INPP5D,LCP2,LYN,NCK1,PIK3R3,PRKCB,PRKCE,PRKCH,PRKD3,PTEN,RAC2,RPS6KB2,VASP,VAV1,WAS |
| iNOS Signaling | 4.90E+00 | CALML5,CD14,HMGA1,IFNGR1,IFNGR2,IKBKB,IRF1,JUN,LY96,MAPK11,MAPK12,MAPK14,MYD88,NFKBIE,RELA,TLR4 |
| IL-8 Signaling | 4.80E+00 | ARRB2,BAX,CCND1,CDC42,CSTB,CXCR2,CYBB,FLT4,GNG11,GNG7,ICAM1,IKBKB,ITGAM,ITGAX,ITGB2,JUN,KDR,MAP4K4,MAPK12,MMP2,MRAS,NCF2,NRAS,PGF,PIK3C2B,PIK3R3,PRKCB,PRKCE,PRKCH,PRKD3,RAC2,RALB,RAP2B,RELA,RHOBTB1,RHOC,RND3,TEK,VASP |
| Granulocyte Adhesion and Diapedesis | 4.80E+00 | C5AR1,CCL13,CCL18,CCL2,CCL4,CCL5,CDH5,CKLF,CSF3R,CX3CL1,CXCL2,CXCL3,CXCR2,CXCR4,FPR1,ICAM1,ICAM2,IL1B,IL1R1,IL1RL1,ITGA4,ITGA5,ITGAL,ITGAM,ITGB2,MMP2,MMP3,MMP8,PECAM1,SELL,SELP,SELPLG,TNF,TNFRSF1B |
| B Cell Receptor Signaling | 4.80E+00 | BCL2A1,BCL6,BTK,CALML5,CD79B,CDC42,CSK,DAPP1,ETS1,IKBKB,INPP5D,INPP5K,JUN,LYN,MAP3K11,MAP3K14,MAPK11,MAPK12,MAPK14,MEF2C,MRAS,NFATC4,NFKBIE,NRAS,PIK3C2B,PIK3R3,PRKCB,PTEN,PTPRC,RAC2,RALB,RAP2B,RELA,RPS6KB2,TCF3,VAV1 |
| T Cell Exhaustion Signaling Pathway | 4.80E+00 | BCL6,FCER1G,GZMB,HLA-DMA,HLA-DMB,HLA-DOA,HLA-DPA1,HLA-DPB1,HLA-DQA1,HLA-DQB1,HLA-DRA,HLA-DRB1,HLA-E,IFNAR2,IL10RA,IL12A,IL12RB1,IRF9,JUN,KDR,LGALS9,MAPK12,MRAS,NFATC4,NRAS,PDCD1LG2,PIK3C2B,PIK3R3,PRDM1,RALB,RAP2B,TGFB1,TGFBR2,TNFRSF14 |
| Hepatic Cholestasis | 4.68E+00 | ABCC1,ADCY1,ADCY3,CD14,CD70,CETP,CSF2,IKBKB,IL12A,IL17B,IL1B,IL1R1,IL1RL1,IL25,JUN,LTB,LY96,MAP3K14,MAPK12,MYD88,NFKBIE,NR5A2,PRKCB,PRKCE,PRKCH,PRKD3,RELA,RXRA,SLCO3A1,TGFB1,TGFB2,TLR4,TNF,TNFRSF1B,TNFSF10,TNFSF12 |

| | | |
|---|---|---|
| **STAT3 Pathway** | 4.63E+00 | CXCR2,FGF2,FLT4,IL10RA,IL12RB1,IL15RA,IL1B,IL1R1,IL1RL1,IL27RA,IL2RB,IL2RG,IL4R,KDR,MAP3K11,MAPK11,MAPK12,MAPK14,MRAS,MYC,NRAS,PDGFB,PDGFRA,PIM1,RALB,RAP2B,SOCS2,TGFB1,TGFB2,TGFBR2 |
| Cardiac Hypertrophy Signaling (Enhanced) | 4.62E+00 | ADCY1,ADCY3,ADRB2,AGTR1,ATP2A3,CACNA1C,CALML5,CD70,CSF2,CXCR2,CYBB,EDNRA,EIF2B4,FGF2,FGF8,FZD4,GNA14,GNG11,GNG7,HDAC7,HDAC9,HSPB3,IKBKB,IL10RA,IL12A,IL12RB1,IL15RA,IL17B,IL1B,IL1R1,IL1RL1,IL25,IL27RA,IL2RB,IL2RG,IL4R,ITGA4,ITGA5,ITPR3,JUN,LTB,MAP3K11,MAP3K14,MAPK11,MAPK12,MAPK14,MEF2C,MRAS,MYC,NFATC4,NRAS,PDE2A,PDE5A,PIK3C2B,PIK3R3,PRKCB,PRKCE,PRKCH,PRKD3,PTEN,RALB,RAP2B,RELA,RPS6KB2,TGFB1,TGFB2,TGFBR2,TNF,TNFRSF1B,TNFSF10,TNFSF12,WNT5B |
| Atherosclerosis Signaling | 4.61E+00 | CCL2,CCR2,CD40,COL11A2,COL1A1,COL1A2,CXCR4,ICAM1,IL1B,ITGA4,ITGB2,LYZ,MMP3,PDGFB,PDGFD,PLA2G4A,PLA2G4C,PLA2G7,PLAAT4,RELA,S100A8,SELP,SELPLG,SERPINA1,TGFB1,TNF,TNFRSF14,TNFSF12 |
| NF-ŒʃB Activation by Viruses | 4.55E+00 | CD4,EIF2AK2,IKBKB,ITGA4,ITGA5,ITGAL,ITGB2,LCK,MAP3K14,MRAS,NFKBIE,NRAS,PIK3C2B,PIK3R3,PRKCB,PRKCE,PRKCH,PRKD3,RALB,RAP2B,RELA,TNFRSF14 |
| Acute Phase Response Signaling | 4.48E+00 | A2M,C2,CEBPB,CP,F8,FN1,HAMP,HP,IKBKB,IL1B,IL1R1,JUN,MAP3K14,MAPK11,MAPK12,MAPK14,MBL2,MRAS,MYD88,NFKBIE,NRAS,OSMR,PIK3R3,RALB,RAP2B,RBP1,RBP3,RELA,SERPINA1,SERPINA3,SOCS2,SOD2,TCF3,TNF,TNFRSF1B,VWF |
| B Cell Development | 4.48E+00 | CD40,CD79B,HLA-DMA,HLA-DMB,HLA-DOA,HLA-DQA1,HLA-DQB1,HLA-DRA,HLA-DRB1,IL7,PTPRC,RAG1 |
| Systemic Lupus Erythematosus In T Cell Signaling Pathway | 4.46E+00 | BCL6,CASP1,CASP10,CASP4,CASP8,CD247,CD3D,CD44,CD70,CDC42,FAS,FCER1G,HLA-DMA,HLA-DMB,HLA-DOA,HLA-DPA1,HLA-DPB1,HLA-DQA1,HLA-DQB1,HLA-DRA,HLA-DRB1,HLA-E,ITGAL,JUN,LAT,MRAS,NOS3,NRAS,PIK3C2B,PIK3R3,RAC2,RALB,RAP2B,RHOBTB1,RHOC,RND3,RPS6KB2,SELPLG,VAV1 |
| Natural Killer Cell Signaling | 4.41E+00 | CD247,CD300A,FCER1G,FCGR3A/FCGR3B,INPP5D,INPP5K,KLRC4-KLRK1/KLRK1,LAIR1,LAT,LCK,LCP2,MRAS,NCK1,NRAS,PIK3C2B,PIK3R3,PRKCB,PRKCE,PRKCH,PRKD3,RAC2,RALB,RAP2B,SH2D1A,SIGLEC7,TYROBP,VAV1 |

| | | |
|---|---|---|
| Fc Epsilon RI Signaling | 4.41E+00 | BTK,CSF2,FCER1A,FCER1G,INPP5D,INPP5K,LAT,LCP2,LYN,MAPK11,MAPK12,MAPK14,MRAS,NRAS,PIK3C2B,PIK3R3,PLA2G4A,PLA2G4C,PRKCB,PRKCE,PRKCH,PRKD3,RAC2,RALB,RAP2B,TNF,VAV1 |
| Graft-versus-Host Disease Signaling | 4.34E+00 | FAS,FCER1G,GZMB,HLA-DMA,HLA-DMB,HLA-DOA,HLA-DQA1,HLA-DQB1,HLA-DRA,HLA-DRB1,HLA-E,IL1B,PRF1,TNF |
| Paxillin Signaling | 4.23E+00 | ACTA2,ACTC1,ACTN1,ARHGEF6,ARHGEF7,CDC42,CSK,ITGA10,ITGA4,ITGA5,ITGA8,ITGAL,ITGAM,ITGAX,ITGB2,MAPK11,MAPK12,MAPK14,MRAS,NCK1,NRAS,PIK3C2B,PIK3R3,RALB,RAP2B |
| PKCŒ∏ Signaling in T Lymphocytes | 4.21E+00 | CACNA1C,CD247,CD3D,CD4,FCER1G,HLA-DMA,HLA-DMB,HLA-DOA,HLA-DQA1,HLA-DQB1,HLA-DRA,HLA-DRB1,IKBKB,JUN,LAT,LCK,LCP2,MAP3K11,MAP3K14,MRAS,NFATC4,NFKBIE,NRAS,PIK3C2B,PIK3R3,RAC2,RALB,RAP2B,RELA,VAV1 |
| T Cell Receptor Signaling | 4.15E+00 | BTK,CALML5,CD247,CD3D,CD4,CD8A,CSK,IKBKB,ITK,JUN,LAT,LCK,LCP2,MRAS,NFATC4,NRAS,PIK3C2B,PIK3R3,PTPRC,PTPRH,RALB,RAP2B,RELA,VAV1 |
| LPS-stimulated MAPK Signaling | 4.07E+00 | CD14,CDC42,IKBKB,JUN,MAP3K14,MAPK11,MAPK12,MAPK14,MRAS,NFKBIE,NRAS,PIK3C2B,PIK3R3,PRKCB,PRKCE,PRKCH,PRKD3,RALB,RAP2B,RELA,TLR4 |
| IL-12 Signaling and Production in Macrophages | 3.99E+00 | CD40,CEBPB,IFNGR1,IKBKB,IL12A,IL12RB1,IRF1,IRF8,JUN,LYZ,MAPK11,MAPK12,MAPK14,MYD88,PIK3C2B,PIK3R3,PRKCB,PRKCE,PRKCH,PRKD3,RELA,RXRA,S100A8,SERPINA1,SPI1,TGFB1,TGFB2,TLR4,TNF |
| Autoimmune Thyroid Disease Signaling | 3.94E+00 | CD40,FAS,FCER1G,GZMB,HLA-DMA,HLA-DMB,HLA-DOA,HLA-DQA1,HLA-DQB1,HLA-DRA,HLA-DRB1,HLA-E,PRF1 |
| Agranulocyte Adhesion and Diapedesis | 3.81E+00 | ACTA2,ACTC1,C5AR1,CCL13,CCL18,CCL2,CCL4,CCL5,CD34,CDH5,CKLF,CX3CL1,CXCL2,CXCL3,CXCR2,CXCR4,FN1,ICAM1,ICAM2,IL1B,IL1R1,ITGA4,ITGA5,ITGB2,MMP2,MMP3,MMP8,MYH11,PECAM1,SELL,SELP,SELPLG,TNF |
| Production of Nitric Oxide and Reactive Oxygen Species in Macrophages | 3.70E+00 | CDC42,CYBA,CYBB,IFNGR1,IFNGR2,IKBKB,IRF1,IRF8,JUN,LYZ,MAP3K11,MAP3K14,MAPK11,MAPK12,MAPK14,NCF2,NCF4,NFKBIE,PIK3C2B,PIK3R3,PRKCB,PRKCE,PRKCH,PRKD3,RAC2,RELA,RHOBTB1,RHOC,RND3,S100A8,SERPINA1,SPI1,TLR4,TNF,TNFRSF1B |
| IL-4 Signaling | 3.65E+00 | HLA-DMA,HLA-DMB,HLA-DOA,HLA-DQA1,HLA-DQB1,HLA-DRA,HLA-DRB1,HMGA1,IL2RG,IL4R,INPP5D,INPP5K,MRAS,NFATC4,NRAS,PIK3C2B,PIK3R3,RALB,RAP2B,RPS6KB2 |

| | | |
|---|---|---|
| **Inflammasome pathway** | 3.62E+00 | AIM2,CASP1,CASP8,IL1B,MYD88,NLRP1,NOD2,PYCARD,TLR4 |
| Communication between Innate and Adaptive Immune Cells | 3.58E+00 | CCL4,CCL5,CD4,CD40,CD79B,CD8A,CSF2,FCER1G,HLA-DRA,HLA-DRB1,HLA-E,IL12A,IL1B,TLR1,TLR4,TLR6,TLR7,TLR8,TNF |
| Integrin Signaling | 3.58E+00 | ACTA2,ACTC1,ACTN1,ARHGAP26,ARHGEF7,ARPC1B,ARPC2,ASAP1,CDC42,ITGA10,ITGA4,ITGA5,ITGA8,ITGAL,ITGAM,ITGAX,ITGB2,MAP3K11,MRAS,NCK1,NRAS,PARVB,PDGFB,PFN1,PIK3C2B,PIK3R3,PTEN,RAC2,RALB,RAP2B,RHOBTB1,RHOC,RND3,TSPAN7,VASP,WAS,WIPF1 |
| GP6 Signaling Pathway | 3.51E+00 | BTK,CALML5,COL11A2,COL13A1,COL15A1,COL1A1,COL1A2,COL21A1,COL4A1,COL4A2,COL5A2,COL6A3,FCER1G,LAMA2,LAMA4,LAT,LCP2,LYN,PIK3C2B,PIK3R3,PRKCB,PRKCE,PRKCH,PRKD3 |
| Apoptosis Signaling | 3.46E+00 | BAK1,BAX,BCL2A1,CASP10,CASP8,CDK1,FAS,GAS2,IKBKB,MAP3K14,MAP4K4,MCL1,MRAS,NFKBIE,NRAS,PRKCE,RALB,RAP2B,RELA,TNF,TNFRSF1B,TP53 |
| NF-ŒʃB Signaling | 3.41E+00 | CASP8,CD40,EIF2AK2,FCER1G,FLT4,IKBKB,IL1B,IL1R1,KDR,LCK,MAP3K14,MAP4K4,MRAS,MYD88,NFKBIE,NRAS,PDGFRA,PIK3C2B,PIK3R3,PRKCB,RALB,RAP2B,RELA,TGFBR2,TLR1,TLR4,TLR6,TLR7,TLR8,TNF,TNFAIP3,TNFRSF1B |
| IL-6 Signaling | 3.40E+00 | A2M,CD14,CEBPB,COL1A1,HSPB3,IKBKB,IL1B,IL1R1,IL1RL1,JUN,MAP3K14,MAP4K4,MAPK11,MAPK12,MAPK14,MCL1,MRAS,NFKBIE,NRAS,PIK3C2B,PIK3R3,RALB,RAP2B,RELA,TNF,TNFRSF1B |
| Apelin Endothelial Signaling Pathway | 3.40E+00 | ADCY1,ADCY3,APLNR,CALML5,CCL2,ICAM1,JUN,KLF2,MAPK12,MEF2C,MRAS,NOS3,NRAS,PIK3C2B,PIK3R3,PRKCB,PRKCE,PRKCH,PRKD3,RALB,RAP2B,RELA,RPS6KB2,TEK |
| Colorectal Cancer Metastasis Signaling | 3.35E+00 | ADCY1,ADCY3,ARRB1,BAX,BIRC5,CCND1,CDC42,FZD4,GNG11,GNG7,IFNGR1,JUN,MAPK12,MMP2,MMP3,MMP8,MRAS,MYC,NRAS,PGF,PIK3C2B,PIK3R3,RAC2,RALB,RAP2B,RELA,RHOBTB1,RHOC,RND3,TCF3,TGFB1,TGFB2,TGFBR2,TLR1,TLR4,TLR6,TLR7,TLR8,TNF,TP53,WNT5B |
| fMLP Signaling in Neutrophils | 3.34E+00 | ARPC1B,ARPC2,CALML5,CDC42,CYBB,FPR1,GNG11,GNG7,ITPR3,MRAS,NCF2,NFATC4,NFKBIE,NRAS,PIK3C2B,PIK3R3,PRKCB,PRKCE,PRKCH,PRKD3,RALB,RAP2B,RELA,WAS |
| Primary Immunodeficiency Signaling | 3.34E+00 | ADA,BTK,CD3D,CD4,CD40,CD8A,IL2RG,LCK,PTPRC,RAG1,RFX5,TAP1,TAP2 |

| | | |
|---|---|---|
| HOTAIR Regulatory Pathway | 3.34E+00 | AR,ATG7,CD44,COL1A1,COL1A2,DNMT3B,EZH2,FOXM1,ICAM1,IRF1,JAM2,MMP2,MMP3,MMP8,MYC,PIK3C2B,PIK3R3,PTEN,RBBP4,RELA,REST,RHOC,SNAI2,TCF3,TGFB1,TLR4,TWIST1,WNT5B |
| Actin Nucleation by ARP-WASP Complex | 3.29E+00 | ARPC1B,ARPC2,CDC42,ITGA4,ITGA5,MRAS,NCK1,NRAS,RAC2,RALB,RAP2B,RHOBTB1,RHOC,RND3,VASP,WAS,WIPF1 |
| CCR5 Signaling in Macrophages | 3.24E+00 | CACNA1C,CALML5,CCL4,CCL5,CD247,CD3D,CD4,FAS,FCER1G,GNG11,GNG7,JUN,MAPK11,MAPK12,MAPK14,MRAS,PRKCB,PRKCE,PRKCH,PRKD3 |
| Osteoarthritis Pathway | 3.23E+00 | ACVRL1,CASP1,CASP10,CASP4,CASP8,CEBPB,CXCR2,DDIT4,DKK1,FGF2,FGF8,FN1,FRZB,FZD4,IL1B,IL1R1,IL1RL1,ITGA4,ITGA5,MEF2C,MMP3,NOTCH1,PGF,RELA,S100A8,S100A9,SMAD7,SMAD9,SOX9,TCF3,TGFB1,TGFBR2,TLR4,TNF,TNFRSF1B |
| Glioma Signaling | 3.20E+00 | CALML5,CAMK1,CAMK1D,CAMK1G,CCND1,CDKN2B,E2F3,IGF2,MRAS,NRAS,PDGFB,PDGFD,PDGFRA,PIK3C2B,PIK3R3,PRKCB,PRKCE,PRKCH,PRKD3,PTEN,RALB,RAP2B,TP53 |
| VDR/RXR Activation | 3.15E+00 | CCL5,CD14,CEBPA,CEBPB,COL13A1,CSF2,CYP24A1,IGFBP5,IGFBP6,IL12A,IL1RL1,KLF4,PRKCB,PRKCE,PRKCH,PRKD3,RXRA,TGFB2,THBD |
| Agrin Interactions at Neuromuscular Junction | 3.14E+00 | ACTA2,ACTC1,ARHGEF6,ARHGEF7,CDC42,ITGA4,ITGA5,ITGAL,ITGB2,JUN,LAMA2,MAPK12,MRAS,NRAS,RAC2,RALB,RAP2B,UTRN |
| Death Receptor Signaling | 3.11E+00 | ACTA2,ACTC1,ARHGDIB,CASP10,CASP8,FAS,GAS2,HSPB3,IKBKB,MAP3K14,MAP4K4,NFKBIE,PARP12,PARP8,RELA,TNF,TNFRSF10B,TNFRSF1B,TNFSF10,TNFSF12 |
| Chemokine Signaling | 2.99E+00 | CALML5,CAMK1,CAMK1D,CAMK1G,CCL13,CCL2,CCL4,CCL5,CXCR4,JUN,MAPK11,MAPK12,MAPK14,MRAS,NRAS,PRKCB,RALB,RAP2B |
| Opioid Signaling Pathway | 2.98E+00 | ADCY1,ADCY3,AP2S1,ARRB1,ARRB2,BLK,CACNA1C,CALML5,CAMK1,CAMK1D,CAMK1G,CDC42,FGR,GNG11,GNG7,HCK,ITPR3,LCK,LYN,MAPK12,MRAS,MYC,NOS3,NRAS,PENK,PRKCB,PRKCE,PRKCH,PRKD3,RAC2,RALB,RAP2B,RGS16,RGS19,RGS3,RGS4,RPS6KA3,RPS6KB2,SIGMAR1 |
| **Endothelin-1 Signaling** | 2.96E+00 | ADCY1,ADCY3,CASP1,CASP10,CASP4,CASP8,EDNRA,GNA14,ITPR3,JUN,MAPK11,MAPK12,MAPK14,MRAS,MYC,NOS3,NRAS,PIK3C2B,PIK3R3,PLA2G4A,PLA2G4C,PLA2G7,PLAAT4,PRKCB,PRKCE,PRKCH,PRKD3,PTGS1,RALB,RAP2B |

| | | |
|---|---|---|
| Molecular Mechanisms of Cancer | 2.94E+00 | ADCY1,ADCY3,ARHGEF15,ARHGEF2,ARHGEF6,ARHGEF7,BAK1,BAX,BRCA1,CASP10,CASP8,CCND1,CCNE2,CDC25C,CDC42,CDK1,CDK12,CDKN2B,E2F3,FAS,FZD4,GNA14,ITGA4,ITGA5,JUN,MAPK11,MAPK12,MAPK14,MAX,MRAS,MYC,NFKBIE,NOTCH1,NRAS,PIK3C2B,PIK3R3,PMAIP1,PRKCB,PRKCE,PRKCH,PRKD3,RAC2,RALB,RAP2B,RELA,RHOBTB1,RHOC,RND3,SMAD7,SMAD9,TCF3,TGFB1,TGFB2,TGFBR2,TP53,WNT5B |
| Thrombin Signaling | 2.93E+00 | ADCY1,ADCY3,ARHGEF15,ARHGEF2,ARHGEF6,CAMK1,CAMK1D,CAMK1G,CDC42,GATA2,GATA3,GNA14,GNG11,GNG7,IKBKB,ITPR3,MAPK11,MAPK12,MAPK14,MRAS,NRAS,PIK3C2B,PIK3R3,PRKCB,PRKCE,PRKCH,PRKD3,RAC2,RALB,RAP2B,RELA,RHOBTB1,RHOC,RND3 |
| PI3K Signaling in B Lymphocytes | 2.90E+00 | BLK,BTK,CALML5,CD180,CD40,CD79B,DAPP1,IKBKB,IL4R,INPP5D,ITPR3,JUN,LYN,MRAS,NFATC4,NFKBIE,NRAS,PRKCB,PTEN,PTPRC,RALB,RAP2B,RELA,TLR4,VAV1 |
| GŒ±q Signaling | 2.85E+00 | AGTR1,BTK,CALML5,CDC42,CSK,GNA14,GNG11,GNG7,GRM5,IKBKB,ITPR3,MRAS,NFATC4,NFKBIE,PIK3C2B,PIK3R3,PRKCB,PRKCE,PRKCH,PRKD3,RAC2,RELA,RGS16,RGS4,RHOBTB1,RHOC,RND3 |
| Tumoricidal Function of Hepatic Natural Killer Cells | 2.82E+00 | BAX,CASP8,FAS,GZMB,ICAM1,ITGAL,M6PR,PRF1,SRGN |
| VEGF Family Ligand-Receptor Interactions | 2.79E+00 | FLT4,KDR,MRAS,NOS3,NRAS,NRP1,NRP2,PGF,PIK3C2B,PIK3R3,PLA2G4A,PLA2G4C,PRKCB,PRKCE,PRKCH,PRKD3,RALB,RAP2B |
| Nur77 Signaling in T Lymphocytes | 2.75E+00 | CALML5,CD247,CD3D,FCER1G,HDAC9,HLA-DMA,HLA-DMB,HLA-DOA,HLA-DQA1,HLA-DQB1,HLA-DRA,HLA-DRB1,RXRA |
| Role of Tissue Factor in Cancer | 2.75E+00 | ARRB1,ARRB2,BLK,CDC42,CSF2,FGR,GNA14,HCK,IL1B,LCK,LYN,MAPK11,MAPK12,MAPK14,MRAS,NRAS,PIK3C2B,PIK3R3,PTEN,RALB,RAP2B,RPS6KA3,TP53 |
| Sperm Motility | 2.74E+00 | BLK,BTK,CALML5,CNGA1,CSF1R,CSK,EPHB4,FES,FGR,FLT4,HCK,ITK,ITPR3,KDR,LCK,LTK,LYN,MAP3K11,MATK,MRAS,PDE2A,PDGFRA,PLA2G4A,PLA2G4C,PLA2G7,PLAAT4,PRKCB,PRKCE,PRKCH,PRKD3,RET,ROR2,TEK,TIE1 |
| Macropinocytosis Signaling | 2.71E+00 | CD14,CDC42,CSF1R,ITGA5,ITGB2,MRAS,NRAS,PDGFB,PDGFD,PIK3C2B,PIK3R3,PRKCB,PRKCE,PRKCH,PRKD3,RALB,RAP2B |

| | | |
|---|---|---|
| HGF Signaling | 2.66E+00 | CCND1,CDC42,ELF4,ELK3,ETS1,ETS2,ITGA4,ITGA5,JUN,MAP3K11,MAP3K14,MAPK12,MRAS,NRAS,PIK3C2B,PIK3R3,PRKCB,PRKCE,PRKCH,PRKD3,RALB,RAP2B |
| IL-10 Signaling | 2.61E+00 | CD14,IKBKB,IL10RA,IL1B,IL1R1,IL1RL1,IL4R,JUN,MAP3K14,MAP4K4,MAPK11,MAPK12,MAPK14,NFKBIE,RELA,TNF |
| Virus Entry via Endocytic Pathways | 2.61E+00 | ACTA2,ACTC1,AP1S2,AP2S1,CDC42,HLA-E,ITGA4,ITGA5,ITGAL,ITGB2,MRAS,NRAS,PIK3C2B,PIK3R3,PRKCB,PRKCE,PRKCH,PRKD3,RAC2,RALB,RAP2B |
| Regulation of IL-2 Expression in Activated and Anergic T Lymphocytes | 2.61E+00 | CALML5,CD247,CD3D,IKBKB,JUN,LAT,MAPK12,MRAS,NFATC4,NFKBIE,NRAS,RALB,RAP2B,RELA,TGFB1,TGFB2,TGFBR2,VAV1 |
| UVC-Induced MAPK Signaling | 2.59E+00 | JUN,MAPK11,MAPK12,MAPK14,MRAS,NRAS,PRKCB,PRKCE,PRKCH,PRKD3,RALB,RAP2B,TP53 |
| IL-15 Production | 2.56E+00 | BLK,BTK,CSF1R,CSK,EPHB4,FES,FGR,FLT4,HCK,IRF1,ITK,KDR,LCK,LTK,LYN,MAP3K11,MATK,PDGFRA,RELA,RET,ROR2,TEK,TIE1 |
| IL-15 Signaling | 2.56E+00 | CSF2,IL15RA,IL2RB,IL2RG,LCK,MAPK11,MAPK12,MAPK14,MRAS,NRAS,PIK3C2B,PIK3R3,RALB,RAP2B,RELA,TNF |
| MIF Regulation of Innate Immunity | 2.55E+00 | CD14,CD74,JUN,LY96,MAPK12,NFKBIE,PLA2G4A,PLA2G4C,RELA,TLR4,TP53 |
| Mechanisms of Viral Exit from Host Cells | 2.55E+00 | ACTA2,ACTC1,LMNB1,LMNB2,NEDD4,PRKCB,PRKCE,PRKCH,PRKD3,SH3GL1,SH3GLB1 |
| Inhibition of Angiogenesis by TSP1 | 2.50E+00 | HSPG2,JUN,KDR,MAPK11,MAPK12,MAPK14,NOS3,TGFB1,TGFBR2,TP53 |
| Differential Regulation of Cytokine Production in Macrophages and T Helper Cells by IL-17A and IL-17F | 2.49E+00 | CCL2,CCL4,CCL5,CSF2,IL12A,IL1B,TNF |
| Renin-Angiotensin Signaling | 2.47E+00 | ADCY1,ADCY3,AGTR1,CCL2,CCL5,ITPR3,JUN,MAPK11,MAPK12,MAPK14,MRAS,NRAS,PIK3C2B,PIK3R3,PRKCB,PRKCE,PRKCH,PRKD3,RALB,RAP2B,RELA,TNF |
| Caveolar-mediated Endocytosis Signaling | 2.43E+00 | ACTA2,ACTC1,CAVIN1,CD48,FLOT1,FLOT2,HLA-E,ITGA10,ITGA4,ITGA5,ITGA8,ITGAL,ITGAM,ITGAX,ITGB2,PTPN1 |
| GM-CSF Signaling | 2.33E+00 | BCL2A1,CCND1,CSF2,ETS1,HCK,LYN,MRAS,NRAS,PIK3C2B,PIK3R3,PIM1,PRKCB,RALB,RAP2B,RUNX1 |
| Acute Myeloid Leukemia Signaling | 2.33E+00 | CCND1,CEBPA,CSF1R,CSF3R,MRAS,MYC,NRAS,PIK3C2B,PIK3R3,PIM1,PML,RALB,RAP2B,RELA,RPS6KB2,RUNX1,SPI1,TCF3 |

| | | |
|---|---|---|
| p38 MAPK Signaling | 2.32E+00 | FAS,HSPB3,IL1B,IL1R1,IL1RL1,MAPK11,MAPK12,MAPK14,MAX,MEF2C,MYC,PLA2G4A,PLA2G4C,RPS6KA3,RPS6KB2,TGFB1,TGFB2,TGFBR2,TNF,TNFRSF1B,TP53 |
| Role of MAPK Signaling in the Pathogenesis of Influenza | 2.20E+00 | BAX,CCL2,CCL5,MAPK11,MAPK12,MAPK14,MRAS,NRAS,PLA2G4A,PLA2G4C,PLA2G7,PLAAT4,RALB,RAP2B,TNF |
| Complement System | 2.20E+00 | C1QA,C1QB,C2,C3AR1,C5AR1,CFD,ITGAM,ITGAX,ITGB2,MBL2 |
| CXCR4 Signaling | 2.18E+00 | ADCY1,ADCY3,CD4,CDC42,CXCR4,ELMO1,GNA14,GNG11,GNG7,ITPR3,JUN,LYN,MAPK12,MRAS,NRAS,PIK3C2B,PIK3R3,PRKCB,PRKCE,PRKCH,PRKD3,RAC2,RALB,RAP2B,RHOBTB1,RHOC,RND3 |
| PPAR Signaling | 2.16E+00 | IKBKB,IL1B,IL1R1,IL1RL1,JUN,MAP3K14,MAP4K4,MRAS,NFKBIE,NRAS,PDGFB,PDGFD,PDGFRA,RALB,RAP2B,RELA,RXRA,TNF,TNFRSF1B |
| Germ Cell-Sertoli Cell Junction Signaling | 2.11E+00 | A2M,ACTA2,ACTC1,ACTN1,AFDN,CDC42,MAP3K11,MAP3K14,MAPK12,MAPK14,MRAS,NRAS,PIK3C2B,PIK3R3,RAC2,RALB,RAP2B,RHOBTB1,RHOC,RND3,SORBS1,TGFB1,TGFB2,TGFBR2,TNF,TUBA1A,TUBB6 |
| Ephrin Receptor Signaling | 2.09E+00 | ARHGEF15,ARPC1B,ARPC2,CDC42,CXCR4,EFNA1,EFNB3,EPHB4,GNA14,GNG11,GNG7,ITGA4,ITGA5,MAP3K14,MAP4K4,MRAS,NCK1,NRAS,PDGFB,PDGFD,PGF,RAC2,RALB,RAP2B,RGS3,SORBS1,WAS,WIPF1 |
| **Angiopoietin Signaling** | 2.08E+00 | BIRC5,DOK2,IKBKB,MRAS,NCK1,NFKBIE,NOS3,NRAS,PIK3C2B,PIK3R3,RALB,RAP2B,RELA,TEK,TIE1 |
| CD40 Signaling | 2.04E+00 | CD40,ICAM1,IKBKB,JUN,MAP3K14,MAPK11,MAPK12,MAPK14,NFKBIE,PIK3C2B,PIK3R3,PTGS1,RELA,TNFAIP3 |
| Breast Cancer Regulation by Stathmin1 | 2.04E+00 | ADCY1,ADCY3,ARHGEF15,ARHGEF2,ARHGEF6,ARHGEF7,CALML5,CAMK1,CAMK1D,CAMK1G,CCNE2,CDC42,CDK1,E2F3,GNG11,GNG7,ITPR3,MRAS,NRAS,PIK3C2B,PIK3R3,PRKCB,PRKCE,PRKCH,PRKD3,RALB,RAP2B,TP53,TUBA1A,TUBB6 |
| IL-7 Signaling Pathway | 2.02E+00 | BAK1,BAX,BCL6,CCND1,IL2RG,IL7,JUN,LYN,MAPK11,MAPK12,MAPK14,MCL1,MYC,PIK3C2B,PIK3R3 |
| Reelin Signaling in Neurons | 2.02E+00 | ARHGEF15,ARHGEF2,ARHGEF6,BLK,FGR,HCK,ITGA4,ITGA5,ITGAL,ITGB2,LCK,LYN,MAP3K11,MAPK12,PIK3C2B,PIK3R3 |
| Prolactin Signaling | 2.02E+00 | CEBPB,IRF1,JUN,MRAS,MYC,NMI,NRAS,PIK3C2B,PIK3R3,PRKCB,PRKCE,PRKCH,PRKD3,RALB,RAP2B,SOCS2 |
| Sumoylation Pathway | 2.02E+00 | AR,ARHGDIB,CDC42,CEBPA,DNMT3A,ETS1,FAS,ISG20,JUN,MAPK12,PML,RAC2,RFC2,RHOBTB1,RHOC,RND3,SP100,TP53,UBE2I |

| | | |
|---|---|---|
| Actin Cytoskeleton Signaling | 1.99E+00 | ACTA2,ACTC1,ACTN1,ARHGEF6,ARHGEF7,ARPC1B,ARPC2,CD14,CDC42,CSK,FGD1,FGF2,FGF8,FN1,ITGA4,ITGA5,MATK,MRAS,MYH11,NCKAP1L,NRAS,PDGFB,PDGFD,PFN1,PIK3C2B,PIK3R3,RAC2,RALB,RAP2B,TMSB10/TMSB4X,VAV1,WAS |
| Role of NFAT in Cardiac Hypertrophy | 1.99E+00 | ADCY1,ADCY3,CACNA1C,CALML5,CAMK1,CAMK1D,CAMK1G,GNG11,GNG7,HDAC7,HDAC9,ITPR3,MAPK11,MAPK12,MAPK14,MEF2C,MRAS,NFATC4,NRAS,PIK3C2B,PIK3R3,PRKCB,PRKCE,PRKCH,PRKD3,RALB,RAP2B,RCAN2,TGFB1,TGFB2,TGFBR2 |
| UVB-Induced MAPK Signaling | 1.99E+00 | JUN,MAPK11,MAPK12,MAPK14,PIK3C2B,PIK3R3,PRKCB,PRKCE,PRKCH,PRKD3,RPS6KA3,TP53 |
| PAK Signaling | 1.98E+00 | ARHGEF6,ARHGEF7,CDC42,ITGA4,ITGA5,MAPK12,MRAS,NCK1,NRAS,PAK1IP1,PDGFB,PDGFD,PDGFRA,PIK3C2B,PIK3R3,RALB,RAP2B,TNF |
| **Epithelial Adherens Junction Signaling** | 1.98E+00 | ACTA2,ACTC1,ACTN1,AFDN,ARPC1B,ARPC2,CDC42,MRAS,MYH11,NOTCH1,NOTCH3,NOTCH4,NRAS,PTEN,PTPRM,RALB,RAP2B,SNAI2,SORBS1,TCF3,TGFB2,TGFBR2,TUBA1A,TUBB6,WAS |
| PEDF Signaling | 1.98E+00 | CASP8,FAS,IKBKB,MAPK11,MAPK12,MAPK14,MRAS,NFKBIE,NRAS,PIK3C2B,PIK3R3,RALB,RAP2B,RELA,SOD2,TP53 |
| Differential Regulation of Cytokine Production in Intestinal Epithelial Cells by IL-17A and IL-17F | 1.96E+00 | CCL2,CCL4,CCL5,CSF2,IL12A,IL1B,TNF |
| SAPK/JNK Signaling | 1.94E+00 | CDC42,FCER1G,GNG11,GNG7,JUN,LCK,MAP3K11,MAP4K4,MAPK12,MINK1,MRAS,NRAS,PIK3C2B,PIK3R3,RAC2,RALB,RAP2B,TP53 |
| HER-2 Signaling in Breast Cancer | 1.94E+00 | CCND1,CCNE2,CDC42,ITGB2,MMP2,MRAS,NRAS,PIK3C2B,PIK3R3,PRKCB,PRKCE,PRKCH,PRKD3,RALB,RAP2B,TP53 |
| Cholecystokinin/Gastrin-mediated Signaling | 1.92E+00 | CDC42,IL1B,ITPR3,JUN,MAPK12,MAPK14,MEF2C,MRAS,NRAS,PRKCB,PRKCE,PRKCH,PRKD3,RAC2,RALB,RAP2B,RHOBTB1,RHOC,RND3,TNF |
| Role of Osteoblasts, Osteoclasts and Chondrocytes in Rheumatoid Arthritis | 1.91E+00 | CALML5,COL1A1,CSF1R,CSF2,CTSK,DKK1,DKK3,FRZB,FZD4,IKBKB,IL1B,IL1R1,IL1RL1,IL7,ITGA5,JUN,MAP3K14,MAPK12,MAPK14,MMP3,MMP8,NFATC4,NFKBIE,PIK3C2B,PIK3R3,RELA,SMAD9,TCF3,TGFB1,TNF,TNFRSF1B,WNT5B |
| p53 Signaling | 1.90E+00 | BAX,BIRC5,BRCA1,CCND1,FAS,HDAC9,JUN,MAPK14,PIK3C2B,PIK3R3,PMAIP1,PML,PPP1R13B,PTEN,SCO2,SNAI2,TNFRSF10B,TP53 |

| | | |
|---|---|---|
| Glioma Invasiveness Signaling | 1.90E+00 | CD44,CDC42,MMP2,MRAS,NRAS,PIK3C2B,PIK3R3,PLAU,RAC2,RALB,RAP2B,RHOBTB1,RHOC,RND3 |
| Wnt/Œ≤-catenin Signaling | 1.90E+00 | CCND1,CD44,CDH5,DKK1,DKK3,FRZB,FZD4,JUN,MYC,NR5A2,RARB,RARG,SOX11,SOX13,SOX17,SOX18,SOX4,SOX9,TCF3,TGFB1,TGFB2,TGFBR2,TLE1,TLE3,TP53,WNT5B |
| IL-17A Signaling in Fibroblasts | 1.90E+00 | CCL2,CEBPB,IKBKB,JUN,MAPK11,MAPK12,MAPK14,NFKBIE,RELA |
| Induction of Apoptosis by HIV1 | 1.86E+00 | BAK1,BAX,CASP8,CXCR4,FAS,IKBKB,MAP3K14,MAPK12,NFKBIE,RELA,TNF,TNFRSF1B,TP53 |
| Role of PKR in Interferon Induction and Antiviral Response | 1.83E+00 | CASP8,EIF2AK2,EIF2S1,IKBKB,IRF1,MAPK14,NFKBIE,RELA,TNF,TP53 |
| Cytotoxic T Lymphocyte-mediated Apoptosis of Target Cells | 1.83E+00 | CASP8,CD247,CD3D,FAS,FCER1G,GZMB,HLA-E,PRF1 |
| **TNFR2 Signaling** | 1.83E+00 | IKBKB,JUN,MAP3K14,NFKBIE,RELA,TNF,TNFAIP3,TNFRSF1B |
| Activation of IRF by Cytosolic Pattern Recognition Receptors | 1.81E+00 | CD40,IFIH1,IFIT2,IKBKB,IRF7,IRF9,ISG15,JUN,MAPK12,NFKBIE,RELA,TNF,ZBP1 |
| FAK Signaling | 1.81E+00 | ACTA2,ACTC1,ARHGAP26,ARHGEF6,ARHGEF7,ASAP1,CSK,ITGA4,ITGA5,MRAS,NRAS,PIK3C2B,PIK3R3,PTEN,RALB,RAP2B,WAS |
| Sertoli Cell-Sertoli Cell Junction Signaling | 1.81E+00 | A2M,ACTA2,ACTC1,ACTN1,AFDN,CDC42,EPB41,ITGA4,ITGA5,JAM2,JUN,MAP3K11,MAP3K14,MAPK11,MAPK12,MAPK14,MRAS,NOS3,NRAS,PTEN,RALB,RAP2B,SORBS1,TNF,TUBA1A,TUBB6,WAS |
| Airway Pathology in Chronic Obstructive Pulmonary Disease | 1.80E+00 | CXCL3,MMP2,MMP8,TNF |
| Sphingosine-1-phosphate Signaling | 1.79E+00 | ADCY1,ADCY3,CASP1,CASP10,CASP4,CASP8,CDC42,PDGFB,PDGFD,PDGFRA,PIK3C2B,PIK3R3,RAC2,RHOBTB1,RHOC,RND3,S1PR1,S1PR4 |
| Mouse Embryonic Stem Cell Pluripotency | 1.79E+00 | FZD4,ID1,ID2,ID3,LIFR,MAPK11,MAPK12,MAPK14,MRAS,MYC,NRAS,PIK3C2B,PIK3R3,RALB,RAP2B,SMAD9,TCF3,TP53 |
| HIF1Œ± Signaling | 1.76E+00 | JUN,LDHA,MAPK11,MAPK12,MAPK14,MMP2,MMP3,MMP8,MRAS,NAA10,NOS3,NRAS,PGF,PIK3C2B,PIK3R3,RALB,RAP2B,SLC2A3,TP53 |
| Coagulation System | 1.76E+00 | A2M,F12,F8,PLAT,PLAU,SERPINA1,TFPI,THBD,VWF |
| IL-17 Signaling | 1.76E+00 | CCL2,CEBPB,JUN,MAP3K14,MAPK11,MAPK12,MAPK14,MMP3,MRAS,NRAS,PIK3C2B,PIK3R3,RALB,RAP2B,RELA |

| | | |
|---|---|---|
| Small Cell Lung Cancer Signaling | 1.76E+00 | CCND1,CCNE2,CDKN2B,IKBKB,MAX,MYC,NFKBIE,PIK3C2B,PIK3R3,PTEN,RARB,RELA,RXRA,TP53 |
| MIF-mediated Glucocorticoid Regulation | 1.76E+00 | CD14,CD74,LY96,NFKBIE,PLA2G4A,PLA2G4C,RELA,TLR4 |
| MSP-RON Signaling Pathway | 1.72E+00 | ACTA2,ACTC1,CCL2,CCR2,F12,IL12A,ITGAM,ITGB2,PIK3C2B,PIK3R3,TLR4,TNF |
| Apelin Liver Signaling Pathway | 1.68E+00 | APLNR,COL11A2,COL1A1,COL1A2,FAS,MAPK12,TNF |
| Glioblastoma Multiforme Signaling | 1.67E+00 | CCND1,CDC42,E2F3,FZD4,IGF2,ITPR3,MRAS,MYC,NRAS,PDGFB,PDGFD,PDGFRA,PIK3C2B,PIK3R3,PTEN,RAC2,RALB,RAP2B,RHOBTB1,RHOC,RND3,TCF3,TP53,WNT5B |
| **IL-1 Signaling** | 1.63E+00 | ADCY1,ADCY3,GNA14,GNG11,GNG7,IKBKB,IL1R1,JUN,MAP3K14,MAPK11,MAPK12,MAPK14,MRAS,MYD88,NFKBIE,RELA |
| Adipogenesis pathway | 1.63E+00 | ATG7,CEBPA,CEBPB,EGR2,EZH2,FABP4,FGF2,FZD4,GTF2H5,HDAC7,HDAC9,KLF3,NFATC4,RBBP4,RBP1,SAP30,SMAD9,SOX9,TGFB1,TNF,TP53 |
| B Cell Activating Factor Signaling | 1.62E+00 | IKBKB,JUN,MAP3K14,MAPK11,MAPK12,MAPK14,NFATC4,NFKBIE,RELA |
| CCR3 Signaling in Eosinophils | 1.62E+00 | CALML5,GNG11,GNG7,ITPR3,MAPK11,MAPK12,MAPK14,MRAS,NRAS,PIK3C2B,PIK3R3,PLA2G4A,PLA2G4C,PRKCB,PRKCE,PRKCH,PRKD3,RALB,RAP2B |
| April Mediated Signaling | 1.62E+00 | IKBKB,JUN,MAP3K14,MAPK11,MAPK12,MAPK14,NFATC4,NFKBIE,RELA |
| Signaling by Rho Family GTPases | 1.62E+00 | ACTA2,ACTC1,ARHGEF15,ARHGEF2,ARHGEF6,ARHGEF7,ARPC1B,ARPC2,CDC42,CDH11,CDH13,CDH5,CYBB,GNA14,GNG11,GNG7,ITGA4,ITGA5,JUN,MAP3K11,MAPK12,MRAS,NCF2,NEDD4,PIK3C2B,PIK3R3,RAC2,RELA,RHOBTB1,RHOC,RND3,WAS,WIPF1 |
| RhoGDI Signaling | 1.62E+00 | ACTA2,ACTC1,ARHGAP4,ARHGDIB,ARHGEF15,ARHGEF2,ARHGEF6,ARHGEF7,ARPC1B,ARPC2,CD44,CDC42,CDH11,CDH13,CDH5,DGKZ,GNA14,GNG11,GNG7,ITGA4,ITGA5,MRAS,RAC2,RHOBTB1,RHOC,RND3 |
| 4-1BB Signaling in T Lymphocytes | 1.60E+00 | IKBKB,JUN,MAP3K14,MAPK11,MAPK12,MAPK14,NFKBIE,RELA |
| IL-17A Signaling in Gastric Cells | 1.60E+00 | CCL5,JUN,MAPK11,MAPK12,MAPK14,RELA,TNF |
| GNRH Signaling | 1.59E+00 | ADCY1,ADCY3,CACNA1C,CALML5,CDC42,GNA14,GNG11,GNG7,ITPR3,JUN,MAP3K11,MAP3K14,MAPK11,MAPK12,MAPK14,MMP2,MRAS,NRAS,PRKCB,PRKCE,PRKCH,PRKD3,RALB,RAP2B,RELA |

| | | |
|---|---|---|
| Chronic Myeloid Leukemia Signaling | 1.59E+00 | CCND1,E2F3,HDAC7,HDAC9,IKBKB,MRAS,MYC,NRAS,PIK3C2B,PIK3R3,RALB,RAP2B,RELA,TGFB1,TGFB2,TGFBR2,TP53 |
| ErbB Signaling | 1.57E+00 | CDC42,JUN,MAPK11,MAPK12,MAPK14,MRAS,NCK1,NRAS,PIK3C2B,PIK3R3,PRKCB,PRKCE,PRKCH,PRKD3,RALB,RAP2B |
| Adrenomedullin signaling pathway | 1.57E+00 | ADCY1,ADCY3,BAX,CALCRL,CALML5,CEBPB,CSK,GNA14,IL1B,ITPR3,MAPK11,MAPK12,MAPK14,MATK,MAX,MMP2,MRAS,NOS3,NRAS,PIK3C2B,PIK3R3,RALB,RAMP2,RAP2B,RELA,RXRA,TNF |
| Telomerase Signaling | 1.55E+00 | ELF4,ELK3,ETS1,ETS2,HDAC7,HDAC9,IL2RB,IL2RG,MRAS,MYC,NRAS,PIK3C2B,PIK3R3,RALB,RAP2B,TINF2,TP53 |
| 3-phosphoinositide Degradation | 1.55E+00 | CDC25C,DOT1L,DUSP11,DUSP2,DUSP5,INPP4B,INPP5D,INPP5K,NUDT1,NUDT11,NUDT3,PPFIA3,PPP1R13B,PTEN,PTPN1,PTPRC,PTPRH,PTPRM,STYXL1,TPTE |
| PTEN Signaling | 1.55E+00 | CCND1,CDC42,FLT4,IKBKB,INPP5D,INPP5K,ITGA4,ITGA5,KDR,MRAS,NRAS,PDGFRA,PIK3R3,PTEN,RAC2,RALB,RAP2B,RELA,RPS6KB2,TGFBR2 |
| Thrombopoietin Signaling | 1.52E+00 | JUN,MRAS,MYC,NRAS,PIK3C2B,PIK3R3,PRKCB,PRKCE,PRKCH,PRKD3,RALB,RAP2B |
| GŒ±12/13 Signaling | 1.52E+00 | BTK,CDC42,CDH11,CDH13,CDH5,IKBKB,JUN,LPAR6,MAPK12,MEF2C,MRAS,NFKBIE,NRAS,PIK3C2B,PIK3R3,RALB,RAP2B,RELA,TBXA2R,VAV1 |
| Glucocorticoid Receptor Signaling | 1.50E+00 | A2M,ADRB2,AR,CCL13,CCL2,CCL5,CD163,CD247,CD3D,CEBPA,CEBPB,CSF2,CXCL3,GTF2H5,ICAM1,IKBKB,IL1B,JUN,KRT17,MAP3K14,MAPK11,MAPK12,MAPK14,MRAS,NFATC4,NFKBIE,NRAS,PBX1,PIK3C2B,PIK3R3,PLAU,POLR2D,RALB,RAP2B,RELA,TAF12,TGFB1,TGFB2,TGFBR2,TNF,UBE2I,VIPR1 |
| RAR Activation | 1.49E+00 | ADCY1,ADCY3,AKR1B10,CSK,GTF2H5,JUN,MAPK11,MAPK12,MAPK14,PIK3R3,PML,PRKCB,PRKCE,PRKCH,PRKD3,PTEN,RARB,RARG,RBP1,RBP3,RELA,RXRA,SMAD7,SMAD9,TGFB1,TGFB2,TRIM24 |
| Thyroid Cancer Signaling | 1.48E+00 | CCND1,MRAS,MYC,NRAS,RALB,RAP2B,RET,RXRA,TCF3,TP53 |
| Myc Mediated Apoptosis Signaling | 1.48E+00 | BAX,CASP8,FAS,MAPK12,MRAS,MYC,NRAS,PIK3C2B,PIK3R3,RALB,RAP2B,TP53 |
| PDGF Signaling | 1.48E+00 | EIF2AK2,INPP5D,INPP5K,JUN,MRAS,MYC,NRAS,PDGFB,PDGFD,PDGFRA,PIK3C2B,PIK3R3,PRKCB,RALB,RAP2B |

| | | |
|---|---|---|
| Systemic Lupus Erythematosus Signaling | 1.46E+00 | CD247,CD3D,CD40,CD79B,FCER1G,FCGR3A/FCGR3B,HLA-E,IL1B,INPP5D,JUN,LAT,LCK,LSM7,LYN,MRAS,NFATC4,NRAS,PIK3C2B,PIK3R3,PRPF38B,PTPRC,RALB,RAP2B,SNRNP40,TLR7,TNF |
| Superpathway of Inositol Phosphate Compounds | 1.46E+00 | CDC25C,DOT1L,DUSP11,DUSP2,DUSP5,INPP5A,INPP5D,INPP5K,ITPKB,NUDT1,NUDT11,NUDT3,PIK3C2B,PIK3R3,PIP4K2A,PPFIA3,PPP1R13B,PTEN,PTPN1,PTPRC,PTPRH,PTPRM,STYXL1,TPTE |
| **TGF-Œ≤ Signaling** | 1.46E+00 | CDC42,IRF7,JUN,MAPK11,MAPK12,MAPK14,MRAS,NRAS,RALB,RAP2B,RUNX3,SMAD7,SMAD9,TGFB1,TGFB2,TGFBR2 |
| Regulation of the Epithelial-Mesenchymal Transition Pathway | 1.44E+00 | ETS1,FGF2,FGF8,FZD4,HMGA2,ID2,JAG2,MMP2,MRAS,NOTCH1,NOTCH3,NOTCH4,NRAS,PDGFD,PIK3C2B,PIK3R3,RALB,RAP2B,RELA,SNAI2,TCF3,TGFB1,TGFB2,TGFBR2,TWIST1,WNT5B |
| Nitric Oxide Signaling in the Cardiovascular System | 1.42E+00 | ATP2A3,CACNA1C,CALML5,FLT4,ITPR3,KDR,NOS3,PDE2A,PDE5A,PGF,PIK3C2B,PIK3R3,PRKCB,PRKCE,PRKCH,PRKD3 |
| Role of JAK1 and JAK3 in Œ≥c Cytokine Signaling | 1.39E+00 | FES,IL15RA,IL2RB,IL2RG,IL4R,IL7,MRAS,NRAS,PIK3C2B,PIK3R3,RALB,RAP2B |
| Cell Cycle: G1/S Checkpoint Regulation | 1.39E+00 | CCND1,CCNE2,CDKN2B,E2F3,HDAC7,HDAC9,MAX,MYC,PAK1IP1,TGFB1,TGFB2,TP53 |
| Granzyme B Signaling | 1.39E+00 | CASP8,GZMB,LMNB1,LMNB2,PRF1 |
| **VEGF Signaling** | 1.39E+00 | ACTA2,ACTC1,ACTN1,EIF2B4,EIF2S1,FLT4,KDR,MRAS,NOS3,NRAS,PGF,PIK3C2B,PIK3R3,PRKCB,RALB,RAP2B |
| **Axonal Guidance Signaling** | 1.38E+00 | ADAM12,ADAM15,ADAMTS1,ARHGEF15,ARHGEF6,ARHGEF7,ARPC1B,ARPC2,CDC42,CXCR4,EFNA1,EFNB3,EPHB4,FES,FZD4,GNA14,GNG11,GNG7,ITGA4,ITGA5,MMP2,MMP3,MMP8,MRAS,NCK1,NFATC4,NRAS,NRP1,NRP2,PDGFB,PDGFD,PFN1,PGF,PIK3C2B,PIK3R3,PLXNB3,PRKCB,PRKCE,PRKCH,PRKD3,RAC2,RALB,RAP2B,RGS3,SEMA3F,SEMA4C,SEMA6A,TUBA1A,TUBB6,VASP,WAS,WIPF1,WNT5B |
| Œ±-Adrenergic Signaling | 1.37E+00 | ADCY1,ADCY3,CALML5,GNG11,GNG7,ITPR3,MRAS,NRAS,PRKCB,PRKCE,PRKCH,PRKD3,PYGL,RALB,RAP2B |
| GDNF Family Ligand-Receptor Interactions | 1.37E+00 | CDC42,DOK2,ITPR3,JUN,MAPK12,MRAS,NCK1,NRAS,PIK3C2B,PIK3R3,RALB,RAP2B,RET |
| Superpathway of D-myo-inositol (1,4,5)-trisphosphate Metabolism | 1.37E+00 | INPP1,INPP5A,INPP5D,INPP5K,ITPKB,PTEN |

| | | |
|---|---|---|
| UVA-Induced MAPK Signaling | 1.37E+00 | JUN,MAPK11,MAPK12,MAPK14,MRAS,NRAS,PARP12,PARP8,PIK3C2B,PIK3R3,RALB,RAP2B,RPS6KA3,RPS6KB2,TP53 |
| Rac Signaling | 1.35E+00 | ARPC1B,ARPC2,CD44,CDC42,CYBB,ITGA4,ITGA5,JUN,MAP3K11,MRAS,NCF2,NRAS,PIK3C2B,PIK3R3,RALB,RAP2B,RELA |
| Hematopoiesis from Pluripotent Stem Cells | 1.35E+00 | CD247,CD3D,CD4,CD8A,CSF2,FCER1G,IL12A,IL7 |
| Leukotriene Biosynthesis | 1.35E+00 | DPEP2,GGT5,LTC4S,MGST2 |
| Erythropoietin Signaling | 1.33E+00 | JUN,MRAS,NFKBIE,NRAS,PIK3C2B,PIK3R3,PRKCB,PRKCE,PRKCH,PRKD3,RALB,RAP2B,RELA |
| Cardiac Hypertrophy Signaling | 1.33E+00 | ADCY1,ADCY3,ADRB2,CACNA1C,CALML5,CDC42,EIF2B4,GNA14,GNG11,GNG7,JUN,MAP3K11,MAP3K14,MAPK11,MAPK12,MAPK14,MEF2C,MRAS,NFATC4,NRAS,PIK3C2B,PIK3R3,RAC2,RALB,RAP2B,RHOBTB1,RHOC,RND3,TGFB1,TGFB2,TGFBR2 |
| Neuregulin Signaling | 1.30E+00 | ITGA4,ITGA5,MATK,MRAS,MYC,NRAS,PIK3R3,PRKCB,PRKCE,PRKCH,PRKD3,PTEN,RALB,RAP2B,RPS6KB2 |
| D-myo-inositol (1,3,4)-trisphosphate Biosynthesis | 1.30E+00 | INPP5A,INPP5D,INPP5K,ITPKB,PTEN |
| Th17 Activation Pathway | 1.30E+00 | CCR6,CSF2,FCER1G,IL12A,IL12RB1,IL1B,IL1R1,MYD88,NFATC4,RELA,RUNX1 |

**Supplemental Table 7**: Clinical and morphometric measures for the participants who contributed single-cell expression data from kidney biopsies

| Characteristic | N | |
|---|---|---|
| Male sex (%) | 44 | 14 (31.8%) |
| Age (years) | 44 | 41.0 ± 11.1 |
| Diabetes duration (years) | 44 | 12.2 ± 7.5 |
| BMI (kg/m$^2$) | 44 | 36.9 ± 7.3 |
| Systolic blood pressure (mmHg) | 44 | 119 ± 12 |
| Diastolic blood pressure (mmHg) | 44 | 72 ± 10 |
| HbA1c (%) * | 42 | 9.2 ± 2.4 |
| GFR (ml/min) | 44 | 159 ± 58 |
| ACR (mg/g) | 44 | 18 (9-48) |
| RAS use (%) | 44 | 19 (43.2%) |
| Mean glomerular volume ($10^6$ μm$^3$) | 38 | 2.55 ± 1.06 |
| Glomerular basement membrane width (nm) | 41 | 457 ± 103 |
| Mesangial fractional volume per glomerulus (%) | 41 | 0.23 ± 0.07 |
| Cortical interstitial fractional volume (%) | 35 | 0.18 ± 0.06 |
| Glomerular filtration surface density (μm$^2$/μm$^3$) | 41 | 0.09 ± 0.02 |
| Foot process width (nm) | 39 | 491 (422-572) |
| Glomerular podocyte fractional volume (%) | 41 | 0.18 ± 0.03 |
| Podocyte number density per glomerulus ($10^6$ μm$^3$) | 41 | 157 ± 98 |
| Fenestrated endothelium (%) | 39 | 47.4 ± 16.1 |

Values are means ± standard deviation or median (interquartile range).
Abbreviations: ACR = albumin:creatinine ratio; BMI = body mass index; GBM = glomerular basement membrane; GFR = glomerular filtration rate; RAS = renin angiotensin system blockers.
